# AI-based differential diagnosis of dementia etiologies on multimodal data

**DOI:** 10.1101/2024.02.08.24302531

**Authors:** Chonghua Xue, Sahana S. Kowshik, Diala Lteif, Shreyas Puducheri, Varuna H. Jasodanand, Olivia T. Zhou, Anika S. Walia, Osman B. Guney, J. Diana Zhang, Serena T. Pham, Artem Kaliaev, V. Carlota Andreu-Arasa, Brigid C. Dwyer, Chad W. Farris, Honglin Hao, Sachin Kedar, Asim Z. Mian, Daniel L. Murman, Sarah A. O’Shea, Aaron B. Paul, Saurabh Rohatgi, Marie-Helene Saint-Hilaire, Emmett A. Sartor, Bindu N. Setty, Juan E. Small, Arun Swaminathan, Olga Taraschenko, Jing Yuan, Yan Zhou, Shuhan Zhu, Cody Karjadi, Ting Fang Alvin Ang, Sarah A. Bargal, Bryan A. Plummer, Kathleen L. Poston, Meysam Ahangaran, Rhoda Au, Vijaya B. Kolachalama

## Abstract

Differential diagnosis of dementia remains a challenge in neurology due to symptom overlap across etiologies, yet it is crucial for formulating early, personalized management strategies. Here, we present an AI model that harnesses a broad array of data, including demographics, individual and family medical history, medication use, neuropsychological assessments, functional evaluations, and multimodal neuroimaging, to identify the etiologies contributing to dementia in individuals. The study, drawing on 51, 269 participants across 9 independent, geographically diverse datasets, facilitated the identification of 10 distinct dementia etiologies. It aligns diagnoses with similar management strategies, ensuring robust predictions even with incomplete data. Our model achieved a micro-averaged area under the receiver operating characteristic curve (AUROC) of 0.94 in classifying individuals with normal cognition, mild cognitive impairment and dementia. Also, the micro-averaged AUROC was 0.96 in differentiating the dementia etiologies. Our model demonstrated proficiency in addressing mixed dementia cases, with a mean AUROC of 0.78 for two cooccurring pathologies. In a randomly selected subset of 100 cases, the AUROC of neurologist assessments augmented by our AI model exceeded neurologist-only evaluations by 26.25%. Furthermore, our model predictions aligned with biomarker evidence and its associations with different proteinopathies were substantiated through postmortem findings. Our framework has the potential to be integrated as a screening tool for dementia in various clinical settings and drug trials, with promising implications for person-level management.

---

Dementia is one of the most pressing health challenges of our time. With nearly 10 million new cases reported annually, this syndrome, characterized by a progressive decline in cognitive function severe enough to impede daily life activities, continues to present considerable clinical and socioeconomic challenges. In 2017, the World Health Organization’s global action plan highlighted the need for prompt and precise diagnosis of dementia as a pivotal strategic objective in response to the growing number of dementia worldwide.^1,2^ As such, diagnostic precision in the varied landscape of dementia remains a critical, yet unmet need, particularly as the global population ages and the demand for more accurate participant screening in drug trials increases.^3^ This challenge primarily stems from the overlapping clinical presentation of different dementia types, which is further complicated by the heterogeneity in findings on magnetic resonance imaging (MRI) scans.^4,5^ The necessity for improvements in the field becomes ever more pressing considering the projected shortage of specialists including neurologists, neuropsychologists and geriatric care providers,^6–8^ emphasizing the urgency to innovate and evolve our diagnostic tools.

Accurate differential diagnosis of dementia is pivotal for prescribing targeted therapeutic interventions, enhancing treatment efficacy and slowing symptom progression. While Alzheimer’s disease (AD) is a leading cause, other forms such as vascular dementia (VD), Lewy body dementia (LBD), and frontotemporal dementia (FTD) are also prevalent.^9–11^ These etiologies can often coexist, as marked by symptom overlap and variable symptom intensity, which further complicate the diagnostic process.^12^ Importantly, diagnostic errors are prevalent among older adults, particularly those with comorbid conditions.^13^ These misdiagnoses can translate into inappropriate medication use and adverse health outcomes.^14^ For example, while patients with early-stage AD may be candidates for anti-amyloid therapies,^15–17^ the coexistence of pathology from other etiologies, such as vascular dementia, can increase the risk of amyloid-related imaging abnormalities.^18^ This highlights the critical need for accurately assessing the full spectrum of etiological factors contributing to dementia to inform appropriate therapeutic strategies and optimize patient care.^19^

The imperative for scalable diagnostic tools in AD and related dementias is becoming increasingly urgent, given the significant challenges in accessing gold-standard testing. Recent regulatory approvals have facilitated the transition of cerebrospinal fluid (CSF) and positron emission tomography (PET) biomarkers from research environments to clinical settings. While promising, the clinical integration of accurate blood-based biomarkers remains an area of active research.^20–22^ Despite these advancements, accessibility to these diagnostic tools is still constrained, not only in remote and economically developing regions but also in urban healthcare centers, as exemplified by prolonged waiting periods for specialist consultations.^23^ This challenge is compounded by a global shortage of specialists, such as behavioral neurologists and neuropsychologists, leading to an overreliance on cognitive assessments that may not be culturally appropriate due to the lack of formal training programs in neuropsychology in many parts of the world.^24,25^ Although conventional methods like clinical evaluations, neuropsychological testing, and MRI remain central to antemortem differential dementia diagnosis, their effectiveness relies on a diminishing pool of specialist clinicians. This underscores an urgent need for healthcare systems to evolve and adapt to the rapidly changing dynamics of dementia diagnosis and treatment.

Machine learning (ML) has the potential to enhance the accuracy and efficiency of dementia diagnosis.^26–28^ Previous ML methods have largely focused on leveraging neuroimaging data to distinguish cognitively normal (NC) individuals from those with mild cognitive impairment (MCI) and dementia (DE), with AD being the main etiology given its ubiquity in dementia diagnosis.^29,30^ A few studies have attempted to discern neuroimaging signatures unique to AD by contrasting them with other dementia types^31–40^ However, this primary emphasis on AD can have limited practical implications given the prevalence and co-occurrence of other etiologies. In addition, a focus on imaging data alone can be insufficient in providing a holistic understanding of an individual’s neurological condition. Recently, we proposed a novel approach to stratify individuals based on cognitive status and discern likely AD cases from non-AD dementia types by incorporating imaging with non-imaging data such as demographics, medical histories, and neuropsychological assessments.^39^ These investigations have begun to illuminate the complex matrix of factors contributing to dementia. However, for ML models to be adopted into clinical practice, they must be able to accommodate the intricacies of mixed etiologies, as well as the inclusion or exclusion of different data modalities that may or may not be available. Therefore, the development of AI methodologies capable of harnessing multimodal data facilitates the accurate quantification of diverse dementia etiologies, irrespective of clinical resources, thereby aligning treatment strategies with individual patient profiles.

In this study, we propose a multimodal machine learning framework that harnesses a diverse array of data, including demographics, personal and family medical history, medication use, neuropsychological assessments, functional evaluations, and multimodal neuroimaging to perform differential dementia diagnosis. Our model, designed to mirror real-world scenarios, aligns diagnoses with similar management strategies and outputs probabilities for each etiology. This approach is intended to mimic clinical reasoning and aid practitioners in dementia screening and treatment planning. The model’s robustness is demonstrated through validation on independent, geographically diverse datasets. In comparative analyses, we found that AI-augmented clinician assessments achieved superior diagnostic accuracy compared to clinician-only assessments. By validating our model against gold-standard biomarker and postmortem data for different etiologies, we further emphasize our model’s ability to dissect the intricate pathophysiology underlying dementia. Our algorithmic framework demonstrates the potential to enhance dementia screening in various clinical settings, illustrating AI’s capacity to improve healthcare outcomes.

## Results

### Glossary 1

Acronym: Description
NC: Normal cognition
MCI: Mild cognitive impairment
DE: Dementia
AD: Alzheimer’s disease
LBD: Lewy body dementia including dementia with Lewy bodies and Parkinson’s disease dementia
VD: Vascular dementia, vascular brain injury, and vascular dementia including stroke
PRD: Prion disease including Creutzfeldt-Jakob disease
FTD: Frontotemporal lobar degeneration and its variants, including primary progressive aphasia, corticobasal degeneration and progressive supranuclear palsy, and with or without amyotrophic lateral sclerosis
NPH: Normal pressure hydrocephalus
SEF: Systemic and environmental factors including infectious diseases (HIV included), metabolic, substance abuse / alcohol, medications, systemic disease, and delirium
PSY: Psychiatric conditions including schizophrenia, depression, bipolar disorder, anxiety, and post-traumatic stress disorder
TBI: Moderate/severe traumatic brain injury, repetitive head injury, and chronic traumatic encephalopathy
ODE: Other dementia conditions including neoplasms, Down syndrome, multiple systems atrophy, Huntington’s disease, seizures, etc.

Leveraging the power of multimodal data obtained from various cohorts (Tables 1 & S1 - S6), our model adopts a nuanced approach to differential dementia diagnosis (Fig. 1). Our framework assigns individuals to one or more of thirteen diagnostic categories (refer to Glossary 1), which were meticulously defined through consensus among a team of expert neurologists. This practical categorization is designed with clinical management pathways in mind, thereby echoing real-world scenarios. For instance, we have grouped dementia with Lewy bodies and Parkinson’s disease dementia under the comprehensive category of Lewy body dementia (LBD). This classification stems from an understanding that the care for these conditions often follows a similar path, typically overseen by a multidisciplinary team of movement disorder specialists. In the context of vascular dementia (VD), we included individuals who exhibited symptoms of a stroke, possible or probable VD, or vascular brain injury. This encompassed cases with symptomatic stroke, cystic infarct in cognitive networks, extensive white matter hyperintensity, and/or executive dysfunction as the primary contributors to the observed cognitive impairment. The inclusion criteria were based on the expectation that such patients would typically receive care from clinicians specializing in stroke and vascular diseases. Likewise, we have considered various psychiatric conditions, such as schizophrenia, depression, bipolar disorders, anxiety, and post-traumatic stress disorder, under one category (PSY), acknowledging that their management predominantly falls within the realm of psychiatric care providers. By aligning diagnostic categories with clinical care pathways, our model serves not only to classify an individual’s condition but also to direct appropriate clinical management strategies.

**Figure 1:**
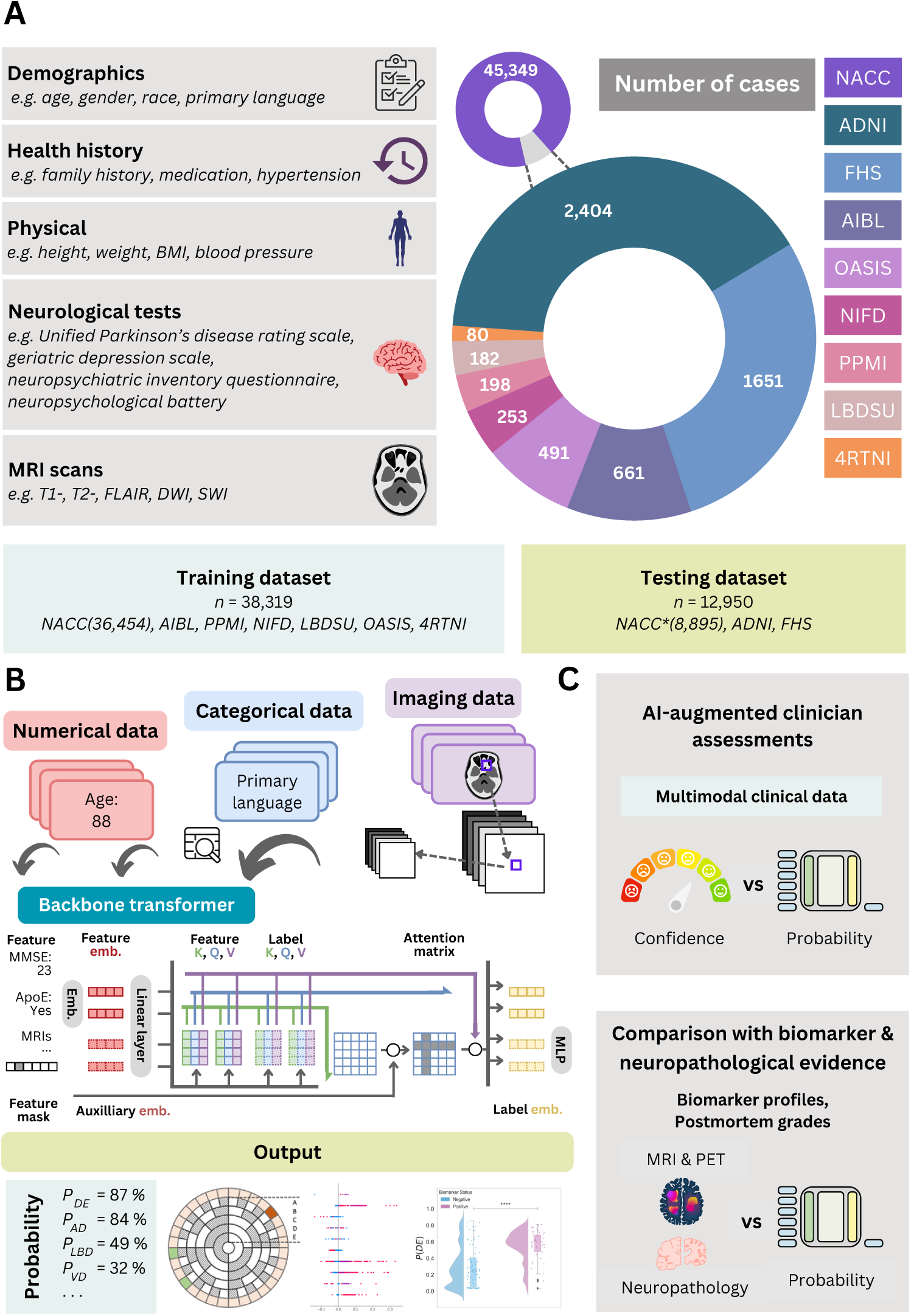
Data, model architecture and modeling strategy. (a) Our model for differential dementia diagnosis was developed using diverse data modalities, including individual-level demographics, health history, neurological testing, physical/neurological exams, and multi-sequence MRI scans. These data sources whenever available were aggregated from nine independent cohorts: 4RTNI, ADNI, AIBL, FHS, LBDSU, NACC, NIFD, OASIS, and PPMI (Tables 1 & S1). For model training, we merged data from NACC, AIBL, PPMI, NIFD, LBDSU, OASIS and 4RTNI. We employed a subset of the NACC dataset for internal testing. For external validation, we utilized the ADNI and FHS cohorts. (b) A transformer served as the scaffold for the model. Each feature was processed into a fixed-length vector using a modality-specific embedding strategy and fed into the transformer as input. A linear layer was used to connect the transformer with the output prediction layer. (c) A subset of the NACC dataset was randomly chosen to conduct a comparative analysis between neurologists’ performance augmented with the AI model and their performance without AI assistance. Similarly, we carried out comparative evaluations with practicing neuroradiologists, who were provided with a randomly selected sample of confirmed dementia cases from the NACC testing cohort, to assess the impact of AI augmentation on their diagnostic performance. For both these evaluations, the model and clinicians had access to the same set of multimodal data. Finally, we assessed the model’s predictions by comparing them with biomarker profiles and pathology grades available from the NACC, ADNI, and FHS cohorts.

### Model performance on NC, MCI and DE

We first sought to evaluate the performance of the model on test cases comprising individuals along the cognitive spectrum of NC, MCI and DE. The receiver operating characteristic (ROC) and precision-recall (PR) curves reflected strong model performance across different averaging methods (Figs. 2a & 2b). In the test set, comprising the NACC dataset unused in training, the Alzheimer’s Disease Neuroimaging Initiative (ADNI) and the Framingham Heart Study (FHS) data, our model demonstrated robust classification abilities for NC, MCI, and DE, achieving a micro-averaged AUROC of 0.94 and a micro-averaged AUPR of 0.90. Additionally, the macro-averaged metrics showed an AUROC of 0.93 and an AUPR value of 0.84. The weighted-average AUROC and AUPR values further demonstrated the model’s efficacy, standing at 0.94 and 0.87, respectively. Detailed model performance metrics across the three test cohorts are provided in Table S7. We also evaluated our model’s effectiveness by benchmarking it against a baseline machine learning algorithm, CatBoost,^41^ using identical case sets. This comparison was executed over two feature subsets, revealing that our model and CatBoost exhibited similar performances on the NACC dataset. Conversely, on the ADNI and FHS datasets, our model surpassed CatBoost, achieving higher AUROC and AUPR scores across all diagnostic categories with improvements ranging from 0.02 to 0.21 for AUROC and 0.03 to 0.17 for AUPR, as detailed in Table S8. This comparison highlights the improved generalizability of our model over traditional machine learning approaches in diagnostic tasks.

**Figure 2:**
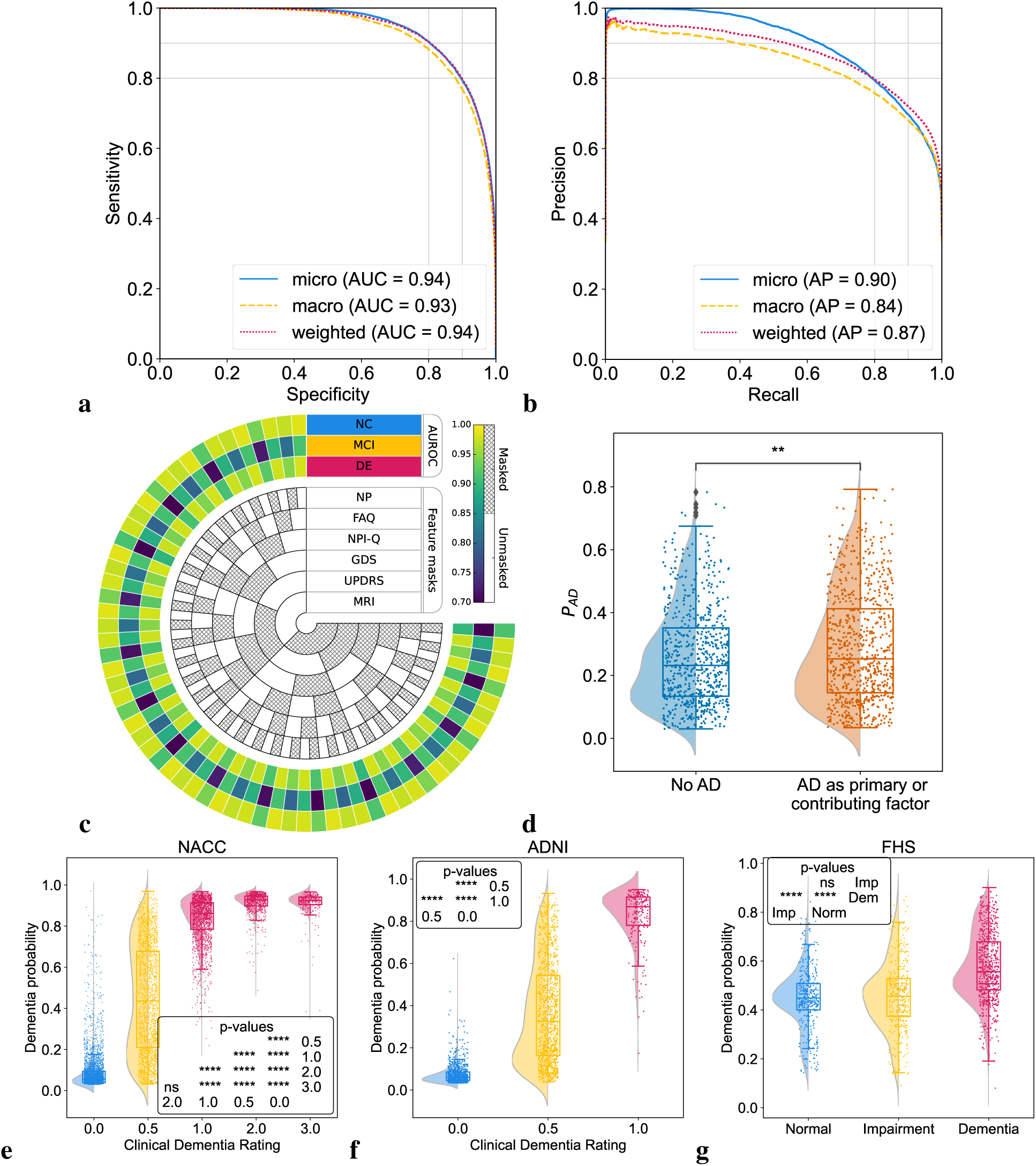
Model performance on individuals along the cognitive spectrum. (a,b) Receiver operating characteristic (ROC) and precision-recall (PR) curves, with their respective micro-average, macro-average, and weighted-average calculations based on the labels for NC, MCI, and DE. These averaging techniques consolidated the model’s performance across the spectrum of cognitive states. Cases from the NACC testing, ADNI and FHS were used. (c) Chord diagram indicating varied levels of model performance in the presence of missing data. The inner concentric circles represent various scenarios in which particular test information was either omitted (masked) or included (unmasked). The three outer concentric rings depict the model’s performance as measured by the area under the receiver operating characteristic curve (AUROC) for the NC, MCI and DE labels. (d, e, f) Raincloud plots with violin and box diagrams are shown to denote the distribution of clinical dementia rating scores (x-axis) versus model-predicted probability of dementia (y-axis), on the NACC, ADNI and FHS cohorts, respectively. (g) Raincloud plots are used to demonstrate the model’s ability to distinguish between MCI cases in the NACC cohort where AD was a factor for cognitive impairment and those attributed to non-AD etiologies. For plots (d-g), significance levels are denoted as ‘ns’ (not significant) for *p ≥* 0.05; * for *p <* 0.05; ** for *p <* 0.01; *** for *p <* 0.001; and **** for *p <* 0.0001 based on Kruskal-Wallis H-test for independent samples followed by post-hoc Dunn’s testing with Bonferroni correction.

### Model performance on incomplete data

To evaluate the model’s resilience to incomplete data, we artificially introduced varying levels of data missingness in the NACC cohort and assessed the impact on its predictive performance by selectively removing portions of the data to simulate different constraints. As depicted in the chord diagram (Fig. 2c), even when confronted with missing features, whether it be MRIs, UPDRS, GDS, NPI-Q, FAQ, NP tests or other parameters, our model consistently produced reliable scores. This reinforces not only its predictive stability, but also its potential applicability in various clinical scenarios where complete datasets are generally unattainable. Examples of this are found in our results on ADNI and FHS, which we used as external testing datasets (Tables S4 & S5). The ADNI cohort exhibited approximately 69% missing data compared to NACC, yet model predictions achieved a weighted-average AUROC of 0.91 and AUPR of 0.86 for NC, MCI, and DE categories. Similarly, with 94% fewer features than NACC, the model’s performance on FHS data also resulted in weighted-average AUROC and AUPR scores of 0.68 and 0.53 for NC, MCI, and DE categories, respectively.

### Model alignment with prodromal disease

We aimed to evaluate the model’s efficacy in identifying MCI individuals with an etiological diagnosis of AD, comparing the model’s predicted AD probabilities, *P* (*AD*), between MCI cases with AD as either a primary or contributing cause of their impairment and an etiological diagnosis of AD. Despite the fact that our model was trained on identifying AD dementia rather than prodromal AD, we found that it consistently assigned higher *P* (*AD*) to cases with MCI due to AD, compared to those with MCI due to other factors (Fig. 2d & Table S9). These results highlight our model’s clinical relevance in facilitating early disease detection and aiding clinicians in making informed decisions. Specifically, our findings support a therapeutic strategy of preemptive intervention in the AD continuum.

### Model alignment with clinical dementia ratings

We conducted a comparison between the model’s predicted DE probability scores, *P* (*DE*), and the clinical dementia ratings (CDR) available for all participants in the NACC testing, and ADNI cohorts (Figs. 2d & 2e, Table S10). Despite not incorporating CDR as input during model training, our predictions exhibited a strong correlation with CDR scores. In our analysis of the NACC dataset, we observed that *P* (*DE*) progressively increased with higher CDR scores, with statistically significant differences manifest across the spectrum of cognitive impairment (*p <* 0.0001). However, this pattern did not hold between CDR scores of 2.0 and 3.0, where no significant statistical difference was discerned. In the ADNI dataset, we found a statistically significant demarcation (*p <* 0.0001) in *P* (*DE*) between the baseline CDR rating and higher gradations. This points to the model’s sensitivity to incremental impairment in clinical dementia assessments. In the FHS dataset (Fig. 2f), which substitutes a consensus panel’s diagnostic categorization (normal, impaired, and dementia) for CDR scores, a marked statistical significance (*p <* 0.0001) was evident in *P* (*DE*) across these diagnostic strata, with the exception of normal versus impaired. This indicates a challenge for the model in distinguishing the early stages of cognitive decline when relying on a limited set of features. Such limitations are likely due to the community-based nature of the FHS cohort and the specificities of consensus panel ratings at FHS (Table S4). Collectively, these findings illuminate the model’s robust capacity to delineate differential cognitive states, showcasing its potential as a tool for identifying levels of cognitive impairment across datasets.

### Evaluation of single and co-occurring dementias

We evaluated our model’s diagnostic ability across ten distinct dementia etiologies. The ROC and PR curves in (Fig. 3a-b) reflect strong model performance on the model’s overall assessment on identifying dementia etiologies across different averaging methods, attaining micro-averaged AUROC and AUPR values of 0.96 and 0.70, respectively. In macro-averaged terms, the AUROC and AUPR stood at 0.91 and 0.36. Moreover, the weighted-average values for AUROC and AUPR were 0.94 and 0.73, respectively. The model’s performance, characterized by high micro-averaged and weighted-average AUROC and AUPR scores, underscores its diagnostic accuracy across a broad spectrum of dementia etiologies. While the lower macro-average AUPR scores indicate that our model may perform better on certain diagnoses relative to others, the weighted-average scores, adjusting for the prevalence of each dementia type, support the model’s effectiveness in a real-world clinical setting, where some dementia types are more common than others.

**Figure 3:**
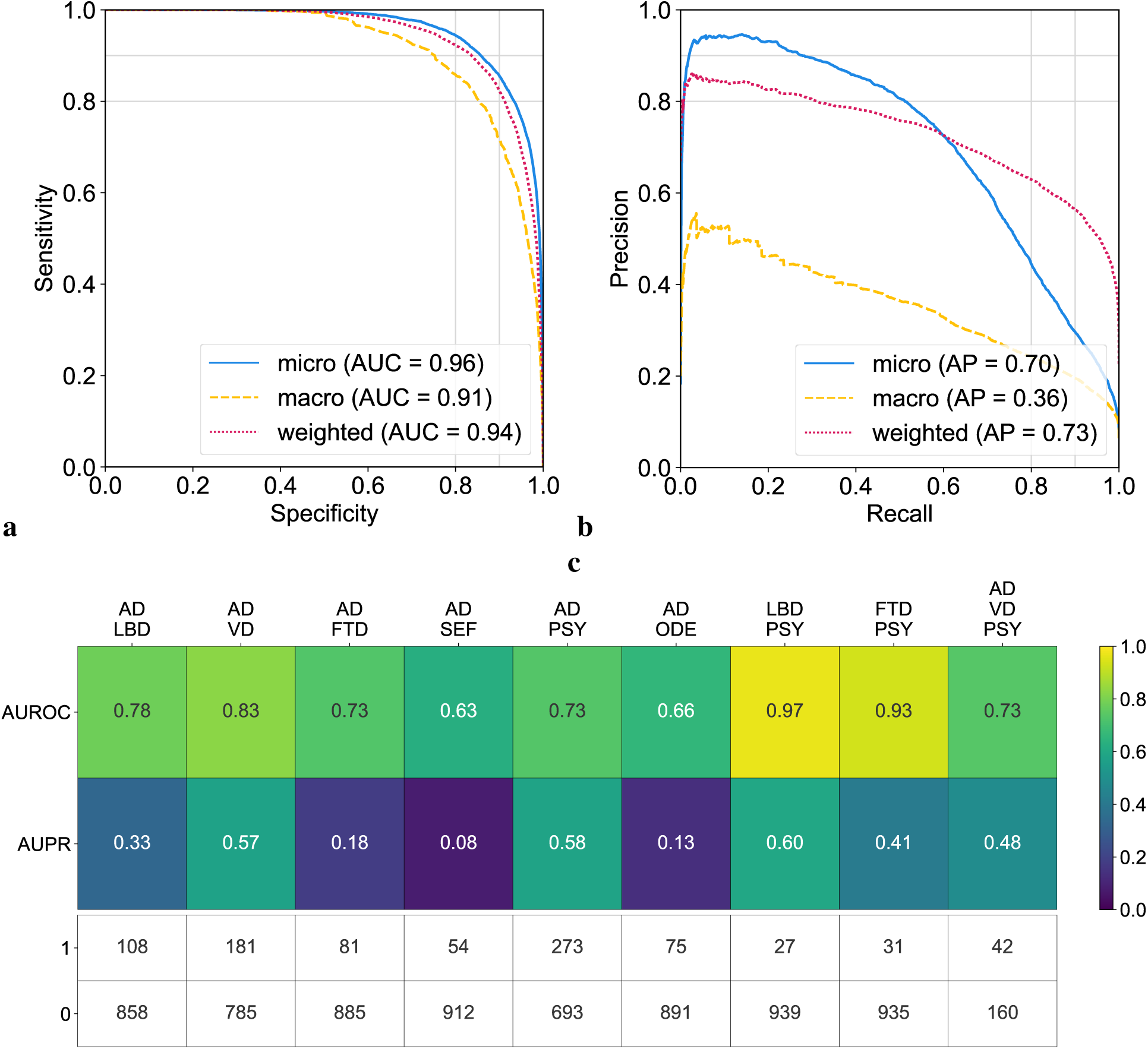
Model assessment on single and co-occurring dementias. (a, b) Receiver operating characteristic (ROC) and precision-recall (PR) curves are provided, utilizing micro-average, macro-average, and weighted-average methods across all the dementia diagnostic labels. These averages were computed to synthesize the performance metrics across all dementia etiologies. (c) Heatmaps are used to depict the model’s performance on co-occurring dementias. We considered all combinations where two or more etiologies co-occurred from the NACC testing cohort, provided there were at least 25 positive samples. This ensured that the maximum variance of the AUROC calculation over all possible continuous distributions was upper bounded by 0.01.^42^ The first row shows the AUROC values and the second row shows the AUPR values. The table also displays the sample sizes for each case, with 1 representing a positive case and 0 indicating a negative sample.

To further assess the model performance on co-occurring dementias, we adopted a maximum variance threshold of 0.01 for AUROC calculations.^42^ This selection aimed to balance the sensitivity and specificity of the model, enabling it to discern subtle diagnostic differences. This resulted in a minimum positive sample size of 25. In instances where two dementias co-occurred (Fig. 3c), the model’s AUROC scores varied from 0.63 to 0.97, reflecting a spectrum of diagnostic accuracy, with the LBD and PSY combination achieving the highest AUROC. AUPR scores ranged from 0.08 to 0.60, again with the conjunction of LBD and PSY recording the highest AUPR value. In the case of AD occurring with two other etiologies (VD & PSY), the AUROC score was 0.73 and the AUPR was 0.48. While our model demonstrated robust diagnostic discrimination, as evidenced by high AUROC values, the variability in AUPR scores may reflect challenges in consistently identifying less prevalent or more complex dementia etiologies within the dataset. Importantly, a similar pattern was found in subsequent analyses of expert neurologists’ performance for conditions such as SEF and TBI (Table S15). Additional performance metrics and visualizations that illustrate our model’s ability to assess single and co-occurring dementias are presented in the Supplement (Table S7 & Fig. S1).

### Model validation with biomarkers

Model-predicted probabilities for AD, FTD, and LBD were aligned with the presence of respective biomarkers, as demonstrated in the raincloud plots in Fig. 4 & Table S11. For AD, *P* (*AD*) correlated with A*β*, tau, and FDG PET biomarkers across the NACC and ADNI cohorts, indicating statistically significant differences between biomarker-negative and positive groups (*p <* 0.0001). Notably, *P* (*AD*) was consistently higher in A*β*, tau, and FDG PET positive groups, demonstrating that our framework’s diagnostic process aligns well with the current amyloid, tau, and neurodegeneration (ATN) criteria for AD diagnosis.^43^ Within the NACC cohort, FTD probabilities, *P* (*FTD*), were significantly associated with MRI and FDG PET biomarkers, with the biomarker positive groups having higher *P* (*FTD*). This result corroborates the capability of our model to detect FTD in alignment with observed patterns of fronto-temporal hypometabolism and atrophy.^44^ Finally, LBD probabilities, *P* (*LBD*), also displayed a clear differentiation when analyzed in relation to DaTscan evidence for LBD,^45^ with the DaTscan positive group exhibiting higher probabilities of LBD. Taken together, these findings validate the model’s effectiveness in capturing the pathophysiological underpinnings of prevalent dementia types in addition to the clinical syndrome, offering etiology-specific probability scores that closely match respective biomarker profiles. This alignment not only substantiates the model’s predictive validity, but also highlights its relevance to contemporary clinical practice as its mechanism for differential diagnosis of dementia reflects established biomarker criteria.

**Figure 4:**
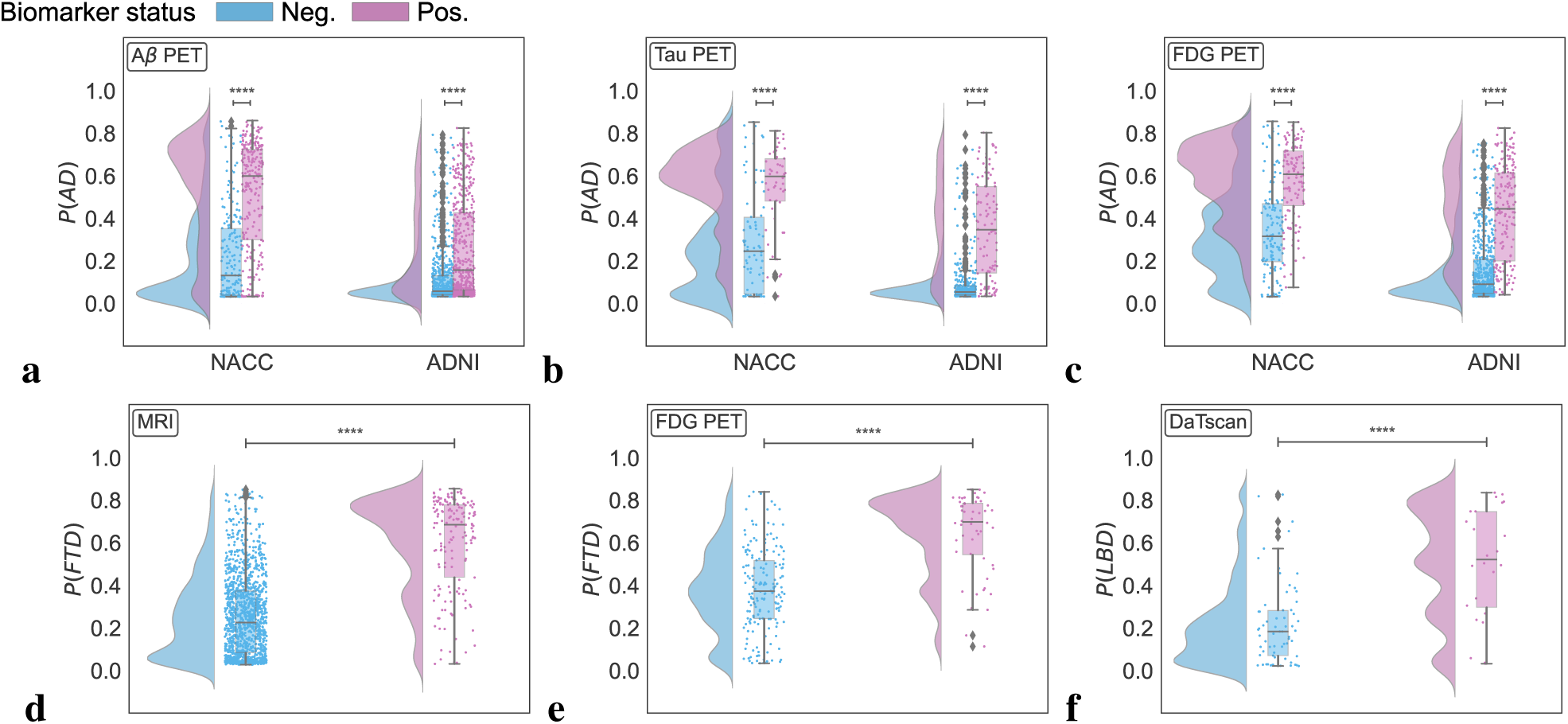
Biomarker-level validation. Raincloud plots representing model probabilities for dementia etiologies across their respective biomarker negative (blue) and positive groups (pink). (a-c) Model predicted probabilities for Alzheimer’s disease (*P* (*AD*)) were analyzed in relation to amyloid *β* (A*β*), tau, and fluorodeoxyglucose (FDG) PET biomarkers. Differences between A*β* negative and positive groups regarding *P* (*AD*) were evaluated using a one-sided Mann-Whitney U test for the NACC cohort and a one-sided t-test for ADNI. Similar analyses for tau and FDG PET biomarkers were conducted using one-sided Mann-Whitney U tests, with **** denoting *p <* 0.0001. (d-e) For frontotemporal lobar degeneration (*P* (*FT D*)), probabilities were assessed across MRI and FDG PET biomarker groups in the NACC cohort, using a one-sided Mann-Whitney U test, marked by **** for *p <* 0.0001. (f) Lewy body dementia (*P* (*LBD*)) probabilities were analyzed between DaTscan negative and positive groups using a one-sided Mann-Whitney U test, with **** indicating *p <* 0.0001.

### Model validation with neuropathological evidence

In cases with postmortem data (Table S12), we validated our model’s etiology-specific probability scores against neuropathological markers of common dementia types (Fig. 5 & Table S13). The composite violin and box plots indicate that, with increasing pathological severity, there is a corresponding elevation in the model-predicted likelihood of the etiology. The first three plots (Figs. 5a-c) compare AD probabilities against three key AD pathological markers with progressive stages: Thal phases of A*β* plaques, Braak stages of neurofibrillary degeneration, and Consortium to Establish a Registry for Alzheimer’s Disease (CERAD) density scores of neocortical neuritic plaques, denoted by A1-A3, B1-B3 and C1-C3, respectively. Each demonstrated an upward shift in the median probability of AD and an expansion of the interquartile range as the stages advanced, with statistical significance (*p <* 0.001 for Thal stage and *p <* 0.0001 for Braak and CERAD stages, respectively). We further evaluated our model’s predicted probabilities against cerebral amyloid angiopathy (CAA), a common pathological finding in AD confirmed postmortem cases. Similarly, we observed that our model predicted significantly higher AD probabilities in individuals with mild, moderate or severe CAA relative to those without CAA. Collectively, these plots illustrate a clear trend where advancing stages of AD-related pathology are associated with increased *P* (*AD*). Finally, significant differences were observed in *P* (*FTD*) and *P* (*V D*) based on their respective pathological markers: *P* (*FTD*) differed significantly between cases with and without TDP-43 pathology (*p <* 0.01) and tauopathy (*p <* 0.05), *P* (*V D*) varied between cases with and without old microinfarcts (*p <* 0.001) and arteriolosclerosis (*p <* 0.001) (Figs. 5e-h). The results are consistent with the well-documented association between TDP-43 protein aggregation and its prevalence in FTD.^46,47^ Additionally, the clear linkage between cerebrovascular pathologies and the incidence of VD is reinforced by our data. Crucially, these outcomes highlight the capability of our AI-driven framework to align model-generated probability scores with a range of neuropathological states beyond AD, supporting its potential utility in the evaluation of broader neurodegenerative diseases.

**Figure 5:**
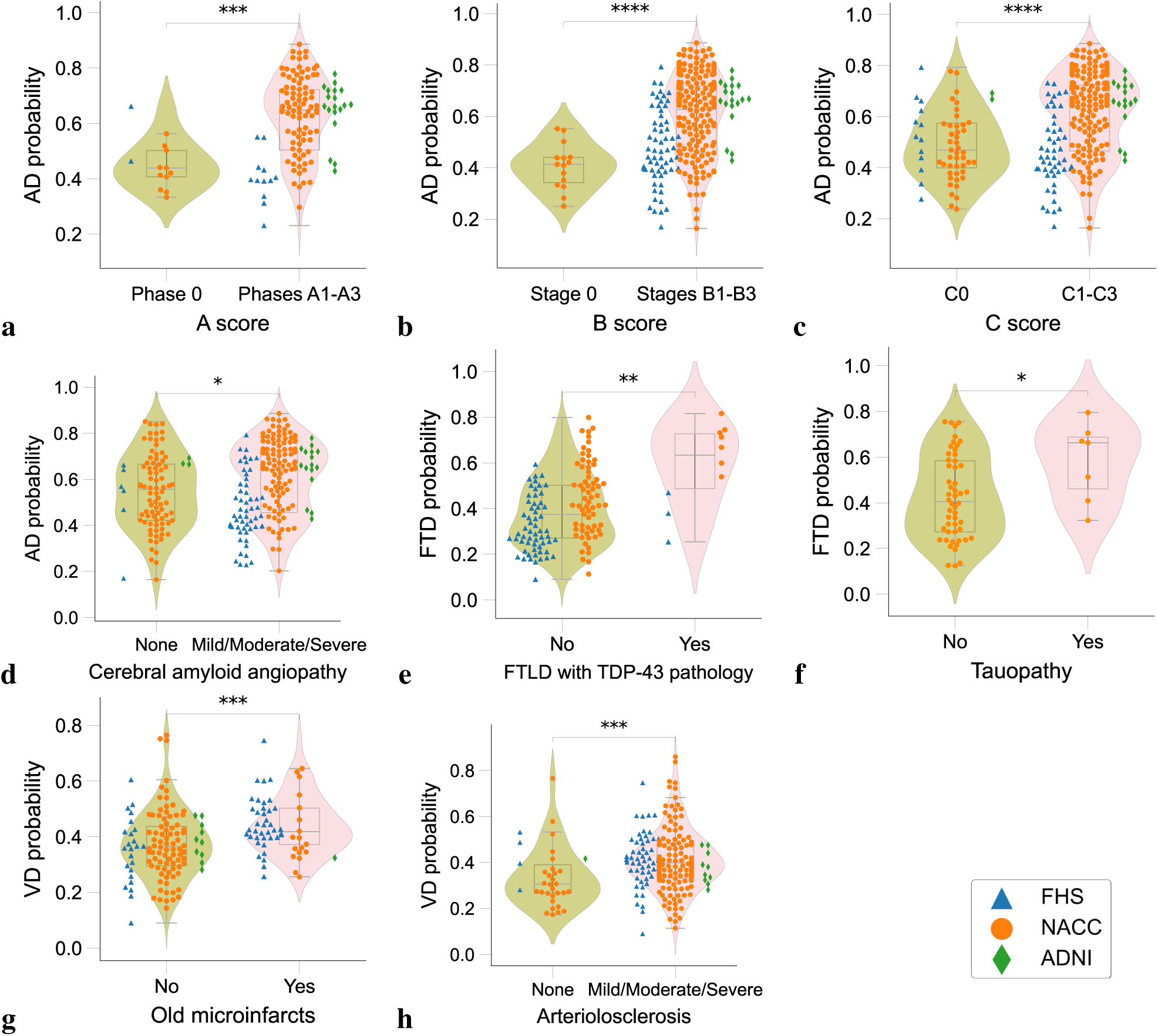
Neuropathological validation. Array of violin plots with integrated box plots, delineating the probability distributions as predicted by the model for different neuropathological grades. The analysis encompasses data from three distinct cohorts: the Framingham Heart Study (FHS), the National Alzheimer’s Coordinating Center (NACC), and the Alzheimer’s Disease Neuroimaging Initiative (ADNI), each denoted by unique markers (triangles, circles, and diamonds, respectively). Statistical significance is encoded using asterisks, determined by Dunn-Bonferroni post-hoc test: one asterisk (*) for *p <* 0.05; two asterisks (**) for *p <* 0.01, three asterisks (***) for *p <* 0.001, and four asterisks (****) for *p <* 0.0001, reflecting increasing levels of statistical significance. Table S13 presents more details on the statistics.

### AI-augmented clinician assessments

We aimed to assess whether our AI framework can compare to, and significantly enhance differential diagnosis of dementia performed by expert clinicians. To this end, we compared our model predicted probabilities with clinicians’ diagnoses, which were made in the form of confidence scores (0 to 100 scale). Neurologists reviewed 100 randomly selected cases, including various dementia subtypes, with comprehensive data including demographics, medical history, neuropsychological tests, and multi-sequence MRI scans. We observed that, in instances where the diagnosis was confirmed (true positives), the neurologists’ confidence scores across NC, MCI, DE, AD, LBD, VD, FTD, NPH, and PSY were higher in comparison to cases deemed non-diagnostic (true negatives) (*p <* 0.01) (Fig. S2a). In contrast, for the same 100 cases, our model’s predicted probabilities on true positive cases for all categories other than ODE were higher than the predicted probabilities for true negative cases (*p <* 0.01), indicating an enhanced ability for our model to detect true positives across more conditions (Fig. S2a). We then analyzed pairwise Pearson correlation coefficients to assess inter-rater agreement for each diagnostic category, both among neurologists’ confidence scores, and between the neurologists’ confidence scores and our model’s predicted probabilities (Fig. S3a). Among clinicians’ assessments, we found the most robust, consistent associations within the NC and DE groups, followed by modest associations between assessments of MCI, AD, LBD, VD, FTD and PSY. In contrast, PRD, NPH, SEF, TBI and ODE demonstrated the least consistency between neurologists’ assessments. This analysis shed light on dementia types that are relatively more challenging to diagnose, as evidenced by the variability in diagnostic confidence among expert clinicians. When comparing neurologists’ confidence scores with our model’s predicted probabilities, we found that the assessments provided by our model were generally consistent with those provided by the neurologists for NC, MCI, DE, AD, and LBD, as indicated by Pearson correlation coefficients that exceeded 0.7 (Fig. S3b). Associations were modest for VD, FTD, PSY, where mean Pearson correlation coefficients were approximately 0.5, while associations were less consistent for PRD, NPH, SEF, TBI, and ODE. The lower correlations observed here reflect the complex nature of these conditions, compounded by a lack of necessary features to tease out their unique signatures.

To determine whether our model could augment the assessments provided by neurologists, we computed AI-assisted neurologist confidence scores, which was defined as the mean of the neurologists’ confidence scores and our model’s predicted probabilities. We then compared the diagnostic performance of individual neurologist assessments with that of AI-augmented neurologist assessments (Figs. 6a-b & Tables S14 & S15). We consistently found significant increases in AUROC and AUPR for all etiologies (*p <* 0.05). There was a mean percent increase in AUROC of 26.25% and a mean percent increase in AUPR of 73.23% across all categories. The greatest improvement in diagnostic performance was for PRD and TBI, where there was a percent increase in mean AUROC of 73% and 72%, respectively, and a percent increase in mean AUPR of 242% and 257%, respectively. In a separate assessment, neuroradiologists evaluated a randomly selected set of 70 clinically diagnosed dementia cases, and were provided with multi-sequence MRIs, as well as demographic information. For these 70 cases, we assessed the diagnostic performance of radiologists and AI-augmented radiologists, which was defined as the mean of the radiologists’ confidence scores and our model’s probabilities (Figs. 6c-d & Tables S14 & S15). Across various dementia etiologies, we observed an average increase of 17.22% in AUROC and 42.17% in AUPR. A significant enhancement in AUROC (*p <* 0.05) was noted across all etiologies, with PRD showing the highest mean AUROC improvement at 68%. AUPR also displayed improvements, most markedly in PRD, where the mean AUPR surged by 190%.

**Figure 6:**
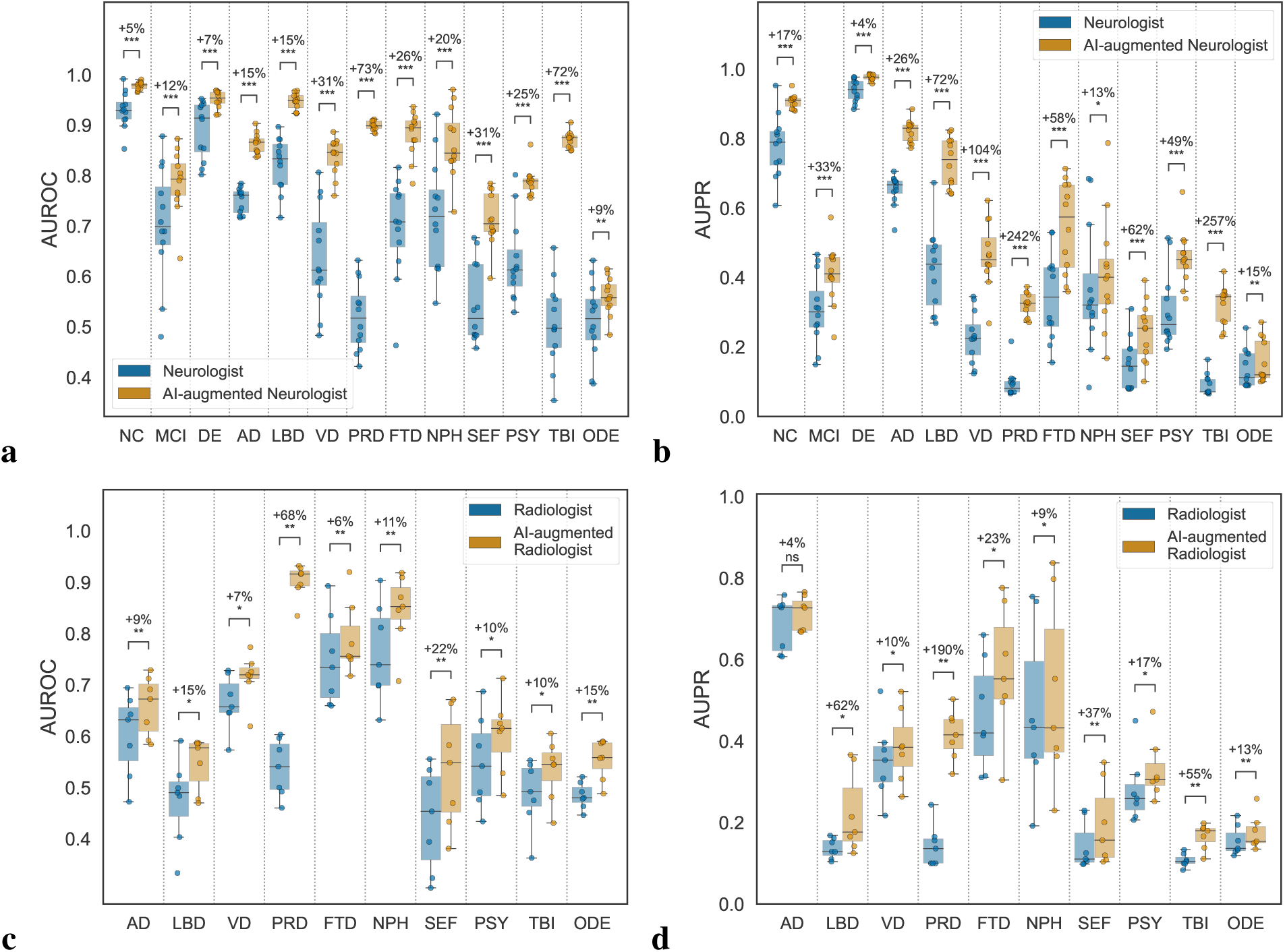
AI-augmented clinician assessments. Comparison between the performance of the assessments provided by practicing clinicians versus model-assisted clinicians is shown. (a-b) For the analysis, neurologists were given 100 randomly selected cases encompassing individual-level demographics, health history, neurological tests, physical as well as neurological examinations, and multi-sequence MRI scans. The neurologists were then tasked with assigning confidence scores for NC, MCI, DE, and the 10 dementia etiologies: AD, LBD, VD, PRD, FTD, NPH, SEF, PSY, TBI, and ODE (see Glossary 1). The boxplots show (a) AUROC and (b) AUPR for individual neurologist and model-assisted neurologist performance (defined as the mean between model and neurologist confidence scores). Pairwise statistical comparisons were conducted using the Wilcoxon signed-rank test and significance levels are denoted as: ns (not significant) for *p ≥* 0.05; * for *p <* 0.05; ** for *p <* 0.01; *** for *p <* 0.001; and **** for *p <* 0.0001. The percent increase in mean performance for each etiology is also presented above each statistical annotation. (c-d) Similarly, in a separate analysis, radiologists were given 70 randomly selected cases with a confirmed dementia diagnosis encompassing individual-level demographics and multi-sequence MRI scans. The radiologists were tasked with assigning confidence scores for the 10 dementia etiologies, and the boxplots show (c) AUROC and (d) AUPR for individual radiologist and model-assisted radiologist performance for the 10 etiologies. Statistical annotations and percent increase in mean performance with respect to each etiology are shown in a similar fashion.

## Discussion

We present an AI model designed for differential dementia diagnosis by processing a range of multimodal data. Unlike our previous work,^39,48^ our model addresses the clinical challenge of distinguishing between various dementia etiologies, including but not limited to AD, VD, and LBD. Such differentiation is crucial for the precise identification of the multi-factorial nature of dementia, which is linked to the optimization of personalized therapeutic interventions and patient management strategies. The model’s robustness was established through its training and validation across a diverse set of independent cohorts. Additionally, our model predictions on various etiologies were corroborated by their validation on a subset of cases for which biomarker and postmortem data were available. In a randomly selected subset of cases, our model’s predictions, when combined with neurologist assessments, outperformed the assessments conducted by neurologists alone. These results underscore our model’s potential in enhancing the efficacy of diagnosing dementia-related disorders.

Our model is designed to address the complex nature of mixed dementias by providing probability scores for each contributing etiology. This approach is significant as it enables clinicians to systematically prioritize possible drivers of cognitive impairment based on available data. The model effectively captures the multi-factorial and overlapping characteristics of various dementia types, offering a clear framework to guide clinical decision-making. For example, misdiagnoses in the initial stages of dementia are frequent, often due to symptom misattribution to psychiatric disorders, a situation further complicated by the presence of multiple co-pathologies.^49,50^ Specifically, LBD has historically been difficult to diagnose as early symptoms often resemble those of AD and PSY. The co-occurrence of LBD and AD further complicates diagnosis and tends to be missed entirely until post-mortem evaluation. ^51^ Our model demonstrated notable performance, particularly in identifying the AD and LBD combination, highlighting its capability in detecting mixed dementias that are commonly recognized only through postmortem analysis.^4,52,53^ This capability is crucial, given that a significant portion of dementia cases are linked to modifiable risk factors.^54^ The insights provided by our model could therefore inform early intervention strategies, potentially altering the disease course and enhancing patient outcomes. Notably, our model represents a significant step forward in the field, surpassing previous machine learning approaches in detecting mixed dementia, thereby offering a valuable tool for refining diagnostic accuracy in clinical practice.

Powered by a transformer architecture as the backbone, the utility of our modeling framework is founded on its robust processing of diverse input types and its adept handling of incomplete datasets. These properties are essential for clinicians requiring immediate and accurate diagnostic information in environments with variable data availability. For example, when a general practitioner records clinical observations and cognitive test results for an elderly person with possible cognitive decline, our model can calculate a probability score indicative of MCI or DE. This function facilitates early medical intervention and more informed decisions regarding specialist referrals. At a specialized memory clinic, the addition of extensive neuroimaging data and in-depth neuropsychological battery to the model may increase the precision of the diagnosis, which, in turn, enhances the formulation of individual management strategies with a revised probability score. Such capacity to tailor its output to the scope of input data exemplifies our modeling framework’s role in different healthcare settings, including those where swift and resource-efficient diagnosis is paramount. The generation of specific, quantifiable probability scores by the model augments its utility, establishing it as a useful component in the healthcare delivery process. Displaying diagnostic accuracy using varied training data — ranging from demographic information to clinical signs, neuroimaging findings, and neurological test results — the model’s versatility facilitates its adaptation to varied clinical operations without necessitating a fundamental overhaul of existing workflows. Consequently, our model fosters a seamless transition across the different levels of dementia care, enabling general practitioners to perform preliminary cognitive screenings and specialists to conduct thorough examinations. Its inclusive functionality assures an accessible and comprehensive tool ensuring fail-safe operation in early detection, continuous monitoring, and the fine-tuning of differential diagnoses, thereby elevating the standard of dementia care.

While our study has the potential to advance the field of differential dementia diagnosis, it does have certain limitations that warrant consideration. Our model was developed and validated on 9 distinct cohorts but its full generalizability across diverse populations and clinical settings remains to be determined. Moving forward, we see potential in evaluating the model’s efficacy across the care continuum, encompassing primary care facilities, geriatric and general neurology practices, family medicine, and specialized clinics in tertiary medical centers. Furthermore, AI models like ours possess the capability to enhance the patient screening procedures for clinical trial recruitment.^55^ Our study’s datasets primarily consist of AD cases, and while AD is the most common type of dementia, this could potentially skew our model towards improved recognition of this specific subtype, introducing a bias. Although we incorporated various dementia etiologies, the imbalanced representation might affect the model’s generalizability and sensitivity towards less frequent types. It is important to note that, beyond data imbalance, certain diseases were inherently more challenging to diagnose given the provided feature set, as exemplified by the lower performance metrics of expert neurologists in conditions such as SEF and TBI (Tables S14, S15). Additionally, we chose to amalgamate mild, moderate, and severe dementia cases into a single category. We acknowledge that this categorization method might not completely reflect the nuanced individual staging practiced in specific healthcare settings, where varying degrees of dementia severity carry distinct implications for treatment and management strategies. Our focus was primarily on differential diagnosis rather than disease staging, which motivated this decision. Future enhancements to our model could potentially include disease staging as an additional dimension, thereby augmenting its granularity and relevance. Finally, our study does not fully address the considerable heterogeneity inherent in AD, which is characterized by diverse clinical presentations and pathological features.^56,57^

The evidence collected from this study signals a convergence between advanced computational methods and the nuanced task of differential dementia diagnosis, crucial for scenarios with scarce resources and the multifaceted realm of mixed dementia, a condition frequently encountered yet diagnostically complex. Our model efficiently integrates multimodal data, showing strong performance across diverse settings. Future validations, encompassing a wider demographic and geographical expanse, will be pivotal to substantiate the model’s robustness and enhance its diagnostic utility in dementia care. Our pragmatic investigation accentuates the potential of neural networks to refine the granularity of diagnostic evaluations in neurocognitive disorders.

## Methods

### Study population

We collected demographics, personal and family history, laboratory results, findings from the physical/neurological exams, medications, neuropsychological tests, and functional assessments as well as multi-sequence magnetic resonance imaging (MRI) scans from 9 distinct cohorts, totaling 51, 269 participants. There were 19, 849 participants with normal cognition (NC), 9, 357 participants with mild cognitive impairment (MCI), and 22, 063 participants with dementia (DE). We further identified 10 primary and contributing causes of dementia: 17, 346 participants with Alzheimer’s disease (AD), 2, 003 participants with dementia with Lewy bodies and Parkinson’s disease dementia (LBD), 2, 032 participants with vascular brain injury or vascular dementia including stroke (VD), 114 participants with Prion disease including Creutzfeldt-Jakob disease (PRD), 3, 076 participants with frontotemporal lobar degeneration and its variants, which includes corticobasal degeneration (CBD) and progressive supranuclear palsy (PSP), and with or without amyotrophic lateral sclerosis (FTD), 138 participants with normal pressure hydrocephalus (NPH), 808 participants suffering from dementia due to infections, metabolic disorders, substance abuse including alcohol, medications, delirium and systemic disease - a category termed as systemic and external factors (SEF), 2, 700 participants suffering from psychiatric diseases including schizophrenia, depression, bipolar disorder, anxiety, and post-traumatic stress disorder (PSY), 265 participants with dementia due to traumatic brain injury (TBI), and 1, 234 participants with dementia due to other causes which include neoplasms, multiple systems atrophy, essential tremor, Huntington’s disease, Down syndrome, and seizures (ODE).

The cohorts include the National Alzheimer’s Coordinating Center (NACC) dataset (*n* = 45, 349),^58^ the Alzheimer’s Disease Neuroimaging Initiative (ADNI) dataset (*n* = 2, 404),^59^ the frontotemporal lobar degeneration neuroimaging initiative (NIFD) dataset (*n* = 253),^60^ the Parkinson’s Progression Marker Initiative (PPMI) dataset (*n* = 198),^61^ the Australian Imaging, Biomarker and Lifestyle Flagship Study of Ageing (AIBL) dataset (*n* = 661),^62^ the Open Access Series of Imaging Studies-3 (OASIS) dataset (*n* = 491),^63^ the 4 Repeat Tauopathy Neuroimaging Initiative (4RTNI) dataset (*n* = 80),^64^ and three in-house datasets maintained by the Lewy Body Dementia Center for Excellence at Stanford University (LBDSU) (*n* = 182),^65^ and the Framingham Heart Study (FHS) (*n* = 1, 651).^66^ Since its inception in 1948, FHS has been dedicated to identifying factors contributing to cardiovascular disease, monitoring multiple generations from Framingham, Massachusetts. Over time, the study has pinpointed major cardiovascular disease risk factors and explored their effects, while also investigating risk factors for conditions like dementia and analyzing the relationship between physical traits and genetics. Additional details on the study population are presented in Tables 1 & S1.

### Inclusion and exclusion criterion

Individuals from each cohort were eligible for study inclusion if they were diagnosed with normal cognition (NC), mild cognitive impairment (MCI), or dementia (DE). We used the National Alzheimer’s Coordinating Center (NACC) dataset,^58^ which is based on the Uniform Data Set (UDS) 3.0 dictionary,^67^ as the baseline for our study. To ensure data consistency, we organized the data from the other cohorts according to the UDS dictionary. For individuals from the NACC cohort who had multiple clinical visits, we initially prioritized the visits at which the person received the diagnostic label of dementia. We then selected the visit with the most data features available prioritizing the availability of neuroimaging information. If multiple visits met all the above criteria, we chose the most recent visit among them. This approach maximized the sample sizes of dementia cases, as well as ensured that each individual had the latest record included in the study while maximizing the utilization of available neuroimaging and non-imaging data. We included participants from the 4RTNI dataset^64^ with frontotemporal lobar degeneration (FTD)-related disorders like progressive supranuclear palsy (PSP) or corticobasal syndrome (CBS). For other cohorts (NIFD,^60^ PPMI,^61^ LBDSU,^65^ AIBL,^62^ ADNI,^59^ and OASIS^63^), participants were included if they had at least one MRI scan within 6 months of an officially documented diagnosis. From the FHS,^66^ we utilized data from the Original Cohort (Gen 1) enrolled in 1948, and the Offspring Cohort (Gen 2) enrolled in 1971. For these participants, we selected available data including demographics, history, clinical exam scores, neuropsychological test scores, and MRI within 6 months of the date of diagnosis. We did not exclude cases based on the absence of features (including imaging) or diagnostic labels. Instead, we employed our innovative model training approach to address missing features or labels (See below).

### Data processing and training strategy

Various non-imaging features (n=391) corresponding to subject demographics, medical history, laboratory results, medications, neuropsychological tests, and functional assessments were included in our study. We combined data from 4RTNI, AIBL, LBDSU, NACC, NIFD, OASIS, and PPMI to train the model. We used a portion of the NACC dataset for internal testing, while the ADNI and FHS cohorts served for external validation (Tables 1, S1–S5). We used a series of steps such as standardizing the data across all cohorts and formatting the features into numerical or categorical variables before using them for model training. We used stratified sampling at the person-level to create the training, validation, and testing splits. As we pooled the data from multiple cohorts, we encountered challenges related to missing features and labels. To address these issues and enhance the robustness of our model against data unavailability, we incorporated several strategies such as random feature masking and masking of missing labels (see below).

### MRI processing

Our investigation harnessed the potential of multi-sequence magnetic resonance imaging (MRI) volumetric scans sourced from diverse cohorts (Table S6). The majority of these scans encompassed T1-weighted (T1w), T2-weighted (T2w), diffusion-weighted (DWI), susceptibility-weighted (SWI), and fluid-attenuated inversion recovery (FLAIR) sequences. The collected imaging data were stored in the NIFTI file format, categorized by participant and the date of their visit. The MRI scans underwent a series of pre-processing steps involving skull stripping, linear registration to the MNI space, and intensity normalization. Skull stripping was performed using SynthStrip,^68^ a computational tool designed for extracting brain voxels from various image types. Then, the MRI scans were registered using FSL’s ‘flirt’ tool for linear registration of whole brain images,^69^ based on the MNI152 atlas.^70^ Prior to linear registration to the MNI space, we utilized the ‘fslorient2std’ function within FSL to standardize the orientation across all scans to match the MNI template’s axis order. As a result, the registered scans followed the dimensions of the MNI152 template, which are 182 *×* 218 *×* 182. Finally, all MRI scans underwent intensity normalization to the range [0,1] to increase the homogeneity of the data. To ensure the purity of the dataset, we excluded calibration, localizer, and 2D scans from the downloaded data before initiating model training.

### Backbone architecture

Our modeling framework harnesses the power of the transformer architecture to interpret and process a vast array of diagnostic parameters, including person-level demographics, medical history, neuroimaging, functional assessments, and neuropsychological test scores. Each of these distinct features is initially transformed into a fixed-length vector using a modality-specific strategy, forming the initial layer of input for the transformer model. Following this, the transformer acts to aggregate these vector inputs, decoding them into a series of predictions. A distinguishing strength of this framework lies in its integration of the transformer’s masking mechanism,^71,72^ strategically deployed to emulate missing features. This capability enhances the model’s robustness and predictive power, allowing it to adeptly handle real-world scenarios characterized by incomplete data.

### Multimodal data embeddings

Transformers use a uniform representation for all input tokens, typically in the form of fixed-length vectors. However, the inherent complexity of medical data, with its variety of modalities, poses a challenge to this requirement. Therefore, medical data needs to be adapted into a unified embedding that our transformer model can process. The data we accessed falls into three primary categories: numerical data, categorical data, and imaging data. Each category requires a specific method of embedding. Numerical data typically encompasses those data types where values are defined in an ordinal manner that holds distinct real-world implications. For instance, chronological age fits into this category, as it serves as an indicator of the aging process. To project numerical data into the input space of the transformer, we employed a single linear layer to ensure an appropriate preservation of the structure inherent to the original data space. Categorical data encompasses those inputs that can be divided into distinct categories yet lack any implicit order or priority. An example of this is gender, which can be categorized as ‘male’or ‘female’. We utilized a lookup table to translate categorical inputs into corresponding embeddings. It is noteworthy that this approach is akin to a linear transformation when the data is one-hot vectorized, but is computationally efficient, particularly when dealing with a vast number of categories. Imaging data, which includes MRI scans in medical applications, can be seen as a special case of numerical data. However, due to their high dimensionality and complexity, it is difficult to compress raw imaging data into a significantly lower-dimensionality vector using a linear transformation, while still retaining essential information. We leveraged the advanced capabilities of modern deep learning architectures to extract meaningful imaging embeddings (see below). Once these embeddings were generated, they were treated as numerical data, undergoing linear projection into vectors of suitable length, thus enabling their integration with other inputs to the transformer.

### Imaging feature extraction

We harnessed the Swin UNETR (Fig. S4),^73,74^ a three-dimensional (3D) transformer-based architecture, to extract embeddings from a multitude of brain MRI scans, encompassing various sequences including T1-weighted (T1w), T2-weighted (T2w), diffusion-weighted (DWI), susceptibility-weighted (SWI), and fluid-attenuated inversion recovery (FLAIR) imaging sequences. The Swin UNETR model consists of a Swin Transformer encoder, designed to operate on 3D patches, seamlessly connected to a convolutional neural network (CNN)-based decoder through multi-resolution skip connections. Commencing with an input volume *X ∈* R*^H×W^ ^×D^*, the encoder segmented *X* into a sequence of 3D tokens with dimensions 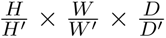, and projected them into a *C*-dimensional space via an embedding layer. It employed a patch size of 2 *×* 2 *×* 2 with a feature dimension of 2 *×* 2 *×* 2 *×* 1 and an embedding space dimension of *C* = 48. The Swin UNETR encoder was subsequently interconnected with a CNN-based decoder at various resolutions through skip connections, collectively forming a ‘U-shaped’ network. This decoder amalgamated the encoder’s outputs at different resolutions, conducted upsampling via deconvolutions, ultimately generating a reconstruction of the initial input volume. The pre-trained weights were the product of self-supervised pre-training of the Swin UNETR encoder, primarily conducted on 3D volumes encompassing the chest, abdomen, and head/neck.^73,74^

The process of obtaining imaging embeddings began with several transformations applied to the MRI scans. These transformations included resampling the scans to standardized pixel dimensions, foreground cropping, and spatial resizing, resulting in the creation of sub-volumes with dimensions of 128 *×* 128 *×* 128. Subsequently, these sub-volumes were input into the Swin UNETR model, which in turn extracted encoder outputs sized at 768 *×* 4 *×* 4 *×* 4. These extracted embeddings underwent downsampling via a learnable embedding module, consisting of four convolutional blocks, to align with the input token size of the downstream transformer. As a result, the MRI scans were effectively embedded into one-dimensional vectors, each of size 256. These vectors were then combined with non-imaging features and directed into the downstream transformer for further processing. The entire process utilized a dataset comprising 8, 155 MRI volumes, which were allocated for model training, validation, and testing (Table S6).

### Random feature masking

To enhance the robustness of the backbone transformer in handling data incompleteness, we leveraged the masking mechanism^71,72^ to emulate arbitrary missing features during training. The masking mechanism, when paired with the attention mechanism, effectively halts the information flow from a given set of input tokens, ensuring that certain features are concealed during prediction. A practical challenge arises when considering the potential combinations of input features, which increase exponentially. With hundreds of features in play, capturing every potential combination is intractable. Inspired by the definition of Shapley values, we deployed an efficient strategy for feature dropout. Given a sample with feature set *S*, *S* is randomly permuted as *σ*; simultaneously, an integer *i* is selected independently from the range [1*, |S|*]. Subsequent to this, the features *σ_i_*_+1_*, σ_i_*_+2_*,…, σ_|S|_* are masked out from the backbone transformer. It’s noteworthy that the dropout process was applied afresh across different training batches or epochs to ensure that the model gets exposed to a diverse array of missing information even within a single sample.

### Handling missing labels

The backbone transformer was trained by amalgamating data from multiple different cohorts, each focused on distinct etiologies, which introduced the challenge of missing labels in the dataset. While most conventional approaches involve discarding records with incomplete output labels during training, we chose a more inclusive strategy to maximize the utility of the available data. Our approach framed the task as a multi-label classification problem, introducing thirteen separate binary heads, one for each target label. With this design, for every training sample, we generated a binary mask indicating the absence of each label. We then masked the loss associated with samples lacking specific labels before backpropagation. This method ensured optimal utilization of the dataset, irrespective of label availability. The primary advantage of this approach lies in its adaptability. By implementing this label-masking strategy, our model can be evaluated against datasets with varying degrees of label availability, granting us the flexibility to address a wide spectrum of real-world scenarios.

### Loss function

Our backbone model was trained by minimizing the loss function (*L*) composed of two loss terms: “Focal Loss (FL)” ^75^ (*L*_FL_) and “Ranking Loss (RL)” (*L*_RL_), along with the standard L2 regularization term. FL is a variant of standard cross-entropy loss that addresses the issue of class imbalance. It assigns low weight to easy (well-classified) instances and employs a balance parameter. This loss function was used for each of the diagnostic categories (a total of 13, see Glossary 1). Therefore, our *L*_FL_ term was:

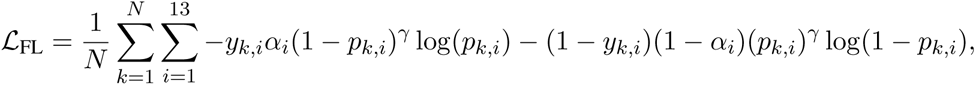

where *N* was the batch size (i.e., *N* = 128), and other parameters and variables were as defined. The focusing parameter *γ* was set to 2, which had been reported to work well in most of the experiments in the original paper.^75^ Moreover, *α_i_ ∈* [0, 1] was the balancing parameter that influenced the weights of positive and negative instances. It was set as the square of the complement of the fraction of samples labeled as 1, varying for each *i* due to the differing level of class imbalance across diagnostic categories (refer to Table 1). The FL term did not take inter-class relationships into account. To address these relationships in our overall loss function, we also incorporated the RL term that induced loss if the sigmoid outputs for diagnostic categories labeled as 0 were not lower than those labeled as 1 by a predefined margin of *ɛ*, for any training sample *k*. We defined the RL term for any pair of diagnostic categories *i* and *j*, as follows:

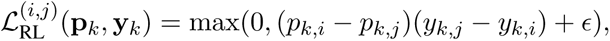

**Table 1:**
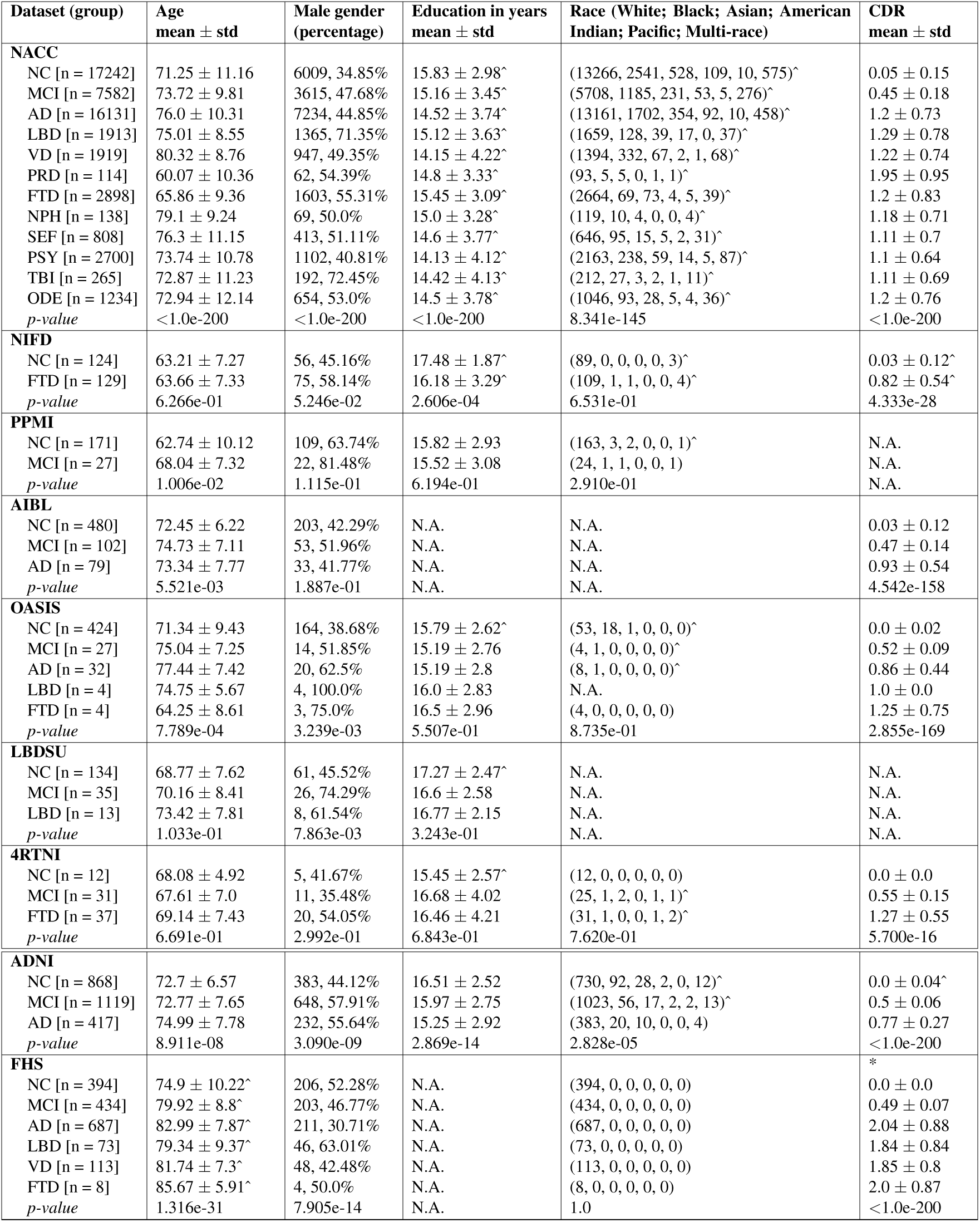
Study population. Nine independent datasets were used for this study, including ADNI, NACC, NIFD, PPMI, OASIS, LBDSU, 4RTNI, and FHS. Data from NACC, NIFD, PPMI, OASIS, LBDSU, and 4RTNI were used for model training. Data from ADNI, FHS, and a held-out set from NACC were used for model testing. The p-value for each dataset indicates the statistical significance of inter-group differences per column. We used one-way ANOVA and *χ*^2^ tests for continuous and categorical variables, respectively. Please refer to Glossary 1 for more information on the acronyms. Here N.A. denotes not available. The symbol ^ indicates that data was not available for some subjects. *∗* Due to the absence of CDR scores in the FHS dataset, we used the following definition: 0.0 - normal cognition, 0.5 - cognitive impairment, 1.0 - mild dementia, 2.0 - moderate dementia, 3.0 - severe dementia.

| Dataset (group) | Age<br>mean ± std | Male gender<br>(percentage) | Education in years<br>mean ± std | Race (White; Black; Asian; American<br>Indian; Pacific; Multi-race) | CDR<br>mean ± std |
| --- | --- | --- | --- | --- | --- |
| **NACC** |  |  |  |  |  |
| NC [n = 17242] | 71.25 ± 11.16 | 6009, 34.85% | 15.83 ± 2.98<sup>^</sup> | (13266, 2541, 528, 109, 10, 575)<sup>^</sup> | 0.05 ± 0.15 |
| MCI [n = 7582] | 73.72 ± 9.81 | 3615, 47.68% | 15.16 ± 3.45<sup>^</sup> | (5708, 1185, 231, 53, 5, 276)<sup>^</sup> | 0.45 ± 0.18 |
| AD [n = 16131] | 76.0 ± 10.31 | 7234, 44.85% | 14.52 ± 3.74<sup>^</sup> | (13161, 1702, 354, 92, 10, 458)<sup>^</sup> | 1.2 ± 0.73 |
| LBD [n = 1913] | 75.01 ± 8.55 | 1365, 71.35% | 15.12 ± 3.63<sup>^</sup> | (1659, 128, 39, 17, 0, 37)<sup>^</sup> | 1.29 ± 0.78 |
| VD [n = 1919] | 80.32 ± 8.76 | 947, 49.35% | 14.15 ± 4.22<sup>^</sup> | (1394, 332, 67, 2, 1, 68)<sup>^</sup> | 1.22 ± 0.74 |
| PRD [n = 114] | 60.07 ± 10.36 | 62, 54.39% | 14.8 ± 3.33<sup>^</sup> | (93, 5, 5, 0, 1, 1)<sup>^</sup> | 1.95 ± 0.95 |
| FTD [n = 2898] | 65.86 ± 9.36 | 1603, 55.31% | 15.45 ± 3.09<sup>^</sup> | (2664, 69, 73, 4, 5, 39)<sup>^</sup> | 1.2 ± 0.83 |
| NPH [n = 138] | 79.1 ± 9.24 | 69, 50.0% | 15.0 ± 3.28<sup>^</sup> | (119, 10, 4, 0, 0, 4)<sup>^</sup> | 1.18 ± 0.71 |
| SEF [n = 808] | 76.3 ± 11.15 | 413, 51.11% | 14.6 ± 3.77<sup>^</sup> | (646, 95, 15, 5, 2, 31)<sup>^</sup> | 1.11 ± 0.7 |
| PSY [n = 2700] | 73.74 ± 10.78 | 1102, 40.81% | 14.13 ± 4.12<sup>^</sup> | (2163, 238, 59, 14, 5, 87)<sup>^</sup> | 1.1 ± 0.64 |
| TBI [n = 265] | 72.87 ± 11.23 | 192, 72.45% | 14.42 ± 4.13<sup>^</sup> | (212, 27, 3, 2, 1, 11)<sup>^</sup> | 1.11 ± 0.69 |
| ODE [n = 1234] | 72.94 ± 12.14 | 654, 53.0% | 14.5 ± 3.78<sup>^</sup> | (1046, 93, 28, 5, 4, 36)<sup>^</sup> | 1.2 ± 0.76 |
| *p-value* | <1.0e-200 | <1.0e-200 | <1.0e-200 | 8.341e-145 | <1.0e-200 |
| **NIFD** |  |  |  |  |  |
| NC [n = 124] | 63.21 ± 7.27 | 56, 45.16% | 17.48 ± 1.87<sup>^</sup> | (89, 0, 0, 0, 0, 3)<sup>^</sup> | 0.03 ± 0.12<sup>^</sup> |
| FTD [n = 129] | 63.66 ± 7.33 | 75, 58.14% | 16.18 ± 3.29<sup>^</sup> | (109, 1, 1, 0, 0, 4)<sup>^</sup> | 0.82 ± 0.54<sup>^</sup> |
| *p-value* | 6.266e-01 | 5.246e-02 | 2.606e-04 | 6.531e-01 | 4.333e-28 |
| **PPMI** |  |  |  |  |  |
| NC [n = 171] | 62.74 ± 10.12 | 109, 63.74% | 15.82 ± 2.93 | (163, 3, 2, 0, 0, 1)<sup>^</sup> | N.A. |
| MCI [n = 27] | 68.04 ± 7.32 | 22, 81.48% | 15.52 ± 3.08 | (24, 1, 1, 0, 0, 1) | N.A. |
| *p-value* | 1.006e-02 | 1.115e-01 | 6.194e-01 | 2.910e-01 | N.A. |
| **AIBL** |  |  |  |  |  |
| NC [n = 480] | 72.45 ± 6.22 | 203, 42.29% | N.A. | N.A. | 0.03 ± 0.12 |
| MCI [n = 102] | 74.73 ± 7.11 | 53, 51.96% | N.A. | N.A. | 0.47 ± 0.14 |
| AD [n = 79] | 73.34 ± 7.77 | 33, 41.77% | N.A. | N.A. | 0.93 ± 0.54 |
| *p-value* | 5.521e-03 | 1.887e-01 | N.A. | N.A. | 4.542e-158 |
| **OASIS** |  |  |  |  |  |
| NC [n = 424] | 71.34 ± 9.43 | 164, 38.68% | 15.79 ± 2.62<sup>^</sup> | (53, 18, 1, 0, 0, 0)<sup>^</sup> | 0.0 ± 0.02 |
| MCI [n = 27] | 75.04 ± 7.25 | 14, 51.85% | 15.19 ± 2.76 | (4, 1, 0, 0, 0, 0)<sup>^</sup> | 0.52 ± 0.09 |
| AD [n = 32] | 77.44 ± 7.42 | 20, 62.5% | 15.19 ± 2.8 | (8, 1, 0, 0, 0, 0)<sup>^</sup> | 0.86 ± 0.44 |
| LBD [n = 4] | 74.75 ± 5.67 | 4, 100.0% | 16.0 ± 2.83 | N.A. | 1.0 ± 0.0 |
| FTD [n = 4] | 64.25 ± 8.61 | 3, 75.0% | 16.5 ± 2.96 | (4, 0, 0, 0, 0, 0) | 1.25 ± 0.75 |
| *p-value* | 7.789e-04 | 3.239e-03 | 5.507e-01 | 8.735e-01 | 2.855e-169 |
| **LBDSU** |  |  |  |  |  |
| NC [n = 134] | 68.77 ± 7.62 | 61, 45.52% | 17.27 ± 2.47<sup>^</sup> | N.A. | N.A. |
| MCI [n = 35] | 70.16 ± 8.41 | 26, 74.29% | 16.6 ± 2.58 | N.A. | N.A. |
| LBD [n = 13] | 73.42 ± 7.81 | 8, 61.54% | 16.77 ± 2.15 | N.A. | N.A. |
| *p-value* | 1.033e-01 | 7.863e-03 | 3.243e-01 | N.A. | N.A. |
| **4RTNI** |  |  |  |  |  |
| NC [n = 12] | 68.08 ± 4.92 | 5, 41.67% | 15.45 ± 2.57<sup>^</sup> | (12, 0, 0, 0, 0, 0) | 0.0 ± 0.0 |
| MCI [n = 31] | 67.61 ± 7.0 | 11, 35.48% | 16.68 ± 4.02 | (25, 1, 2, 0, 1, 1)<sup>^</sup> | 0.55 ± 0.15 |
| FTD [n = 37] | 69.14 ± 7.43 | 20, 54.05% | 16.46 ± 4.21 | (31, 1, 0, 0, 1, 2)<sup>^</sup> | 1.27 ± 0.55 |
| *p-value* | 6.691e-01 | 2.992e-01 | 6.843e-01 | 7.620e-01 | 5.700e-16 |
| **ADNI** |  |  |  |  |  |
| NC [n = 868] | 72.7 ± 6.57 | 383, 44.12% | 16.51 ± 2.52 | (730, 92, 28, 2, 0, 12)<sup>^</sup> | 0.0 ± 0.04<sup>^</sup> |
| MCI [n = 1119] | 72.77 ± 7.65 | 648, 57.91% | 15.97 ± 2.75 | (1023, 56, 17, 2, 2, 13)<sup>^</sup> | 0.5 ± 0.06 |
| AD [n = 417] | 74.99 ± 7.78 | 232, 55.64% | 15.25 ± 2.92 | (383, 20, 10, 0, 0, 4) | 0.77 ± 0.27 |
| *p-value* | 8.911e-08 | 3.090e-09 | 2.869e-14 | 2.828e-05 | <1.0e-200 |
| **FHS** |  |  |  |  |  |
| NC [n = 394] | 74.9 ± 10.22<sup>^</sup> | 206, 52.28% | N.A. | (394, 0, 0, 0, 0, 0) | *<br>0.0 ± 0.0 |
| MCI [n = 434] | 79.92 ± 8.8<sup>^</sup> | 203, 46.77% | N.A. | (434, 0, 0, 0, 0, 0) | 0.49 ± 0.07 |
| AD [n = 687] | 82.99 ± 7.87<sup>^</sup> | 211, 30.71% | N.A. | (687, 0, 0, 0, 0, 0) | 2.04 ± 0.88 |
| LBD [n = 73] | 79.34 ± 9.37<sup>^</sup> | 46, 63.01% | N.A. | (73, 0, 0, 0, 0, 0) | 1.84 ± 0.84 |
| VD [n = 113] | 81.74 ± 7.3<sup>^</sup> | 48, 42.48% | N.A. | (113, 0, 0, 0, 0, 0) | 1.85 ± 0.8 |
| FTD [n = 8] | 85.67 ± 5.91<sup>^</sup> | 4, 50.0% | N.A. | (8, 0, 0, 0, 0, 0) | 2.0 ± 0.87 |
| *p-value* | 1.316e-31 | 7.905e-14 | N.A. | 1.0 | <1.0e-200 |

Overall, the RL term was:

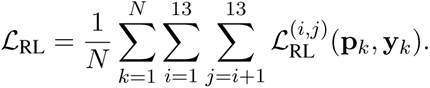

Combining all terms, our overall loss function (*L*) was:

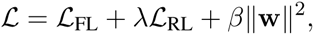

where *λ* and *β* were the weights that controlled the importance of *L*_RL_ and the L2 regularization terms, respectively. The training was done using the mini-batch strategy with the AdamW optimizer,^76^ an improved version of the Adam optimizer,^77^ with a learning rate of 0.001 for a total of 256 epochs. Additionally, we utilized a cosine learning rate scheduler with warm restarts,^78^ initiating the first restart after 64 epochs and extending the restart period by a factor of 2 for each subsequent restart. The values of *ɛ, λ,* and *β* were determined to be *ɛ* = 0.25, *λ* = 0.005, and *β* = 0.0005, respectively, based on an evaluation of the overall model performance on the validation set. During training, the model performance was evaluated on the validation set at the end of each epoch, and the model with the highest performance was selected.

### Traditional machine learning models

To assess our model’s ability to classify NC, MCI and DE cases, we compared its performance with the CatBoost model, a tree-based classification framework.^39,41^ Given the variability in available features across the test cohorts, we divided the data into two feature subsets, as detailed in Tables S2, S4 and S5. This stratification enabled a comparison with CatBoost, offering insights into our model’s performance using a range of parameters. The first feature subset consisted of variables common across all cohorts, including demographics, MMSE, and Boston Naming Test scores. The second subset expanded on this by incorporating additional neuropsychological measures found in the NACC and ADNI cohorts, such as trail making tests A and B, logical memory IIA delayed recall, MoCA scores, and digit span forward and backward tests. We trained separate CatBoost models for each feature set but applied our model to both subsets without retraining, allowing for a consistent evaluation across different feature configurations.

### Biomarker validation

The predicted probabilities of the model for various etiologies were cross-validated with established gold-standard biomarkers pertinent to each respective etiology. Both the NACC and ADNI test cohorts were used in AD biomarker analyses, while only NACC data were used for FTD and LBD due to biomarker availability. In the NACC dataset, binary UDS variables were used to define positivity for amyloid *β* (A*β*), tau and fluorodeoxyglucose F18 (FDG) PET biomarkers for AD due to varying PET processing methods across centers. Binary UDS variables were also used to define FDG and MRI evidence for FTD, and dopamine transporter scan (DATscan) as evidence for LBD. In ADNI, the University of California, Berkeley (UCB) A*β* PET processing pipeline yields Freesurfer-defined cortical summary and reference regions, as well as centiloids (CL). A cut-off value of 20 CL was chosen to define positivity.^79^ For tau, The UCB processing pipeline yields standardized uptake value ratios (SUVr) in Freesurfer-defined regions. A meta-temporal region of interest (ROI) was constructed following established standards.^80^ A Gaussian mixture model (GMM) with two components identified 1.74 SUVr as the optimal threshold to separate the two distributions, where values greater than 1.74 indicated tau PET positivity. Finally, the UCB FDG PET processing pipeline yields a meta-ROI, on which a GMM with two components identified 1.21 SUVr as the best threshold, where values less than 1.21 indicating positivity for neurodegeneration. Information regarding the PET processing protocols can be found in the summaries of UCB amyloid, tau, and FDG PET methods available on the LONI Image Data Archive website.^81^

### Neuropathologic validation

The model’s predictive capacity for various dementia etiologies was substantiated through alignment with neuropathological evaluations sourced from the NACC, FHS and ADNI cohorts (Table S12). We included participants who conformed to the study’s inclusion criteria, had undergone MRI scans no more than three years prior to death, and for whom neuropathological data were available. Standardization of data was conducted in accordance with the Neuropathology Data Form Version 10 protocols from the National Institute on Aging.^82^ We pinpointed neuropathological indicators that influence the pathological signature of each dementia etiology, such as arteriolosclerosis, the presence of neurofibrillary tangles and amyloid plaques, cerebral amyloid angiopathy (CAA), and markers of tauopathy. These indicators were carefully chosen to reflect the complex pathological terrain that defines each form of dementia. To examine the Thal phase for amyloid plaques (A score), subjects were categorized into two groups: one encompassing Phase 0, indicative of no amyloid plaque presence, and a composite group merging Phases 1-5, reflecting varying degrees of amyloid pathology. The model’s predictive performance was then compared across these groupings. For the Braak stage of neurofibrillary degeneration (B score), we consolidated stages I-VI into a single collective, representing the presence of AD-type neurofibrillary pathology, whereas stage 0 was designated for cases devoid of AD-type neurofibrillary degeneration. With respect to the density of neocortical neuritic plaques, assessed by the (CERAD or C score), individuals without neuritic plaques constituted one group, while those with any manifestation of neuritic plaques — sparse, moderate, or frequent (C1-C3) — were aggregated into a separate group for comparative analysis of the model’s predictive outcomes. To evaluate model alignment with the severity of CAA, subjects were classified into two groups: one representing the absence of CAA, and another encapsulating all stages of CAA severity, ranging from mild to severe. Furthermore, to evaluate the model’s concordance with non-AD pathologies, we analyzed the association between the model-generated probabilities of FTD with the presence of TDP-43 pathology and tauopathy, and VD with the presence of old microinfarcts and arteriolosclerosis.

### AI-augmented clinician assessments

We aimed to ascertain if our model could bolster the diagnostic prowess of clinicians specializing in dementia care and diagnosis. To this end, a group of 12 neurologists and 7 neuroradiologists were invited to participate in diagnostic tasks on a subset of NACC cases (see ‘Data processing and training strategy’). Neurologists were presented with 100 cases, which included 15 cases each of NC and MCI, and 7 cases for each of the dementia etiologies. The data encompassed person-level demographics, medical history, social history, neuropsychological tests, functional assessments, and multi-sequence MRI scans where possible (i.e., T1-weighted, T2-weighted, FLAIR, DWI and SWI sequences). They were asked to provide their diagnostic impressions, as well as a confidence score ranging from 0 to 100 for the diagnosis of each of the 13 labels. These confidence scores quantitatively reflect the clinician’s certainty in their diagnosis, with higher scores indicating greater certainty. This scoring system facilitated a quantitative comparison between the clinicians’ diagnostic certainty and the predictive probabilities generated by our model. Similarly, neuroradiologists were provided with the same multi-sequence MRI scans used by our model, along with information on age, gender, race, and education status from 70 clinically diagnosed DE cases. They were also tasked with providing diagnostic impressions, as well as confidence scores concerning the origin of dementia (Refer to Glossary 1). To evaluate the potential enhancement of clinical judgments by our model, we calculated AI-augmented confidence scores by averaging the clinicians’ confidence scores with our model’s predicted probabilities. We then assessed the diagnostic accuracy of the clinicians’ original and AI-augmented confidence scores using AUROC and AUPR metrics. The specifics of the case samples and questionnaires provided to the neurologists and neuroradiologists are detailed in the Supplementary Information.

### Statistical analysis

We used one-way ANOVA and the *χ*^2^ test for continuous and categorical variables, respectively to assess the overall differences in the population characteristics between the diagnostic groups across the study cohorts. We applied the Kruskal-Wallis H-test for independent samples and subsequently conducted post-hoc Dunn’s testing with Bonferroni correction to evaluate the relationship between clinical dementia rating (CDR) scores and the model-predicted probabilities, as well as between neuropathologic scores and the model-predicted probabilities. We used the two-sample Kolmogorov-Smirnov (K-S) test to compare model predicted AD probabilities, *P* (*AD*), between MCI cases with an etiological diagnosis of AD and MCI cases without one. We opted for non-parametric tests because the Shapiro-Wilk test indicated significant deviations from normality. In order to assess whether the model’s predicted probabilities for AD, FTD and LBD were significantly higher for their respective biomarker positive cases compared to biomarker negative ones, a one-sided Mann-Whitney U test was conducted when the Shapiro-Wilk test indicated significant deviations from normality. ADNI’s A*β* groups did not significantly deviate from normality and were therefore compared using the one-sided independent samples t-test. To compare model predictions with expert-driven assessments, we used the Brunner Munzel test to identify statistically significant increases in the mean disease probability scores between the levels of scoring categories. We conducted a Shapiro-Wilk test on the distributions of the true negative and true positive cases for each etiology. The Brunner-Munzel test was then used to compare the expert and model confidence scores for the true negative and true positive cases for each etiology. To evaluate the inter-rater reliability of label-specific confidence scores, we performed pairwise Pearson correlation analyses between clinicians’ scores and those generated by the model.^83^ We calculated the average correlation coefficient across pairs and determined its 95% confidence interval. In addition, we estimated the mean Pearson correlation coefficient between the confidence score of neurologists and the model’s score for each diagnostic label using a bootstrapping approach. Pairwise statistical comparisons of AI-augmented clinician diagnostic performance (AUROC and AUPR) and clinicians only diagnostic performance were performed with the one-sided Wilcoxon signed rank test. All statistical analyses were conducted at a significance level of 0.05.

### Performance metrics

We generated receiver operating characteristic (ROC) and precision-recall (PR) curves from predictions on both the NACC test data and other datasets. From each ROC and PR curve, we further derived the area under the curve values (AUC and AUPR, respectively). Further, we computed micro-, macro- and weighted-average AUC and AUPR values. Of note, the micro-average approach consolidates true positives, true negatives, false positives, and false negatives from all classes into a unified curve, providing a global performance metric. In contrast, the macro-average calculates individual ROC/PR curves for each class before computing their unweighted mean, disregarding potential class imbalances. The weighted-average, while similar in approach to macro-averaging, assigns a weight to each class’s ROC/PR curve proportionate to its representation in the dataset, thereby acknowledging class prevalence. We also evaluated the model’s accuracy, sensitivity, specificity, and Matthews correlation coefficient, with the latter being a balanced measure of quality for classes of varying sizes in a binary classifier.

### Computational hardware and software

All MRI and non-imaging data were processed on a workstation equipped with an Intel i9 14-core 3.3 GHz processor and 4 NVIDIA RTX 2080Ti GPUs. Our software development utilized Python (version 3.11.7) and the models were developed using PyTorch (version 2.1.0). We used several other Python libraries to support data analysis, including pandas (version 1.5.3), scipy (version 1.10.1), tensorboardX (version 2.6.2), torchvision (version 0.15), and scikit-learn (version 1.2.2). Training the model on a single Quadro RTX8000 GPU on a shared computing cluster had an average runtime of 7 minutes per epoch, while the inference task took less than a minute per instance. All clinicians reviewed MRIs using 3D Slicer (version 4.10.2) and logged their findings in REDCap (version 11.1.3).

### Data and code availability

Data from ADNI, AIBL, NACC, NIFD, OASIS, PPMI and 4RTNI can be downloaded from publicly available resources. Data from FHS and LBDSU can be obtained upon request, subject to institutional approval. Details on our model can be found on the Kolachalama Laboratory’s GitHub page (https://github.com/vkola-lab).

## Data Availability

All data produced in the present study are available upon reasonable request to the authors

## Acknowledgements

This project was supported by grants from the Karen Toffler Charitable Trust (VBK), National Institute on Aging’s Artificial Intelligence and Technology Collaboratories (P30-AG073014, VBK), the American Heart Association (20SFRN35460031, VBK & RA), Gates Ventures (RA & VBK), the Michael J. Fox Foundation (KLP), and the National Institutes of Health (R01-HL159620 [VBK], R21-CA253498 [VBK], R43-DK134273 [VBK], RF1-AG062109 [RA & VBK], U19-AG068753 [RA], P20-GM130447 [OT], K23-NS075097 [KLP], P50-AG047366 [KLP], and R01-NS115114 [KLP]). We acknowledge grant support from Boston University, CTSI 1UL1TR001430, for the REDCap Survey. We acknowledge the efforts of several individuals from the ADNI, AIBL, FHS, LBDSU, NACC, NIFD, OASIS, PPMI, and 4RTNI for providing access to data. Finally, we thank Drs. Shangran Qiu, Joyce C. Lee, Courtney E. Takahashi, Andrew M. Stern and Jesse B. Mez for several useful discussions.

The NACC database is funded by NIA grant U24-AG072122. The ADNI database is funded by NIA grant U01-AG024904. More details are outlined in the supplement.

## Contributions

C.X. and S.S.K. contributed equally to this work. S.S.K., D.L., S.P., V.H.J., O.T.Z., A.S.W., A.K., C.K., and T.F.A.A. performed data collection. C.X. and S.S.K. designed and developed the machine learning framework. C.X., S.S.K., D.L., S.P., V.H.J., O.B.G., and M.A. performed model training and validation. S.S.K., S.P., V.H.J., and M.A. performed statistical analysis. C.X., S.S.K., D.L., S.P., V.H.J., O.T.Z., A.S.W., O.B.G., J.D.Z., S.T.P. and M.A. generated the figures and tables. V.C.A.A., B.C.D., C.W.F., H.H., S.K., A.Z.M., D.L.M., S.O., A.B.P., S.R., M-H.S-H., E.A.S., B.N.S., J.E.S., A.S., O.T., J.Y., Y.Z. and S.Z. are practicing clinicians who reviewed the cases. S.A.B. and B.A.P. provided guidance on the modeling framework. K.L.P. and R.A. provided access to data. V.B.K. wrote the manuscript. All authors reviewed, edited and approved the manuscript. V.B.K. conceived, designed and directed the study.

## Ethics declarations

V.B.K. is on the scientific advisory board for Altoida Inc., and serves as a consultant to AstraZeneca. S.K. serves as consultant to AstraZeneca. C.W.F. is a consultant to Boston Imaging Core Lab. K.L.P. is a member of the scientific advisory boards for Curasen, Biohaven, and Neuron23, receiving consulting fees and stock options, and for Amprion, receiving stock options. R.A. is a scientific advisor to Signant Health and NovoNordisk. She also serves as a consultant to Davos Alzheimer’s Collaborative. The remaining authors declare no competing interests.

## Notes

### Competing Interest Statement

The authors have declared no competing interest.

### Funding Statement

This project was supported by various grants.

### Summary of Updates

Major changes in the text and results.

