## Supplement for "AI-based differential diagnosis of dementia etiologies on multimodal data"

| Dataset (group) | MMSE<br>mean ± std | MOCA<br>mean ± std | APOE<br>N, % positive |
|---|---|---|---|
| NACC | | | |
| NC [n = 17242] | 28.76 ± 1.57^ | 26.28 ± 2.7^ | 3852, 30.07%^ |
| MCI [n = 7582] | 26.94 ± 2.59^ | 22.61 ± 3.53^ | 1913, 38.2%^ |
| AD [n = 16131] | 18.94 ± 6.79^ | 15.15 ± 5.78^ | 6840, 56.05%^ |
| LBD [n = 1913] | 19.7 ± 7.07^ | 16.13 ± 5.95^ | 600, 44.02%^ |
| VD [n = 1919] | 19.56 ± 6.85^ | 16.19 ± 5.6^ | 577, 42.06%^ |
| PRD [n = 114] | 13.37 ± 10.03^ | 16.18 ± 7.6^ | 6, 23.08%^ |
| FTD [n = 2898] | 19.09 ± 8.13^ | 17.06 ± 6.66^ | 740, 31.97%^ |
| NPH [n = 138] | 20.54 ± 6.46^ | 17.38 ± 5.1^ | 38, 42.7%^ |
| SEF [n = 808] | 20.63 ± 6.33^ | 17.21 ± 5.46^ | 225, 39.4%^ |
| PSY [n = 2700] | 20.33 ± 6.52^ | 16.15 ± 5.83^ | 911, 46.13%^ |
| TBI [n = 265] | 21.07 ± 6.29^ | 17.24 ± 6.82^ | 74, 39.15%^ |
| ODE [n = 1234] | 20.32 ± 6.99^ | 17.11 ± 5.89^ | 403, 42.38%^ |
| *p-value* | <1.0e-200 | <1.0e-200 | <1.0e-200 |
| NIFD | | | |
| NC [n = 124] | 29.35 ± 0.76 | 27.58 ± 1.53^ | N.A. |
| FTD [n = 129] | 24.75 ± 4.54^ | 19.69 ± 5.72^ | N.A. |
| *p-value* | 1.961e-23 | 2.645e-16 | N.A. |
| PPMI | | | |
| NC [n = 171] | N.A. | 27.51 ± 2.37^ | N.A. |
| MCI [n = 27] | N.A. | 24.69 ± 3.27^ | N.A. |
| *p-value* | N.A. | 3.004e-07 | N.A. |
| AIBL | | | |
| NC [n = 480] | 28.7 ± 1.24 | N.A. | 12, 2.5% |
| MCI [n = 102] | 27.1 ± 2.08 | N.A. | 12, 11.76% |
| AD [n = 79] | 20.42 ± 5.46 | N.A. | 14, 17.72% |
| *p-value* | 4.585e-121 | N.A. | 8.951e-09 |
| OASIS | | | |
| NC [n = 424] | 28.99 ± 1.25^ | N.A. | 140, 33.02% |
| MCI [n = 27] | 28.15 ± 1.67 | N.A. | 11, 40.74% |
| AD [n = 32] | 23.91 ± 5.05 | N.A. | 20, 62.5% |
| LBD [n = 4] | 25.5 ± 2.69 | N.A. | 2, 50.0% |
| FTD [n = 4] | 18.33 ± 8.26^ | N.A. | 4, 100.0% |
| *p-value* | 4.439e-50 | N.A. | 4.510e-05 |
| LBDSU | | | |
| NC [n = 134] | N.A. | 27.43 ± 2.23^ | N.A. |
| MCI [n = 35] | N.A. | 24.0 ± 3.14 | N.A. |
| LBD [n = 13] | N.A. | 16.69 ± 4.75 | N.A. |
| *p-value* | N.A. | 2.231e-30 | N.A. |
| 4RTNI | | | |
| NC [n = 12] | 27.2 ± 2.4^ | 24.2 ± 2.44^ | N.A. |
| MCI [n = 31] | 26.1 ± 3.95^ | 21.19 ± 4.83^ | N.A. |
| FTD [n = 37] | 21.49 ± 7.2^ | 17.14 ± 7.4^ | N.A. |
| *p-value* | 1.657e-03 | 3.605e-03 | N.A. |
| ADNI | | | |
| NC [n = 868] | 29.09 ± 1.12 | 25.97 ± 2.65^ | 138, 29.61%^ |
| MCI [n = 1119] | 27.64 ± 1.85 | 23.07 ± 3.25^ | 438, 47.2%^ |
| AD [n = 417] | 23.19 ± 2.23 | 16.93 ± 4.53^ | 229, 64.33%^ |
| *p-value* | <1.0e-200 | 3.984e-199 | 3.117e-22 |
| FHS | | | |
| NC [n = 394] | 26.05 ± 3.36^ | N.A. | N.A. |
| MCI [n = 434] | 25.2 ± 3.4^ | N.A. | N.A. |
| AD [n = 687] | 21.63 ± 5.08^ | N.A. | N.A. |
| LBD [n = 73] | 22.91 ± 4.16^ | N.A. | N.A. |
| VD [n = 113] | 22.41 ± 5.44^ | N.A. | N.A. |
| FTD [n = 8] | 20.33 ± 3.4^ | N.A. | N.A. |
| *p-value* | 3.132e-26 | N.A. | N.A. |

Table S1: **Study population.** Nine independent datasets were used for this study, including ADNI, NACC, NIFD, PPMI, OASIS, LBDSU, 4RTNI, and FHS. Data from NACC, NIFD, PPMI, OASIS, LBDSU, and 4RTNI were used for model training. Data from ADNI, FHS, and a held-out set from NACC were used for model testing. The p-value for each dataset indicates the statistical significance of inter-group differences per column. We used one-way ANOVA and $\chi^2$ tests for continuous and categorical variables, respectively. Please refer to Glossary 1 for more information on the acronyms. Here N.A. denotes not available. The symbol ^ indicates that data was not available for some subjects.

| Cohort | Features |
| --- | --- |
| NACC | • Primary reason for visit<br>• Principal referral source<br>• subject's month of birth<br>• subject's year of birth<br>• Hispanic/Latino ethnicity<br>• Hispanic origins<br>• Race<br>• Second race<br>• Third race<br>• Primary language<br>• Years of education<br>• Marital status<br>• Living situation<br>• Level of independence<br>• Type of residence<br>• Is the subject left- or right-handed?<br>• Subject's age at visit<br>• Derived NIH race definitions<br>• Indicator of first-degree family member with cognitive impairment<br>• Indicator of mother with cognitive impairment<br>• Indicator of father with cognitive impairment<br>• In this family, is there evidence of a dominantly inherited AD mutation?<br>• In this family, is there evidence for an AD mutation (from a list of specific mutations)?<br>• Source of evidence for AD mutation<br>• In this family, is there evidence for an FTLD mutation?<br>• In this family, is there evidence for an FTLD mutation (from a list of specific mutations)?<br>• Source of evidence for FTLD mutation<br>• In this family, is there evidence for a mutation other than an AD or FTLD mutation?<br>• Source of evidence for other mutation<br>• Smoked cigarettes in last 30 days<br>• Smoked more than 100 cigarettes in life<br>• Total years smoked cigarettes<br>• Average number of packs smoked per day<br>• If the subject quit smoking, the age at which he/she last smoked (i.e., quit)<br>• In the past three months, has the subject consumed any alcohol?<br>• During the past three months, how often did the subject have at least one drink of any alcoholic beverage such as wine, beer, malt liquor, or spirits?<br>• Heart attack/cardiac arrest<br>• More than one heart attack/cardiac arrest?<br>• Year of most recent heart attack<br>• Atrial fibrillation<br>• Angioplasty/endarterectomy/stent<br>• Cardiac bypass procedure<br>• Pacemaker and/or defibrillator<br>• Pacemaker<br>• Congestive heart failure<br>• Angina<br>• Heart valve replacement or repair<br>• Other cardiovascular disease<br>• Stroke<br>• More than one stroke reported as of the Initial Visit<br>• Most recently reported year of stroke as of the Initial Visit<br>• Transient ischemic attack (TIA)<br>• More than one TIA reported as of the Initial Visit<br>• Most recently reported year of TIA as of the Initial Visit<br>• Parkinson's disease (PD)<br>• Year of PD diagnosis<br>• Other Parkinsonian disorder<br>• Year of Parkinsonian disorder diagnosis<br>• Seizures<br>• Traumatic brain injury (TBI)<br>• Traumatic brain injury (TBI) with brief loss of consciousness<br>• brain trauma - brief unconsciousness<br>• TBI with extended loss of consciousness - 5 minutes or longer<br>• brain trauma - extended unconsciousness<br>• TBI without loss of consciousness - as might result from military detonations or sports injury<br>• brain trauma - chronic deficit<br>• Year of most recent TBI<br>• Other neurological condition<br>• Diabetes<br>• If Recent/active or Remote/inactive diabetes, which type?<br>• Hypertension<br>• Hypercholesterolemia<br>• Vitamin B12 deficiency<br>• Thyroid disease<br>• Arthritis<br>• Type of Arthritis<br>• Arthritis, region affected - spine<br>• Region affected - unknown<br>• Incontinence - urinary<br>• Incontinence - bowel<br>• Sleep apnea history reported at Initial Visit<br>• REM sleep behavior disorder (RBD) history reported at Initial Visit<br>• Hyposomnia/insomnia history reported at Initial Visit<br>• Other sleep disorder history reported at Initial Visit<br>• Alcohol abuse - clinically significant occurring over a 12-month period manifested in one of the following areas: work, driving, legal, or social<br>• Other abused substances - clinically significant impairment occurring over a 12-month period manifested in one of the following areas: work, driving, legal, or social<br>• Post-traumatic stress disorder (PTSD)<br>• bipolar disorder<br>• Schizophrenia<br>• Active depression in the last two years<br>• Depression episodes more than two years ago<br>• Anxiety<br>• Obsessive-compulsive disorder (OCD)<br>• Developmental neuropsychiatric disorders (e.g., autism spectrum disorder [ASD], attention-deficit hyperactivity disorder [ADHD], dyslexia)<br>• Other psychiatric disorder<br>• History of traumatic brain injury (TBI)<br>• Subject's sex<br>• Arthritis, region affected - upper extremity<br>• Arthritis, region affected - lower extremity<br>• NPI-Q co-participant<br>• Delusions severity<br>• Hallucinations severity<br>• Agitation or aggression severity<br>• Depression or dysphoria severity |

Table S2

| Cohort | Features |
|---|---|
| NACC | • Elation or euphoria severity<br>• Apathy or indifference severity<br>• Disinhibition severity<br>• Irritability or lability severity<br>• Motor disturbance severity<br>• Nighttime behaviors severity<br>• Appetite and eating severity<br>• Anxiety severity<br>• Number of APOE e4 alleles<br>• Were there abnormal neurological exam findings?<br>• Are focal deficits present indicative of central nervous system disorder?<br>• Is gait disorder present indicative of central nervous system disorder?<br>• Are there eye movement abnormalities present indicative of central nervous system disorder?<br>• Parkinsonian signs<br>• Resting tremor - left arm<br>• Resting tremor - right arm<br>• Slowing of fine motor movements - left side<br>• Slowing of fine motor movements - right side<br>• Rigidity - left arm<br>• Rigidity - right arm<br>• bradykinesia<br>• Parkinsonian gait disorder<br>• Postural instability<br>• Neurological sign considered by the examiner to be most likely consistent with cerebrovascular disease<br>• Cortical cognitive deficit (e.g., aphasia, apraxia, neglect)<br>• Focal or other neurological findings consistent with SIVD (subcortical ischemic vascular dementia)<br>• Motor (may include weakness of combination of face, arm, and leg; reflex changes, etc.) - left side<br>• Motor (may include weakness of combination of face, arm, and leg; reflex changes, etc.) - right side<br>• Cortical visual field loss - left side<br>• Cortical visual field loss - right side<br>• Somatosensory loss - left side<br>• Somatosensory loss - right side<br>• Higher cortical visual problem suggesting posterior cortical atrophy (e.g., prosopagnosia, simultagnosia, Balint's syndrome) or apraxia of gaze<br>• Findings suggestive of progressive supranuclear palsy (PSP), corticobasal syndrome (CBS), or other related disorders<br>• Eye movement changes consistent with PSP<br>• Dysarthria consistent with PSP<br>• Axial rigidity consistent with PSP<br>• Gait disorder consistent with PSP<br>• Apraxia of speech<br>• Apraxia consistent with CBS - left side<br>• Apraxia consistent with CBS - right side<br>• Cortical sensory deficits consistent with CBS - left side<br>• Cortical sensory deficits consistent with CBS - right side<br>• Ataxia consistent with CBS - left side<br>• Ataxia consistent with CBS - right side<br>• Alien limb consistent with CBS - left side<br>• Alien limb consistent with CBS - right side<br>• Dystonia consistent with CBS, PSP, or related disorder - left side<br>• Dystonia consistent with CBS, PSP, or related disorder - right side<br>• Myoclonus consistent with CBS - left side<br>• Myoclonus consistent with CBS - right side<br>• Findings suggesting ALS (e.g., muscle wasting, fasciculations, upper motor and/or lower motor neuron signs)<br>• Normal pressure hydrocephalus - gait apraxia<br>• Other findings (e.g., cerebella ataxia, chorea, myoclonus)<br>• Were all findings unremarkable?<br>• Was any part of the MMSE completed?<br>• Administration of the MMSE was:<br>• Language of MMSE administration<br>• Subject was unable to complete one or more sections due to visual impairment<br>• Subject was unable to complete one or more sections due to hearing impairment<br>• Orientation subscale score - Time<br>• Orientation subscale score - Place<br>• Intersecting pentagon subscale score<br>• Total MMSE score (using D-L-R-O-W)<br>• The remainder of the battery was administered:<br>• Language of test administration<br>• If this test has been administered to the subject within the past 3 months, specify the date previously administered (month)<br>• If this test has been administered to the subject within the past 3 months, specify the date previously administered (day)<br>• If this test has been administered to the subject within the past 3 months, specify the date previously administered (year)<br>• Total score from the previous test administration<br>• Total number of story units recalled from this current test administration<br>• Logical Memory IIA - Delayed - Total number of story units recalled<br>• Logical Memory IIA - Delayed - Time elapsed since Logical Memory IA - Immediate<br>• Total score for copy of Benson figure<br>• Total score for 10- to 15-minute delayed drawing of Benson figure<br>• Recognized original stimulus from among four options<br>• Digit span forward trials correct<br>• Digit span forward length<br>• Digit span backward trials correct<br>• Digit span backward length<br>• Animals - Total number of animals named in 60 seconds<br>• Vegetable - Total number of vegetables named in 60 seconds<br>• Trail Making Test Part A - Total number of seconds to complete<br>• Part A - Number of commission errors<br>• Part A - Number of correct lines<br>• Trail Making Test Part B - Total number of seconds to complete<br>• Part B - Number of commission errors<br>• Part B - Number of correct lines<br>• WAIS-R Digit Symbol<br>• Boston Naming Test (30) - Total score<br>• Number of correct F-words generated in 1 minute<br>• Number of F-words repeated in 1 minute<br>• Number of non-F-words and rule violation errors in 1 minute<br>• Number of correct L-words generated in 1 minute<br>• Number of L-words repeated in 1 minute<br>• Number of non-L-words and rule violation errors in 1 minute<br>• Total number of correct F-words and L-words<br>• Total number of F-word and L-word repetition errors<br>• Total number of non-F/L-words and rule violation errors |

Table S2

| Cohort | Features |
|---|---|
| NACC | • Per clinician, based on the neuropsychological examination, the subject's cognitive status is deemed<br>• Modality of communication used to administer neuropsychological battery<br>• Was any part of MoCA administered?<br>• If no part of MoCA administered, reason code<br>• Where was MoCA administered?<br>• Language of MoCA administration<br>• Subject was unable to complete one or more sections due to visual impairment<br>• Subject was unable to complete one or more sections due to hearing impairment<br>• MoCA Total Raw Score - uncorrected<br>• MoCA Total Score - corrected for education<br>• MoCA: Visuospatial/executive - Trails<br>• MoCA: Visuospatial/executive - Cube<br>• MoCA: Visuospatial/executive - Clock contour<br>• MoCA: Visuospatial/executive - Clock numbers<br>• MoCA: Visuospatial/executive - Clock hands<br>• MoCA: Language - Naming<br>• MoCA: Memory - Registration (two trials)<br>• MoCA: Attention - Digits<br>• MoCA: Attention - Letter A<br>• MoCA: Attention - Serial 7s<br>• MoCA: Language - Repetition<br>• MoCA: Language - Fluency<br>• MoCA: Abstraction<br>• MoCA: Delayed recall - No cue<br>• MoCA: Delayed recall - Category cue<br>• MoCA: Delayed recall - Recognition<br>• MoCA: Orientation - Date<br>• MoCA: Orientation - Month<br>• MoCA: Orientation - Year<br>• MoCA: Orientation - Day<br>• MoCA: Orientation - Place<br>• MoCA: Orientation - City<br>• Craft Story 21 Recall (Immediate) - Total story units recalled, verbatim scoring<br>• Craft Story 21 Recall (Immediate) - Total story units recalled, paraphrase scoring<br>• Number Span Test: Forward - Number of correct trials<br>• Number Span Test: Forward - Longest span forward<br>• Number Span Test: backward - Number of correct trials<br>• Number Span Test: backward - Longest span backward<br>• Craft Story 21 Recall (Delayed) - Total story units recalled, verbatim scoring<br>• Craft Story 21 Recall (Delayed) - Total story units recalled, paraphrase scoring<br>• Craft Story 21 Recall (Delayed) - Delay time<br>• Craft Story 21 Recall (Delayed) - Cue (boy) needed<br>• Multilingual Naming Test (MINT) - Total score<br>• Multilingual Naming Test (MINT) - Total correct without semantic cue<br>• Multilingual Naming Test (MINT) - Semantic cues: Number given<br>• Multilingual Naming Test (MINT) - Semantic cues: Number correct with cue<br>• Multilingual Naming Test (MINT) - Phonemic cues: Number given<br>• Multilingual Naming Test (MINT) - Phonemic cues: Number correct with cue<br>• MoCA blind Total raw score - uncorrected<br>• MoCA-blind Total Score - corrected for education<br>• Rey Auditory Verbal Learning: Trial 1 total recall<br>• Rey Auditory Verbal Learning: Trial 1 intrusions<br>• Rey Auditory Verbal Learning: Trial 2 total recall<br>• Rey Auditory Verbal Learning: Trial 2 intrusions<br>• Rey Auditory Verbal Learning: Trial 3 total recall<br>• Rey Auditory Verbal Learning: Trial 3 intrusions<br>• Rey Auditory Verbal Learning: Trial 4 total recall<br>• Rey Auditory Verbal Learning: Trial 4 intrusions<br>• Rey Auditory Verbal Learning: Trial 5 total recall<br>• Rey Auditory Verbal Learning: Trial 5 intrusions<br>• Rey Auditory Verbal Learning: Trial 6 total recall<br>• Rey Auditory Verbal Learning: Trial 6 intrusions<br>• Oral Trail Making Test - Part A: Total number of seconds to complete<br>• Oral Trail Making Test - Part A: Number of commission errors<br>• Oral Trail Making Test - Part A: Number of correct lines<br>• Oral Trail Making Test Part B: Total number of seconds to complete<br>• Oral Trail Making Test Part B: Number of commission errors<br>• Oral Trail Making Test Part B: Number of correct lines<br>• Rey Auditory Verbal Learning: total delayed recall<br>• Rey Auditory Verbal Learning: delayed intrusions<br>• Rey Auditory Verbal Learning: recognition total correct<br>• Rey Auditory Verbal Learning: recognition total false positives<br>• Verbal naming test: total correct without a cue<br>• Verbal naming test: total correct with a phonemic cue<br>• How valid do you think the participant's responses are?<br>• What makes this participant's responses less valid? Hearing impairment<br>• What makes this participant's responses less valid? Distractions<br>• What makes this participant's responses less valid? Interruptions<br>• What makes this participant's responses less valid? Lack of effort or disinterest<br>• What makes this participant's responses less valid? Fatigue<br>• What makes this participant's responses less valid? Emotional issues<br>• What makes this participant's responses less valid? Unapproved assistance<br>• What makes this participant's responses less valid? Other<br>• In the past four weeks, did the subject have any difficulty or need help with: Writing checks, paying bills, or balancing a checkbook<br>• In the past four weeks, did the subject have any difficulty or need help with: Assembling tax records, business affairs, or other paper<br>• In the past four weeks, did the subject have any difficulty or need help with: Shopping alone for clothes, household necessities, or groceries<br>• In the past four weeks, did the subject have any difficulty or need help with: Playing a game of skill such as bridge or chess, working on a hobby<br>• In the past four weeks, did the subject have any difficulty or need help with: Heating water, making a cup of coffee, turning off the stove<br>• In the past four weeks, did the subject have any difficulty or need help with: Preparing a balanced meal<br>• In the past four weeks, did the subject have any difficulty or need help with: Keeping track of current events |

Table S2

| Cohort | Features |
|---|---|
| NACC | • In the past four weeks, did the subject have any difficulty or need help with: Paying attention to and understanding a TV program, book, or magazine<br>• In the past four weeks, did the subject have any difficulty or need help with: Remembering appointments, family occasions, holidays, medications<br>• In the past four weeks, did the subject have any difficulty or need help with: Traveling out of the neighborhood, driving, or arranging to take public transportation<br>• Is the subject able to complete the GDS, based on the clinician's best judgment?<br>• Are you basically satisfied with your life?<br>• Have you dropped many of your activities and interests?<br>• Do you feel that your life is empty?<br>• Do you often get bored?<br>• Are you in good spirits most of the time?<br>• Are you afraid that something bad is going to happen to you?<br>• Do you feel happy most of the time?<br>• Do you often feel helpless?<br>• Do you prefer to stay at home, rather than going out and doing new things?<br>• Do you feel you have more problems with memory than most?<br>• Do you think it is wonderful to be alive now?<br>• Do you feel pretty worthless the way you are now?<br>• Do you feel full of energy?<br>• Do you feel that your situation is hopeless?<br>• Do you think that most people are better off than you are?<br>• Total GDS Score<br>• Abrupt onset (re: cognitive status)<br>• Stepwise deterioration (re: cognitive status)<br>• Somatic complaints<br>• Emotional incontinence<br>• History or presence of hypertension<br>• History of stroke<br>• Focal neurological symptoms<br>• Focal neurological signs<br>• Hachinski ischemic score<br>• Cerebrovascular disease contributing to cognitive impairment<br>• Relationship between stroke and cognitive impairment<br>• Imaging evidence<br>• Single strategic infarct<br>• Multiple infarcts<br>• Extensive white matter hyperintensity<br>• Other imaging evidence<br>• Subject taking any medications<br>• Total number of medications reported at each visit<br>• Reported current use of any type of antihypertensive or blood pressure medication<br>• Reported current use of an antihypertensive combination therapy<br>• Reported current use of an angiotensin converting enzyme (ACE) inhibitor<br>• Reported current use of an antiadrenergic agent<br>• Reported current use of a beta-adrenergic blocking agent (beta-blocker)<br>• Reported current use of a calcium channel blocking agent<br>• Reported current use of a diuretic<br>• Reported current use of a vasodilator<br>• Reported current use of an angiotensin II inhibitor<br>• Reported current use of lipid lowering medication<br>• Reported current use of nonsteroidal anti-inflammatory medication<br>• Reported current use of an anticoagulant or antiplatelet agent<br>• Reported current use of an antidepressant<br>• Reported current use of an antipsychotic agent<br>• Reported current use of an FDA-approved medication for Alzheimer's disease symptoms<br>• Reported current use of an antiparkinson agent<br>• Reported current use of estrogen hormone therapy<br>• Reported current use of estrogen + progestin hormone therapy<br>• Reported current use of a diabetes medication<br>• Reported current use of an anxiolytic, sedative, or hypnotic agent<br>• UPDRS normal<br>• Speech<br>• Facial expression<br>• Tremor at rest - face, lips, chin<br>• Tremor at rest - right hand<br>• Tremor at rest - left hand<br>• Tremor at rest - right foot<br>• Tremor at rest - left foot<br>• Action or postural tremor - right hand<br>• Action or postural tremor - left hand<br>• Rigidity - neck<br>• Rigidity - right upper extremity<br>• Rigidity - left upper extremity<br>• Rigidity - right lower extremity<br>• Rigidity - left lower extremity<br>• Finger taps - right hand<br>• Finger taps - left hand<br>• Hand movements - right hand<br>• Hand movements - left hand<br>• Alternating movement - right hand<br>• Alternating movement - left hand<br>• Leg agility - right leg<br>• Leg agility - left leg<br>• Arising from chair<br>• Posture<br>• Gait<br>• Posture stability<br>• body bradykinesia and hypokinesia<br>• subject's height (inches)<br>• subject's weight (lbs)<br>• body mass index (BMI)<br>• Subject blood pressure (sitting), systolic<br>• Subject blood pressure (sitting), diastolic<br>• Subject resting heart rate (pulse)<br>• Without corrective lenses, is the subject's vision functionally normal?<br>• Does the subject usually wear corrective lenses?<br>• If the subject usually wears corrective lenses, is the subject's vision functionally normal with corrective lenses?<br>• Without a hearing aid(s), is the subject's hearing functionally normal?<br>• Does the subject usually wear a hearing aid(s)?<br>• If the subject usually wears a hearing aid(s), is the subject's hearing functionally normal with a hearing aid(s)?<br>• Imaging (MRI scans) |

Table S2: **Features from the NACC cohort.** This table shows the list of all the features extracted from the NACC cohort, which were used for model training.

| Cohort | Features | |
|---|---|---|
| AIBL | • Subject's age at visit<br>• Subject's sex<br>• Number of APOE e4 alleles<br>• Total number of story units recalled from this current test administration | • Logical Memory IIA - Delayed - Total number of story units recalled<br>• Total MMSE score (using D-L-R-O-W)<br>• Imaging (MRI scans) |
| NIFD | • Subject's age at visit<br>• Subject's sex<br>• Derived NIH race definitions<br>• Years of education<br>• Digit span forward length<br>• Digit span backward length | • Animals - Total number of animals named in 60 seconds<br>• Total MMSE score (using D-L-R-O-W)<br>• MoCA Total Score - corrected for education<br>• Total GDS Score<br>• Imaging (MRI scans) |
| PPMI | • Subject's age at visit<br>• Subject's sex<br>• Derived NIH race definitions<br>• Years of education<br>• Hispanic/Latino ethnicity | • Trail Making Test Part A - Total number of seconds to complete<br>• Trail Making Test Part B - Total number of seconds to complete<br>• MoCA Total Score - corrected for education<br>• Imaging (MRI scans) |
| OASIS | • Subject's age at visit<br>• Subject's sex<br>• Years of education<br>• Hispanic/Latino ethnicity<br>• Derived NIH race definitions<br>• Total years smoked cigarettes<br>• Pacemaker<br>• Cardiac bypass procedure<br>• Heart attack/cardiac arrest<br>• Congestive heart failure<br>• Atrial fibrillation<br>• Transient ischemic attack (TIA)<br>• Angioplasty/endarterectomy/stent<br>• Other cardiovascular disease<br>• Hypercholesterolemia<br>• Alcohol abuse - clinically significant occurring over a 12-month period manifested in one of the following areas: work, driving, legal, or social<br>• Other abused substances - clinically significant impairment occurring over a 12-month period manifested in one of the following areas: work, driving, legal, or social<br>• brain trauma - chronic deficit<br>• brain trauma - extended unconsciousness<br>• brain trauma - brief unconsciousness<br>• Diabetes<br>• Thyroid disease<br>• Vitamin B12 deficiency<br>• Incontinence - bowel<br>• Active depression in the last two years<br>• Depression episodes more than two years ago<br>• Other psychiatric disorder<br>• Seizures<br>• Hypertension<br>• Stroke<br>• Smoked more than 100 cigarettes in life<br>• Average number of packs smoked per day<br>• Traumatic brain injury (TBI)<br>• Incontinence - urinary<br>• Agitation or aggression severity<br>• Motor disturbance severity<br>• Delusions severity<br>• Disinhibition severity<br>• Hallucinations severity<br>• Depression or dysphoria severity<br>• Nighttime behavior severity<br>• Apathy or indifference severity<br>• Elation or euphoria severity | • Anxiety severity<br>• Appetite and eating severity<br>• Irritability or lability severity<br>• Number of APOE e4 alleles<br>• Digit span forward trials correct<br>• Digit span backward trials correct<br>• Digit span forward length<br>• Digit span backward length<br>• Total MMSE score (using D-L-R-O-W)<br>• Trail making test Part A - Total number of seconds to complete<br>• Trail making test Part B - Total number of seconds to complete<br>• Logical memory IIA - Delayed - Total number of story units recalled<br>• Total number of story units recalled from this current test administration<br>• Animals - Total number of animals named in 60 seconds<br>• Boston naming test (30) - Total score<br>• Total GDS score<br>• In the past four weeks, did the subject have any difficulty or need help with: Assembling tax records, business affairs, or other paper<br>• In the past four weeks, did the subject have any difficulty or need help with: Shopping alone for clothes, household necessities, or groceries<br>• In the past four weeks, did the subject have any difficulty or need help with: Writing checks, paying bills, or balancing a checkbook<br>• In the past four weeks, did the subject have any difficulty or need help with: Heating water, making a cup of coffee, turning off the stove<br>• In the past four weeks, did the subject have any difficulty or need help with: Playing a game of skill such as bridge or chess, working on a hobby<br>• In the past four weeks, did the subject have any difficulty or need help with: Traveling out of the neighborhood, driving, or arranging to take public transportation<br>• In the past four weeks, did the subject have any difficulty or need help with: Paying attention to and understanding a TV program, book, or magazine<br>• In the past four weeks, did the subject have any difficulty or need help with: Preparing a balanced meal<br>• In the past four weeks, did the subject have any difficulty or need help with: Keeping track of current events<br>• In the past four weeks, did the subject have any difficulty or need help with: Remembering appointments, family occasions, holidays, medications<br>• Imaging (MRI scans) |

Table S3: **Features from the AIBL, NIFD, PPMI, and OASIS cohorts.** This table provides a systematic enumeration of the variables extracted from the AIBL, NIFD, PPMI and OASIS cohorts, illustrating the range of features employed in our analytical model and emphasizing the breadth of the dataset compilation.

| Cohort | Features | |
|---|---|---|
| LBDSU | • Subject's age at visit<br>• Subject's sex<br>• Derived NIH race definitions<br>• Years of education | • Hispanic/Latino ethnicity<br>• MoCA Total Score - corrected for education<br>• Imaging (MRI scans) |
| 4RTNI | • Subject's sex<br>• Subject's age at visit<br>• Years of education<br>• Hispanic/Latino ethnicity<br>• Derived NIH race definitions<br>• Agitation or aggression severity<br>• Motor disturbance severity<br>• Delusions severity<br>• Disinhibition severity<br>• Hallucinations severity<br>• Depression or dysphoria severity<br>• Nighttime behavior severity<br>• Apathy or indifference severity | • Elation or euphoria severity<br>• Anxiety severity<br>• Appetite and eating severity<br>• Irritability or lability severity<br>• UPDRS normal<br>• Total MMSE score (using D-L-R-O-W)<br>• MoCA Total Score - corrected for education<br>• Trail making test Part A - Total number of seconds to complete<br>• Trail making test Part B - Total number of seconds to complete<br>• Part A - Number of correct lines<br>• Part B - Number of correct lines<br>• Total GDS Score<br>• Imaging (MRI scans) |
| FHS | • Subject's sex<br>• Subject's age at visit<br>• Hispanic/Latino ethnicity<br>• Race<br>• Derived NIH race definitions<br>• Marital status<br>• Left- or right-handedness<br>• subject's weight (lbs)<br>• subject's height (inches)<br>• Body mass index (BMI) | • Blood pressure (sitting), systolic<br>• Blood pressure (sitting), diastolic<br>• Smoked cigarettes in last 30 days<br>• Total MMSE score (using D-L-R-O-W)<br>• Boston naming test (30) - Total score<br>• History of stroke<br>• Reported current use of a diabetes medication<br>• Reported current use of lipid lowering medication<br>• Imaging (MRI scans) |

Table S4: **Features from the LBDSU, 4RTNI and FHS cohorts.** This table enumerates the features collected from the LBDSU, 4RTNI, and FHS cohorts, illustrating the range of features employed in our analytical model and emphasizing the breadth of the dataset compilation. Of note, FHS was used as an external dataset to validate our model's predictive performance.

| Cohort | Features |
|---|---|
| ADNI | • Subject's age at visit<br>• Subject's sex<br>• Years of education<br>• Hispanic/Latino ethnicity<br>• Derived NIH race definitions<br>• Primary language<br>• Marital status<br>• Type of residence<br>• Is the subject left- or right-handed?<br>• Indicator of mother with cognitive impairment<br>• Indicator of father with cognitive impairment<br>• Orientation subscale score - Time<br>• Orientation subscale score - Place<br>• Total years smoked cigarettes<br>• Heart attack/cardiac arrest<br>• Hypertension<br>• History or presence of hypertension<br>• Stroke<br>• History of stroke<br>• Focal neurological symptoms<br>• Focal neurological signs<br>• Hachinski ischemic score<br>• Subject's height (inches)<br>• Subject's weight (lbs)<br>• Subject blood pressure (sitting), systolic<br>• Subject blood pressure (sitting), diastolic<br>• Subject resting heart rate (pulse)<br>• Depression episodes more than two years ago<br>• Are you basically satisfied with your life?<br>• Have you dropped many of your activities and interests?<br>• Do you feel that your life is empty<br>• Do you often get bored?<br>• Are you in good spirits most of the time?<br>• Are you afraid that something bad is going to happen to you?<br>• Do you feel happy most of the time?<br>• Do you often feel helpless?<br>• Do you prefer to stay at home, rather than going out and doing new things?<br>• Do you feel you have more problems with memory than most?<br>• Do you think it is wonderful to be alive now?<br>• Do you feel pretty worthless the way you are now<br>• Do you feel full of energy?<br>• Do you feel that your situation is hopeless?<br>• Do you think that most people are better off than you are?<br>• Abrupt onset (re: cognitive status)<br>• Stepwise deterioration (re: cognitive status)<br>• Somatic complaints<br>• Emotional incontinence<br>• Other psychiatric disorder<br>• Indicator of first-degree family member with cognitive impairment<br>• Alcohol abuse - clinically significant occurring over a 12-month period manifested in one of the following areas: work, driving, legal, or social<br>• Agitation or aggression severity<br>• Motor disturbance severity<br>• Delusions severity<br>• Disinhibition severity<br>• Hallucinations severity<br>• Depression or dysphoria severity<br>• Nighttime behavior severity<br>• Apathy or indifference severity<br>• Elation or euphoria severity<br>• Anxiety severity<br>• Appetite and eating severity<br>• Irritability or lability severity<br>• Number of APOE e4 alleles<br>• Multilingual Naming Test (MINT) - Total score<br>• Digit span forward trials correct<br>• Digit span backward trials correct<br>• Digit span forward length<br>• Digit span backward length<br>• Total MMSE score (using D-L-R-O-W)<br>• Trail making test Part A - Total number of seconds to complete<br>• Trail making test Part B - Total number of seconds to complete<br>• Logical memory IIA - Delayed - Total number of story units recalled<br>• Total number of story units recalled from this current test administration<br>• Animals - Total number of animals named in 60 seconds<br>• MoCA Total Score - corrected for education<br>• MoCA: Visuospatial/executive - Trails<br>• MoCA: Visuospatial/executive - Cube<br>• MoCA: Visuospatial/executive - Clock contour<br>• MoCA: Visuospatial/executive - Clock numbers<br>• MoCA: Visuospatial/executive - Clock hands<br>• MoCA: Language - Naming<br>• MoCA: Attention - Digits<br>• MoCA: Attention - Letter A<br>• MoCA: Attention - Serial 7s<br>• MoCA: Language - Repetition<br>• MoCA: Language - Fluency<br>• MoCA: Abstraction<br>• MoCA: Delayed recall - No cue<br>• MoCA: Orientation - Date<br>• MoCA: Orientation - Month<br>• MoCA: Orientation - Year<br>• MoCA: Orientation - Day<br>• MoCA: Orientation - Place<br>• MoCA: Orientation - City<br>• Boston naming test (30) - Total score<br>• Total GDS score<br>• In the past four weeks, did the subject have any difficulty or need help with: Assembling tax records, business affairs, or other paper<br>• In the past four weeks, did the subject have any difficulty or need help with: Shopping alone for clothes, household necessities, or groceries<br>• In the past four weeks, did the subject have any difficulty or need help with: Writing checks, paying bills, or balancing a checkbook<br>• In the past four weeks, did the subject have any difficulty or need help with: Heating water, making a cup of coffee, turning off the stove<br>• In the past four weeks, did the subject have any difficulty or need help with: Playing a game of skill such as bridge or chess, working on a hobby<br>• In the past four weeks, did the subject have any difficulty or need help with: Traveling out of the neighborhood, driving, or arranging to take public transportation<br>• In the past four weeks, did the subject have any difficulty or need help with: Paying attention to and understanding a TV program, book, or magazine<br>• In the past four weeks, did the subject have any difficulty or need help with: Preparing a balanced meal<br>• In the past four weeks, did the subject have any difficulty or need help with: Keeping track of current events<br>• In the past four weeks, did the subject have any difficulty or need help with: Remembering appointments, family occasions, holidays, medications<br>• Imaging (MRI scans) |

Table S5: **Features from the ADNI cohort.** This table shows the list of all the features extracted from the ADNI cohort, which were used for model testing.

| Dataset (group) | T1 | T2 | FLAIR | SWI |
|---|---|---|---|---|
| NACC | 1970 | 352 | 318 | 32 |
| NIFD | 633 | 414 | 537 | 3 |
| PPMI | 241 | N.A. | N.A. | N.A. |
| AIBL | 681 | N.A. | 334 | N.A. |
| OASIS | 662 | N.A. | N.A. | N.A. |
| LBDSU | 181 | N.A. | N.A. | N.A. |
| 4RTNI | 165 | 119 | 120 | N.A. |
| ADNI | 1055 | N.A. | N.A. | N.A. |
| FHS | 115 | 109 | 114 | N.A. |

Table S6: **MRI sequences used for model development.** T1-weighted, T2-weighted, fluid attenuated inversion recovery and susceptibility weighted imaging were included from NACC, NIFD, PPMI, AIBL, OASIS, LDBSU and 4RTNI for model training. A portion of MRIs from the NACC dataset along with MRIs from ADNI, and FHS were reserved for model testing.

| Dataset (group) | Balanced Accuracy | Precision | Sensitivity | Specificity | F1 Score | MCC | AUROC | AUPR |
|---|---|---|---|---|---|---|---|---|
| NACC | | | | | | | | |
| NC | 0.93 | 0.9 | 0.92 | 0.94 | 0.91 | 0.85 | 0.98 | 0.97 |
| MCI | 0.83 | 0.52 | 0.80 | 0.85 | 0.63 | 0.56 | 0.91 | 0.67 |
| DE | 0.94 | 0.92 | 0.94 | 0.93 | 0.93 | 0.87 | 0.99 | 0.98 |
| AD | 0.89 | 0.84 | 0.87 | 0.91 | 0.86 | 0.78 | 0.96 | 0.93 |
| LBD | 0.87 | 0.43 | 0.80 | 0.95 | 0.56 | 0.56 | 0.96 | 0.68 |
| VD | 0.83 | 0.28 | 0.74 | 0.92 | 0.41 | 0.42 | 0.93 | 0.47 |
| PRD | 0.67 | 0.09 | 0.35 | 0.99 | 0.14 | 0.17 | 0.96 | 0.12 |
| FTD | 0.89 | 0.36 | 0.90 | 0.89 | 0.51 | 0.53 | 0.96 | 0.67 |
| NPH | 0.55 | 0.12 | 0.11 | 1.00 | 0.12 | 0.12 | 0.91 | 0.077 |
| SEF | 0.66 | 0.069 | 0.42 | 0.90 | 0.12 | 0.13 | 0.82 | 0.064 |
| PSY | 0.79 | 0.24 | 0.71 | 0.86 | 0.36 | 0.36 | 0.90 | 0.36 |
| TBI | 0.62 | 0.07 | 0.26 | 0.98 | 0.11 | 0.12 | 0.90 | 0.098 |
| ODE | 0.68 | 0.11 | 0.46 | 0.89 | 0.17 | 0.18 | 0.84 | 0.11 |
| ADNI | | | | | | | | |
| NC | 0.83 | 0.64 | 0.97 | 0.69 | 0.77 | 0.64 | 0.94 | 0.89 |
| MCI | 0.76 | 0.82 | 0.63 | 0.88 | 0.71 | 0.53 | 0.87 | 0.83 |
| DE | 0.90 | 0.64 | 0.91 | 0.89 | 0.75 | 0.71 | 0.97 | 0.88 |
| AD | 0.91 | 0.70 | 0.89 | 0.92 | 0.78 | 0.74 | 0.97 | 0.86 |
| FHS | | | | | | | | |
| NC | 0.59 | 0.35 | 0.42 | 0.76 | 0.38 | 0.17 | 0.66 | 0.33 |
| MCI | 0.53 | 0.40 | 0.13 | 0.93 | 0.20 | 0.098 | 0.59 | 0.34 |
| DE | 0.68 | 0.68 | 0.68 | 0.68 | 0.68 | 0.36 | 0.73 | 0.71 |
| AD | 0.65 | 0.63 | 0.52 | 0.78 | 0.57 | 0.32 | 0.72 | 0.64 |
| LBD | 0.52 | 0.077 | 0.068 | 0.96 | 0.072 | 0.032 | 0.62 | 0.071 |
| VD | 0.65 | 0.18 | 0.44 | 0.85 | 0.26 | 0.20 | 0.74 | 0.30 |
| FTD | 0.59 | 0.016 | 0.25 | 0.92 | 0.03 | 0.045 | 0.71 | 0.028 |

Table S7: **Model performance.** This table presents the performance metrics of our model across the NACC, ADNI, and FHS datasets. Specifically, the results for the NACC testing dataset are based on the input features outlined in Table S2. For the ADNI and FHS datasets, the results are derived from a restricted set of input features detailed in Table S4, S5. Of note, these results are influenced by the use of a limited selection of input features. Despite this limitation, the model, which was initially trained on the NACC data incorporating a broader feature set, demonstrates the capability to generalize and make predictions on the ADNI and FHS datasets. This indicates the model's robustness and its potential to yield predictions even with significant missing input feature information, albeit with some reduction in performance. Demographic information for each cohort can be found in Table S1.

| Dataset (group) | Balanced Accuracy | | Precision | | Sensitivity | | Specificity | | F1 Score | | MCC | | AUROC | | AUPR | |
|---|---|---|---|---|---|---|---|---|---|---|---|---|---|---|---|---|
| NACC | Our | CatBoost | Our | CatBoost | Our | CatBoost | Our | CatBoost | Our | CatBoost | Our | CatBoost | Our | CatBoost | Our | CatBoost |
| NC | 0.71 | 0.74 | 0.53 | 0.60 | 0.87 | 0.80 | 0.55 | 0.69 | 0.66 | 0.69 | 0.41 | 0.47 | 0.79 | 0.84 | 0.69 | 0.74 |
| MCI | 0.53 | 0.52 | 0.34 | 0.38 | 0.11 | 0.05 | 0.96 | 0.98 | 0.17 | 0.09 | 0.11 | 0.09 | 0.66 | 0.71 | 0.26 | 0.30 |
| DE | 0.73 | 0.75 | 0.90 | 0.72 | 0.51 | 0.76 | 0.95 | 0.75 | 0.65 | 0.74 | 0.52 | 0.51 | 0.82 | 0.86 | 0.83 | 0.86 |
| ADNI | | | | | | | | | | | | | | | | |
| NC | 0.66 | 0.70 | 0.45 | 0.55 | 0.99 | 0.75 | 0.33 | 0.66 | 0.62 | 0.63 | 0.37 | 0.39 | 0.81 | 0.77 | 0.62 | 0.59 |
| MCI | 0.53 | 0.56 | 0.68 | 0.58 | 0.11 | 0.32 | 0.95 | 0.79 | 0.20 | 0.42 | 0.13 | 0.14 | 0.64 | 0.63 | 0.59 | 0.55 |
| DE | 0.77 | 0.83 | 0.86 | 0.55 | 0.55 | 0.79 | 0.98 | 0.86 | 0.67 | 0.65 | 0.64 | 0.57 | 0.95 | 0.94 | 0.81 | 0.80 |
| FHS | | | | | | | | | | | | | | | | |
| NC | 0.69 | 0.53 | 0.38 | 0.26 | 0.78 | 0.56 | 0.60 | 0.50 | 0.51 | 0.35 | 0.32 | 0.05 | 0.73 | 0.51 | 0.42 | 0.22 |
| MCI | 0.51 | 0.51 | 0.34 | 0.67 | 0.03 | 0.02 | 0.98 | 1.00 | 0.06 | 0.04 | 0.03 | 0.09 | 0.66 | 0.55 | 0.36 | 0.32 |
| DE | 0.65 | 0.57 | 0.73 | 0.58 | 0.48 | 0.55 | 0.82 | 0.59 | 0.58 | 0.56 | 0.32 | 0.15 | 0.74 | 0.59 | 0.73 | 0.65 |

(a)

| Dataset (group) | Balanced Accuracy | | Precision | | Sensitivity | | Specificity | | F1 Score | | MCC | | AUROC | | AUPR | |
|---|---|---|---|---|---|---|---|---|---|---|---|---|---|---|---|---|
| NACC | Our | CatBoost | Our | CatBoost | Our | CatBoost | Our | CatBoost | Our | CatBoost | Our | CatBoost | Our | CatBoost | Our | CatBoost |
| NC | 0.84 | 0.87 | 0.69 | 0.79 | 0.92 | 0.87 | 0.75 | 0.86 | 0.79 | 0.83 | 0.65 | 0.72 | 0.93 | 0.94 | 0.88 | 0.90 |
| MCI | 0.65 | 0.59 | 0.35 | 0.45 | 0.48 | 0.23 | 0.82 | 0.94 | 0.41 | 0.31 | 0.27 | 0.24 | 0.75 | 0.79 | 0.33 | 0.39 |
| DE | 0.83 | 0.88 | 0.90 | 0.83 | 0.74 | 0.91 | 0.93 | 0.85 | 0.81 | 0.87 | 0.69 | 0.76 | 0.94 | 0.95 | 0.93 | 0.94 |
| ADNI | | | | | | | | | | | | | | | | |
| NC | 0.76 | 0.74 | 0.54 | 0.67 | 0.99 | 0.66 | 0.53 | 0.82 | 0.70 | 0.66 | 0.52 | 0.47 | 0.91 | 0.87 | 0.84 | 0.78 |
| MCI | 0.67 | 0.62 | 0.75 | 0.62 | 0.49 | 0.51 | 0.86 | 0.73 | 0.59 | 0.56 | 0.38 | 0.25 | 0.79 | 0.67 | 0.73 | 0.56 |
| DE | 0.88 | 0.86 | 0.71 | 0.57 | 0.82 | 0.86 | 0.93 | 0.86 | 0.76 | 0.68 | 0.71 | 0.62 | 0.96 | 0.94 | 0.84 | 0.81 |
| FHS | | | | | | | | | | | | | | | | |
| NC | 0.69 | 0.52 | 0.38 | 0.45 | 0.78 | 0.07 | 0.60 | 0.97 | 0.51 | 0.13 | 0.32 | 0.10 | 0.73 | 0.52 | 0.42 | 0.26 |
| MCI | 0.51 | 0.50 | 0.34 | 0.00 | 0.03 | 0.00 | 0.98 | 1.00 | 0.06 | 0.00 | 0.03 | 0.00 | 0.66 | 0.60 | 0.36 | 0.34 |
| DE | 0.65 | 0.52 | 0.73 | 0.51 | 0.48 | 0.98 | 0.82 | 0.06 | 0.58 | 0.67 | 0.32 | 0.12 | 0.74 | 0.60 | 0.73 | 0.65 |

(b)

Table S8: **Model performance comparison with CatBoost.** This table presents the performance comparison between our model and CatBoost across the NACC, ADNI, and FHS datasets on NC, MCI, DE using two different subsets of features. The first subset was composed of common demographics information, as well as MMSE and Boston Naming Test scores. The second subset created on the first subset by incorporating additional neuropsychological measures found in the NACC and ADNI cohorts, such as trail making tests A and B, logical memory IIA delayed recall, MoCA scores, and digit span forward and backward tests. The unavailable features in the ADNI and FHS dataset are imputed for the CatBoost model. (a) Results with the first subset. (b) Results with the second subset. These findings indicate that our model has better generalization capabilities compared to Catboost when applied to external cohorts.

| Comparison | KS 2samp statistic | p-value |
|---|---|---|
| No AD and AD | 0.09 | 4.29e-3 |

Table S9: **Statistical analysis of model alignment with prodromal AD.** Two-sample Kolmogorov-Smirnov test for goodness of fit statistics for prodromal disease plots indicating statistical significance between MCI cases with AD as an etiological factor compared to those without AD.

| Cohort | Kruskal statistic | p-value |
|---|---|---|
| NACC | 6921.73 | $<$1e-200 |
| ADNI | 1518.79 | $<$1e-200 |
| FHS | 292.04 | 3.84e-64 |

Table S10: **Statistical analysis of model alignment with clinical dementia rating (CDR).** Significant Kruskal-Wallis H-test statistics observed in the CDR plots reveal substantial variability among the CDR groups. Lower p-values ($p < 0.0001$) indicate that there is a statistically significant difference in the distribution of dementia probabilities across at least two of the CDR groups. We then performed Dunn-Bonferroni posthoc tests for detailed pairwise comparisons among the CDR groups within each cohort.

| Etiology | Cohort | Biomarker | Statistic | p-value |
|---|---|---|---|---|
| AD | NACC | A$\beta$ PET | 10303.50$^\dagger$ | 2.04e-25 |
| AD | NACC | Tau PET | 935.50$^\dagger$ | 6.48e-8 |
| AD | NACC | FDG PET | 3730.0$^\dagger$ | 3.00e-15 |
| AD | ADNI | A$\beta$ PET | -12.06$^\ddagger$ | 9.74e-31 |
| AD | ADNI | Tau PET | 5857.50$^\dagger$ | 4.10e-27 |
| AD | ADNI | FDG PET | 14924.0$^\dagger$ | 5.66e-43 |
| FTD | NACC | MR | 30935.50$^\dagger$ | 1.52e-51 |
| FTD | NACC | FDG PET | 1599.50$^\dagger$ | 2.08e-13 |
| PD | NACC | DaTScan | 318.50$^\dagger$ | 6.26e-06 |

Table S11: **Statistical analysis of biomarker validation.** Statistics for biomarker validation plots indicating statistical significance between biomarker negative and positive groups across etiologies. † indicates Mann-Whitney U test; ‡ indicates independent samples t-test.

| Cohort | Age at death (years) mean ± std | Male gender (percentage) | CDR mean ± std |
|---|---|---|---|
| NACC | | | |
| AD [n = 131] | 79.28 ± 10.61 | 74, 56.49% | 1.44 ± 0.75 |
| LBD [n = 44] | 76.64 ± 8.31 | 36, 81.82% | 1.44 ± 0.66 |
| VD [n = 15] | 83.73 ± 10.39 | 7, 46.67% | 1.37 ± 0.86 |
| PRD [n = 5] | 562.6 ± 4.96 | 5, 100% | 0.7 ± 0.24 |
| FTD [n = 20] | 67.15 ± 8.61 | 13, 65% | 1.82 ± 0.98 |
| NPH [n = 1] | 77 ± 0.0 | 1, 100% | 0.5 ± 0.0 |
| SEF [n = 3] | 73 ± 24.04 | 2, 66.67% | 0.83 ± 0.23 |
| TBI [n = 1] | 87 ± 0.0 | 1, 100% | 2.0 ± 0.0 |
| ODE [n = 11] | 68.82 ± 14.99 | 7, 63.64% | 1.45 ± 0.86 |
| *p-value* | 1.428e-08 | 5.956e-02 | 1.471e-01 |
| ADNI | | | |
| AD [n = 19] | 82.05 ± 9.03 | 13, 68.42% | 0.86 ± 0.22 |
| FHS | | | |
| AD [n = 55] | 91 ± 7.0 | 18, 32.72% | 1.01 ± 0.15 |
| LBD [n = 4] | 92.75 ± 2.16 | 3, 75% | 1.0 ± 0.0 |
| VD [n = 5] | 93 ± 3.35 | 4, 80% | 1.2 ± 0.4 |
| FTD [n = 2] | 79 ± 4.0 | 1, 50% | 1.0 ± 0.0 |
| *p-value* | 1.584e-01 | 8.227e-02 | 2.972e-01 |

Table S12: **Cases with post mortem findings used for model validation.** Model validation was conducted using cases with post-mortem findings from three independent datasets: ADNI, NACC, and FHS. Continuous variables were analyzed using one-way ANOVA, while categorical variables were assessed with $\chi^2$ tests. The p-values derived for each dataset reflect the statistical significance of differences between groups for each column.

| Etiology | Neuropath | Kruskal statistics |
|---|---|---|
| AD | A score | 144.91 |
| AD | B score | 358.69 |
| AD | C score | 147.04 |
| AD | Cerebral amyloid angiopathy | 46.17 |
| VD | Old microinfarcts | 33.74 |
| VD | Arteriolosclerosis | 185.18 |
| FTD | Tauopathy | 95.99 |
| FTD | FTLD with TDP-43 pathology | 164.20 |

Table S13: **Statistical analysis of neuropathological validation.** Kruskal-Wallis H-test statistics for the neuropathological validation plots, which indicate statistical significance between dementia etiologies and neuropathological indicators. Since the p-values for all plots are very small ($p < 0.0001$), we applied Dunn-Bonferroni post-hoc comparisons between neuropathological scores to determine which groups are different in each etiology.

| Etiology | Rater | AUC Median | AUC IQR | Wilcoxon signed rank test p-value |
|---|---|---|---|---|
| NC | Neurologist | 0.930 | 0.034 | 4.88e-04 |
| | AI-augmented Neurologist | 0.980 | 0.011 | |
| MCI | Neurologist | 0.699 | 0.115 | 4.88e-04 |
| | AI-augmented Neurologist | 0.793 | 0.063 | |
| DE | Neurologist | 0.914 | 0.094 | 2.44e-04 |
| | AI-augmented Neurologist | 0.954 | 0.022 | |
| AD | Neurologist | 0.761 | 0.042 | 2.44e-04 |
| | AI-augmented Neurologist | 0.866 | 0.026 | |
| LBD | Neurologist | 0.833 | 0.080 | 2.44e-04 |
| | AI-augmented Neurologist | 0.949 | 0.025 | |
| VD | Neurologist | 0.613 | 0.125 | 2.44e-04 |
| | AI-augmented Neurologist | 0.846 | 0.043 | |
| PRD | Neurologist | 0.517 | 0.093 | 2.44e-04 |
| | AI-augmented Neurologist | 0.899 | 0.015 | |
| FTD | Neurologist | 0.708 | 0.106 | 2.44e-04 |
| | AI-augmented Neurologist | 0.895 | 0.043 | |
| NPH | Neurologist | 0.719 | 0.153 | 2.44e-04 |
| | AI-augmented Neurologist | 0.844 | 0.075 | |
| SEF | Neurologist | 0.517 | 0.140 | 2.44e-04 |
| | AI-augmented Neurologist | 0.704 | 0.075 | |
| PSY | Neurologist | 0.613 | 0.073 | 2.44e-04 |
| | AI-augmented Neurologist | 0.789 | 0.021 | |
| TBI | Neurologist | 0.497 | 0.098 | 2.44e-04 |
| | AI-augmented Neurologist | 0.875 | 0.023 | |
| ODE | Neurologist | 0.516 | 0.082 | 6.10e-03 |
| | AI-augmented Neurologist | 0.558 | 0.042 | |
| AD | Radiologist | 0.632 | 0.104 | 7.81e-03 |
| | AI-augmented Radiologist | 0.672 | 0.092 | |
| LBD | Radiologist | 0.490 | 0.068 | 1.56e-02 |
| | AI-augmented Radiologist | 0.577 | 0.074 | |
| VD | Radiologist | 0.657 | 0.057 | 1.56e-02 |
| | AI-augmented Radiologist | 0.720 | 0.022 | |
| PRD | Radiologist | 0.541 | 0.089 | 7.81e-03 |
| | AI-augmented Radiologist | 0.916 | 0.029 | |
| FTD | Radiologist | 0.734 | 0.126 | 7.81e-03 |
| | AI-augmented Radiologist | 0.756 | 0.063 | |
| NPH | Radiologist | 0.739 | 0.131 | 7.81e-03 |
| | AI-augmented Radiologist | 0.853 | 0.062 | |
| SEF | Radiologist | 0.454 | 0.163 | 7.81e-03 |
| | AI-augmented Radiologist | 0.548 | 0.172 | |
| PSY | Radiologist | 0.542 | 0.122 | 1.56e-02 |
| | AI-augmented Radiologist | 0.615 | 0.064 | |
| TBI | Radiologist | 0.492 | 0.075 | 2.34e-02 |
| | AI-augmented Radiologist | 0.545 | 0.055 | |
| ODE | Radiologist | 0.480 | 0.030 | 7.81e-03 |
| | AI-augmented Radiologist | 0.558 | 0.051 | |

Table S14: **Statistical analysis of AI-augmented clinician AUROCs.** Statistics for individual neurologists' AUROC and AI-augmented neurologists' AUROC. One-tailed Wilcoxon signed-rank test was performed.

| Etiology | Rater | AP Median | AP IQR | Wilcoxon signed-rank test p-value |
|---|---|---|---|---|
| NC | Neurologist | 0.790 | 0.098 | 4.88e-04 |
| | AI-augmented Neurologist | 0.911 | 0.023 | |
| MCI | Neurologist | 0.301 | 0.105 | 4.88e-04 |
| | AI-augmented Neurologist | 0.411 | 0.074 | |
| DE | Neurologist | 0.942 | 0.051 | 2.44e-04 |
| | AI-augmented Neurologist | 0.977 | 0.013 | |
| AD | Neurologist | 0.667 | 0.037 | 2.44e-04 |
| | AI-augmented Neurologist | 0.830 | 0.046 | |
| LBD | Neurologist | 0.439 | 0.174 | 2.44e-04 |
| | AI-augmented Neurologist | 0.740 | 0.127 | |
| VD | Neurologist | 0.225 | 0.088 | 2.44e-04 |
| | AI-augmented Neurologist | 0.451 | 0.084 | |
| PRD | Neurologist | 0.081 | 0.032 | 2.44e-04 |
| | AI-augmented Neurologist | 0.327 | 0.051 | |
| FTD | Neurologist | 0.344 | 0.170 | 2.44e-04 |
| | AI-augmented Neurologist | 0.574 | 0.238 | |
| NPH | Neurologist | 0.321 | 0.131 | 2.12e-02 |
| | AI-augmented Neurologist | 0.401 | 0.130 | |
| SEF | Neurologist | 0.145 | 0.113 | 2.44e-04 |
| | AI-augmented Neurologist | 0.254 | 0.112 | |
| PSY | Neurologist | 0.265 | 0.112 | 4.88e-04 |
| | AI-augmented Neurologist | 0.452 | 0.054 | |
| TBI | Neurologist | 0.071 | 0.038 | 2.44e-04 |
| | AI-augmented Neurologist | 0.345 | 0.078 | |
| ODE | Neurologist | 0.113 | 0.091 | 8.06e-03 |
| | AI-augmented Neurologist | 0.120 | 0.106 | |
| AD | Radiologist | 0.728 | 0.112 | 5.47e-02 |
| | AI-augmented Radiologist | 0.727 | 0.073 | |
| LBD | Radiologist | 0.129 | 0.037 | 1.56e-02 |
| | AI-augmented Radiologist | 0.177 | 0.131 | |
| VD | Radiologist | 0.353 | 0.088 | 1.56e-02 |
| | AI-augmented Radiologist | 0.385 | 0.097 | |
| PRD | Radiologist | 0.136 | 0.061 | 7.81e-03 |
| | AI-augmented Radiologist | 0.415 | 0.073 | |
| FTD | Radiologist | 0.420 | 0.196 | 1.56e-02 |
| | AI-augmented Radiologist | 0.553 | 0.177 | |
| NPH | Radiologist | 0.433 | 0.239 | 3.91e-02 |
| | AI-augmented Radiologist | 0.432 | 0.302 | |
| SEF | Radiologist | 0.111 | 0.073 | 7.81e-03 |
| | AI-augmented Radiologist | 0.157 | 0.146 | |
| PSY | Radiologist | 0.259 | 0.063 | 1.56e-02 |
| | AI-augmented Radiologist | 0.305 | 0.055 | |
| TBI | Radiologist | 0.104 | 0.016 | 7.81e-03 |
| | AI-augmented Radiologist | 0.180 | 0.034 | |
| ODE | Radiologist | 0.137 | 0.044 | 7.81e-03 |
| | AI-augmented Radiologist | 0.154 | 0.041 | |

Table S15: **Statistical analysis of AI-augmented clinician APs.** Statistics for individual neurologists' AP and AI-augmented neurologists' AP. One-tailed Wilcoxon signed-rank test was performed.

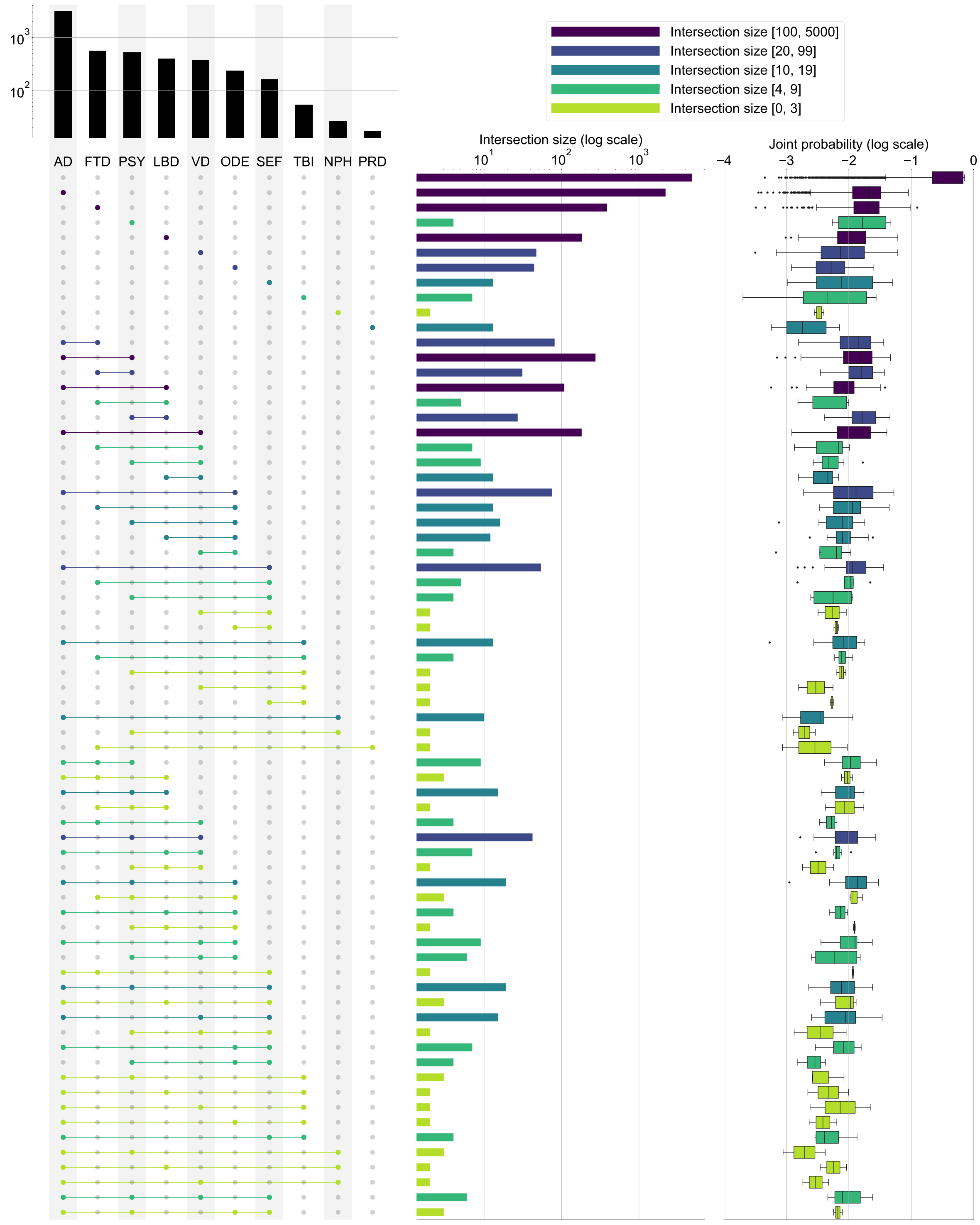


Figure S1: **Model assessment on mixed dementias.** Visualization of the distribution and model-predicted probabilities of various etiological categories found within the NACC testing dataset. The left segment enumerates both single and co-occurring diagnostic categories, offering a tally of each condition's frequency within the dataset. In the center, a logarithmic scale is used to delineate the overlap among these categories, shedding light on their relative commonality and the extent of their coexistence. This method grants a refined perspective on the prevalence of comorbid conditions. Additionally, the legend in the upper right interprets the counts within the central panel, providing a reference for the logarithmic data representation. The panel on the far right features a box-and-whisker plot, delineating the spread and central tendency of the model's predicted probabilities for each combination of diagnostic categories.

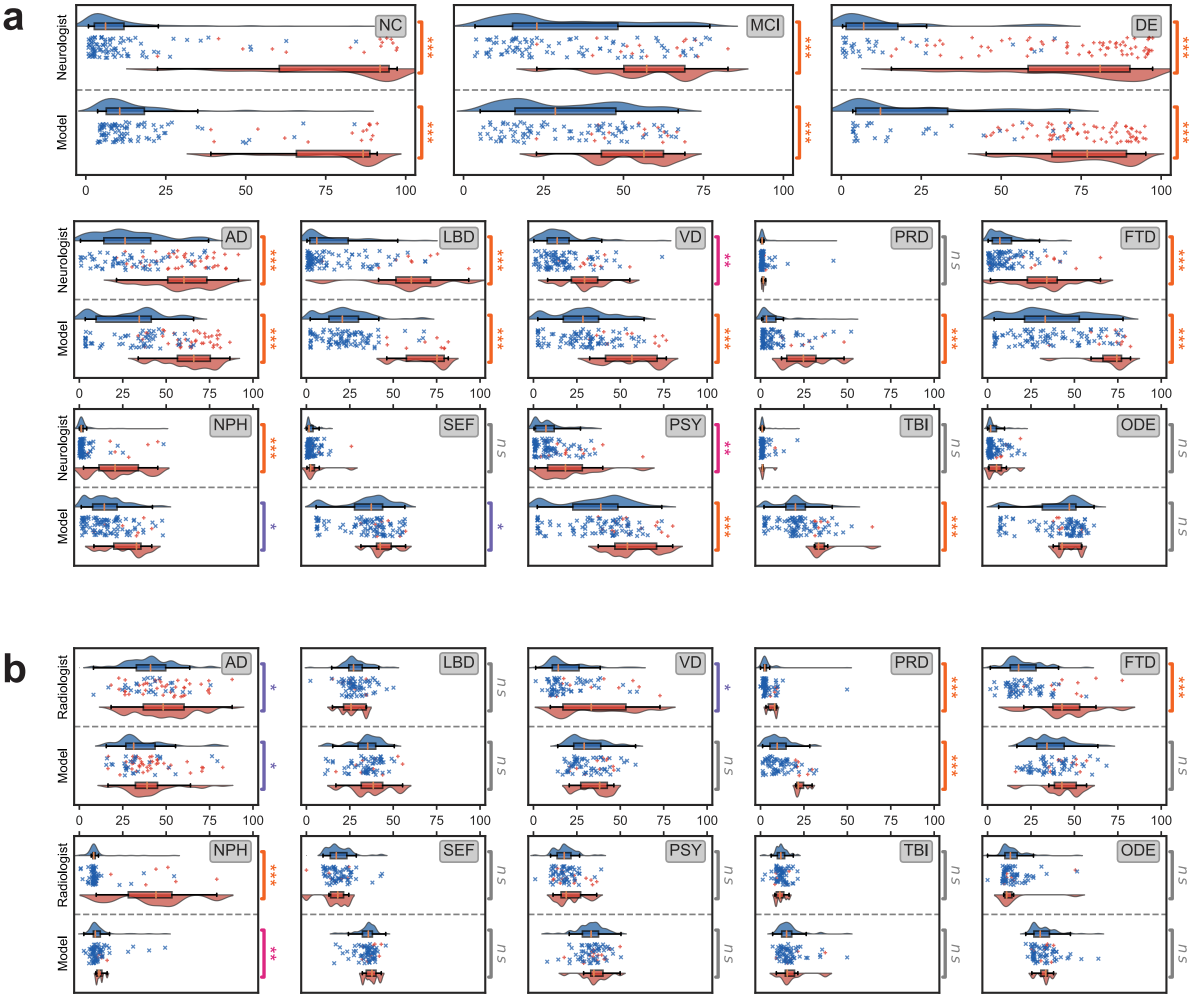


Figure S2: **Head to head comparison between model and clinicians** Comparison between model-predicted probability scores and the assessments provided by practicing clinicians is shown. (a) For the analysis, neurologists were given 100 randomly selected cases encompassing individual-level demographics, health history, neurological tests, physical as well as neurological examinations, and multi-sequence MRI scans. The neurologists were then tasked with assigning confidence scores for NC, MCI, DE, and the 10 dementia etiologies: AD, LBD, VD, PRD, FTD, NPH, SEF, PSY, TBI, and ODE (see Glossary 1). Neurologists' confidence scores were averaged to produce a single consensus confidence score for each case. In the visual representation, the boxplot in blue indicates the distribution of confidence scores for true negative cases, while the boxplot in red signifies true positive cases. The symbol '+' represents true positive cases, and 'x' denotes true negative cases. Significance levels are denoted as: ns (not significant) for $p \geq 0.05$; * for $p < 0.05$; ** for $p < 0.01$; *** for $p < 0.001$; and **** for $p < 0.0001$. These levels were determined using pairwise comparisons via the Brunner-Munzel test. (b) Similarly, in a separate analysis, radiologists were given 70 randomly selected cases with a confirmed dementia diagnosis encompassing individual-level demographics and multi-sequence MRI scans. The radiologists were tasked with assigning confidence scores for the 10 dementia etiologies. Similar to that of (a), the visual representation consists of boxplots and scatterplots that represent the distribution of model and radiologists' consensus confidence scores for true negative and true positive cases. Brunner-Munzel statistical test results are shown as pairwise annotations of ns, *, **, ***, or ****.

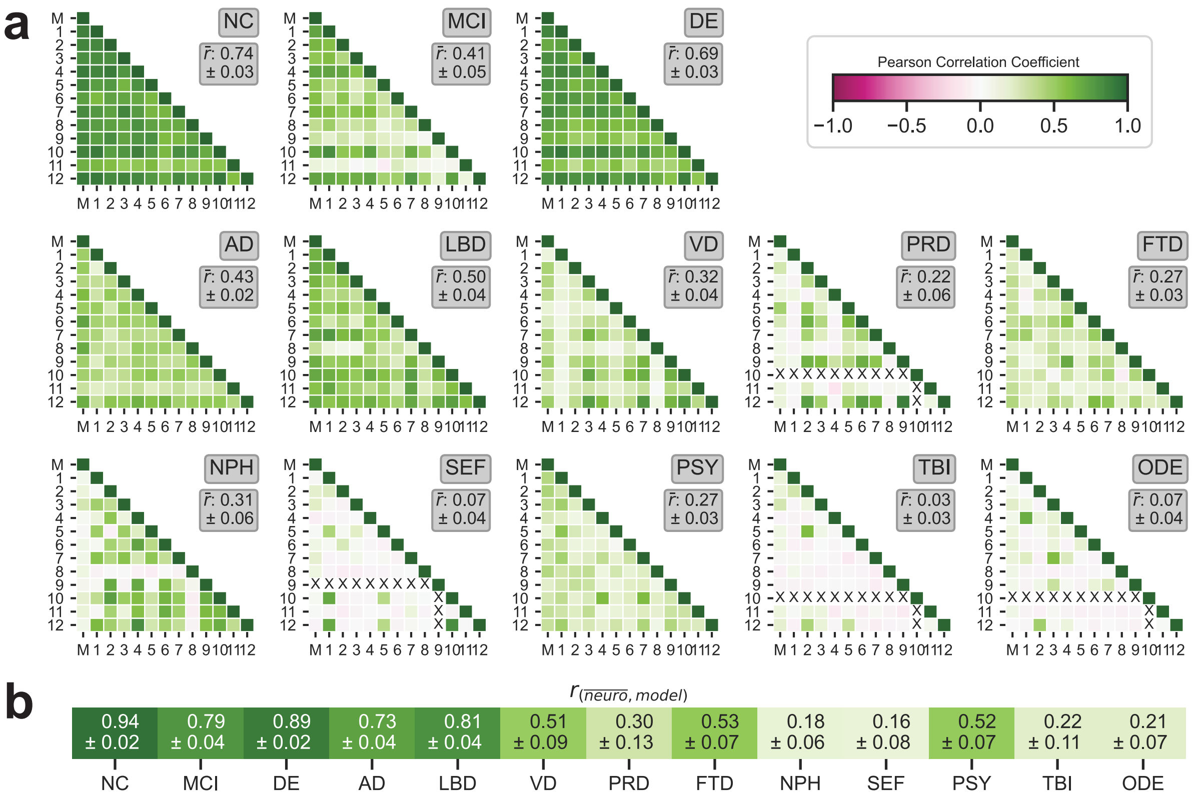


Figure S3: **Neurologist and model interrater agreement.** (a) The figure presents the Pearson correlation coefficient across different diagnostic categories, comparing assessments from the neurologists ($n = 12$) and the model, marked as 'M'. Each diagnostic category from NC to ODE includes a matrix reflecting correlation coefficient values between individual neurologists and the model. Shades of green signify positive correlation, indicating agreement between the model and neurologists, whereas magenta shades suggest negative correlations, indicating potential discrepancies in assessments. The mean pairwise Pearson correlation coefficient for each etiology is presented along with a $95\%$ confidence interval. The symbol 'X' denotes rater pairs where the Pearson correlation was not calculable, due to one or both raters giving label-specific confidence scores with no variance. (b) The heatmap shows the mean Pearson correlation coefficients between model probabilities and neurologist confidence scores for each label. The correlation coefficient and its confidence interval for each etiology was estimated with a non-parametric bootstrapping approach.

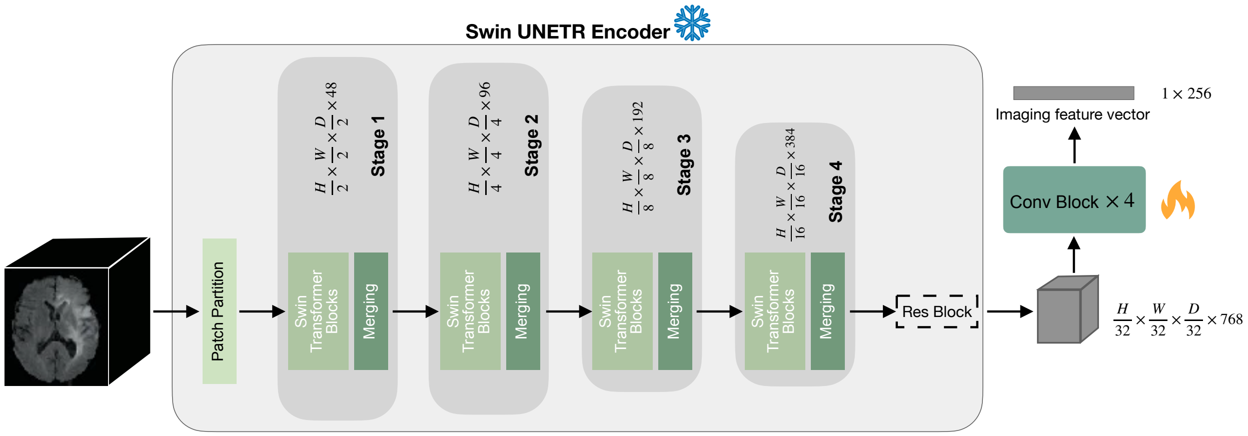


Figure S4: **Image feature extraction**. The Swin UNETR encoder, utilizing pre-trained weights, was leveraged to extract image embeddings from multi-sequence MRI scans into a latent space representation. Subsequently, these embeddings underwent a series of downsampling convolutional operations to achieve a condensed token dimension of $1 \times 256$. This dimensional reduction facilitated a consistent input format for both imaging and non-imaging data into the backbone transformer. Within this architecture, the Swin UNETR encoder's weights remained static (frozen), ensuring the integrity of the pre-trained features, while the downsampling blocks were subject to optimization during the training phase, allowing for adaptive learning of the imaging feature vector.

### Neurologist approach to the ratings

**Neurologist 1** The clinical data was reviewed initially, taking note of potential contributors such as extreme age or education (e.g., age $> 90$, education less than 9 grades), primary language, and language of cognitive testing. Pertinent factors like a history of TIA or Stroke, Parkinson's disease (PD) diagnosis and/or PD medication usage, known genetic mutations, closed head injury, alcohol or substance use disorders, chronic psychiatric symptoms/disorders, and APOE genotype were assessed. Next, the current level of functional abilities was evaluated from the provided initial description (e.g., independent living, requiring assistance with some or all activities) and Functional Activities Questionnaire (FAQ) responses. FAQ scores of 9 or higher typically indicated significant limitations with instrumental activities of daily living, supporting a dementia diagnosis. FAQ scores ranging from 4 to 8 would align with mild cognitive impairment (MCI) if cognitive test scores indicated cognitive decline. Subsequently, cognitive test scores were reviewed, with focus on age, education, and gender-adjusted z-scores. For those with normal cognition, no z-scores deviated by 1 standard deviation below the mean (i.e., no score of -1.0 or worse). Persons with MCI would exhibit at least one z-score of -1.5 or worse (e.g., -1.75) or two scores of -1.0 in the same cognitive domain. Persons with dementia would typically present with two or more scores at -2.0 or worse. Interpretation for patients with very low education or non-native language cognitive testing was approached cautiously. Following this, brain MRIs (T1-weighted images) were reviewed for signs of atrophy, the pattern of atrophy, and cerebrovascular disease. When available, diffusion-weighted imaging was used to identify a diffusion restriction pattern commonly seen in prion diseases. Functional abilities and cognitive test scores were used to classify persons as normal, MCI, or dementia. For persons between categories, a continuum scale was employed. For instance, a score of 80 for MCI and 20 for dementia would indicate an $80\%$ likelihood of classification as MCI and a $20\%$ likelihood of classification as dementia. For individuals with MCI or dementia, the most likely diagnostic category or categories were selected. In cases of mixed dementia or unclear causation, multiple diagnostic categories were chosen, with their scores summing to 100. Each category's score reflected the estimated contribution and, for mixed dementias, the extent of their contribution. For example, a score of 70 for Alzheimer's disease (AD), 20 for Lewy body dementia (LBD), and 10 for vascular dementia would signify an estimated $70\%$ contribution from AD, $20\%$ from LBD, and $10\%$ from cerebrovascular disease.

**Neurologist 2** The evaluation of case reports began with a comprehensive analysis of demographics, available medical history, apoE4 status, structured family history, and an assessment of the patient's level of functional independence. Subsequently, a thorough examination of corresponding clinical scales and neuropsychological test results was conducted. Careful observations were made regarding the subject's educational background, the presence of visual or hearing impairments, and whether the tests were conducted in the subject's native language. Following this, the synthesis of clinical data allowed for the prediction of the presence of mild cognitive impairment (MCI), dementia, or cognitive states falling below the MCI threshold, often referred to as 'normal'cognition. These predictions were quantified, with the most probable diagnosis assigned a rating exceeding $50\%$, while the others received lower ratings, reflecting the confidence in the diagnosis. Subsequently, the MRI sequences were examined alongside the case report to identify factors contributing to the patient's clinical condition. Distinctly, findings such as medial temporal atrophy and parietal atrophy were prominently associated with Alzheimer's disease, whereas the presence of flair hyperintensity and focal encephalomalacia without an alternative cause was considered indicative of vascular burden and/or dementia, especially when accompanied by deep and/or brainstem microhemorrhages. Brainstem atrophy was frequently observed in cases suggestive of potential stroke or Lewy body conditions, and the use of DWI sequences allowed for the potential identification of conditions like prion disease and epilepsy-related disorders. In assessing the clinical significance of these contributors, the most plausible factors were rated highest, while other contributors received lower but still significant ratings, typically exceeding $50\%$. However, distinguishing psychiatric features stemming from a neurodegenerative process from those arising as independent comorbid issues occasionally posed a challenge. Importantly, observed vascular burden in imaging, even when it didn't independently warrant a dementia diagnosis, was consistently acknowledged under the vascular category, often rated highly due to the confidence in its clinical significance.

**Neurologist 3** In the approach to differential diagnosis for dementia, a detailed case overview encompassed a wide spectrum of clinical information including demographics, vitals, and comprehensive personal and medical histories, alongside results from systematic physical, neurological, psychiatric, and neuro-cognitive evaluations. Cognitive function was assessed using clinician impressions from neuropsychiatric evaluations and standardized testing with MMSE or MoCA, facilitating the distinction between normal cognition, mild cognitive impairment, and dementia. Functional assessments provided insights into the impact of neurological disorders on daily living activities. Specific scales and questionnaires, such as the Hachinski Ischemic Score, evaluations for Progressive Supranuclear Palsy (PSP), and Corticobasal Syndrome (CBS), the Unified Parkinson's Disease Rating Scale, and the Neuropsychiatric Inventory Questionnaire, were instrumental in identifying localized or generalized neurological deficits, signs and symptoms of Parkinson's disease and related conditions, and characteristic features of Lewy body dementia, such as visual hallucinations. The presence of typical symptoms for disorders like normal pressure hydrocephalus also contributed to fine-tuning the differential diagnosis. The Geriatric Depression Scale was employed to discern if primary psychiatric disorders might mimic dementia presentations. An extensive review of neurocognitive testing data aided in differentiating Alzheimer's disease from other cognitive disorders. Detailed MRI analyses, revealing anomalies such as cortical atrophy, ischemic changes, and ventriculomegaly, further refined the diagnostic process.

**Neurologist 4** The patient's cognitive status, ranging from normal cognition to mild cognitive impairment (MCI) or dementia, was primarily determined based on neuropsychiatric test results and the functional assessment scale. Special consideration was given to patients with Parkinson's syndrome, as their movement disorders could impact functional assessment scores. When neuropsychiatric testing clearly indicated dementia, diagnosis was straightforward. However, cases teetering on the borderline between MCI and AD required a closer examination, where functional assessment scores, medical history, and physical examination findings were collectively considered, factoring in the influence of motor disorders on the assessment. This process involved adjusting the probability estimate based on clinical judgment. Regarding etiological diagnosis, a comprehensive evaluation was carried out, taking into account both medical history and imaging data. Cases presenting with Parkinson's symptoms led to differential diagnoses that included Parkinson's disease dementia, dementia with Lewy bodies, corticobasal degeneration, progressive supranuclear palsy, and others. In instances where imaging revealed markers of cerebral small vessel disease, the possibility of vascular dementia was explored. Notably, when prominent mental symptoms were coupled with significant atrophy in one side of the frontal and temporal lobes, consideration was given to frontotemporal degeneration. Infectious, metabolic, traumatic, and hereditary causes were also taken into account, guided by the relevant medical history. The adjustment of probability in these cases was guided by personal judgment.

**Neurologist 5** The assessment combined insights from clinical and medication history, specific neurological examinations, and neuropsychological test scores. Initially, attention was given to basic demographic data, such as age and the subject's living situation. Subsequently, a comprehensive evaluation of medical and social history was conducted, considering potential dementia risk factors and relevant habits. The presence or absence of ApoE alleles was noted. Medication history was scrutinized, particularly medications associated with vascular co-morbidities like antihypertensives and anticoagulants, indicative of vascular disease risk. The presence of antidepressants was acknowledged, considering potential psychiatric conditions linked to cognitive decline. During the review of neurological examinations, focus was placed on gaze, tremor, parkinsonism, and gait assessment. Neuropsychological examination scores were analyzed, first taking note of the number of abnormal tests. MoCA scores were used when available, alongside other tests like WMS. Language assessment, often relying on Animals and Digit span backwards, played a crucial role. Z scores and absolute scores were considered for test abnormality determination. Cognitive decline characterized by language and memory loss pointed to Alzheimer's disease. The presence of hallucinations and parkinsonism suggested Lewy body dementia, or if Parkinson's disease (PD) was advanced, it pointed to PD dementia. Executive dysfunction and disinhibition were signs of frontotemporal dementia. Hydrocephalus-associated urinary symptoms and specific findings hinted at normal pressure hydrocephalus. Mild cognitive impairment (MCI) was identified through mildly abnormal tests and preserved daily activities. MRIs were considered, yet clinical synopsis took precedence when imaging findings did not align with the clinical scenario. In offering a final diagnosis, a single label was assigned in cases of diagnosis confidence, while multiple labels were used if overlapping symptoms or psychiatric co-morbidities/alcoholism could obscure the presentation. In such scenarios, several labels were assigned with varying confidence levels. For instance, in equivocal cases of dementia and MCI, ratings were employed to determine the likelihood of each diagnosis. If both MCI and dementia were considered, dropdowns for each dementia subtype were used to indicate the more probable dementia type. When distinguishing between dementia and psychiatric conditions or acute encephalopathy proved challenging, all relevant options were marked alongside dementia.

**Neurologist 6** In assessing clinical cases for dementia, the process began with a comprehensive review of key demographic and historical data, encompassing details like age, gender, educational background, family history, and existing medical comorbidities, to provide context for interpreting the cognitive presentation. The clinical records were systematically examined, with a specific focus on the critical domains relevant to diagnosing dementia syndromes. Key tools for initial assessment, such as the Mini-Mental State Exam (MMSE) and the Montreal Cognitive Assessment (MoCA) scores, provided an initial screening of the severity and pattern of cognitive impairment. Very low scores indicated advanced dementia, while higher scores within the mild impairment range prompted a more detailed review of neuropsychological test data. This battery of neurocognitive tests revealed the specific profile of cognitive deficits within domains such as memory, language, executive function, and visuospatial abilities, each of which hinted at potential etiologies. A fundamental component of the diagnostic process involved evaluating for any concurrent neurological signs, which entailed a meticulous examination of physical findings, with a particular focus on motor exam results, including assessments for rigidity, tremors, and gait disorders often associated with Parkinsonian disorders. Additionally, the Hachinski Ischemic Scale score was considered for insights into potential vascular contributions. Furthermore, it was imperative to observe the individual's functional status and any neuropsychiatric symptoms, as they bore diagnostic and prognostic significance. The clinician had to ascertain whether the deficits impeded daily activities. Behavioral manifestations such as depression, hallucinations, delusions, and agitation could provide critical distinctions between various dementia types. Once these key components were systematically reviewed, the clinician synthesized the data to formulate a comprehensive differential diagnosis. Cognitive testing profiles, behavioral presentation, family history, age of onset, and the presence of neurological signs were all weighed and considered in a holistic manner. Common differentials in dementia assessment included Alzheimer's disease, vascular cognitive impairment, dementia with Lewy bodies, Parkinson's disease dementia, and frontotemporal lobar degeneration. Lastly, the MRI results were scrutinized for any uncommon findings that could either support or contradict the differential diagnosis. This involved assessing major structural abnormalities or alterations, such as hydrocephalus or severe atrophy, which could provide further backing for the final diagnosis.

**Neurologist 7** The interpretation method followed a structured approach. Initially, cognitive impairment severity—whether normal, mild cognitive impairment (MCI), or dementia—was determined by assessing Functional Assessment Scale Score, independence level, and neuropsychiatric testing. This assessment incorporated past medical history to exclude other potential causes of functional limitations. Etiology assessment comprised several considerations. Vascular dementia was diagnosed when factors such as stroke history, cerebrovascular disease risk factors, focal neurological deficits, Hachinski infarction score, and specific MRI findings indicating infarctions, white matter hyperintensities, and perivascular spaces were present. Parkinsonism, as evaluated by the Unified Parkinson's Disease Rating Scale (UPDRS), prompted investigation for Lewy body dementia (LBD), normal pressure hydrocephalus (NPH), vascular dementia (VD), frontotemporal lobar degeneration (FTD), and variants. LBD was considered for cases with visual hallucinations, Parkinsonism, cognitive impairment, and unremarkable MRI findings, while NPH diagnosis hinged on ventricular dilation and radiological features. FTD identification relied on executive function deficits, abnormal behavior, language impairment, and MRI-documented frontal/temporal lobe atrophy. Mental illness was contemplated for individuals with relevant medical history and substantial neuropsychiatric inventory and geriatric depression scale (GDS) symptoms. Prion disease recognition was based on distinctive MRI patterns. Conditions like infectious, metabolic, substance abuse, delirium, and psychiatric disorders were considered through medical history, coupled with the absence of specific MRI abnormalities. Lastly, multiple system atrophy (MSA) was diagnosed in cases displaying Parkinson's symptoms, defecation issues, ataxia, and cerebellar atrophy on MRI, while traumatic brain injury (TBI) diagnosis was associated with head trauma history, cognitive decline, localized lesions, and secondary atrophy.

**Neurologist 8** The evaluation process initiated with a comprehensive assessment of patient demographics, medical/family history, and risk factors. Cardiovascular and cerebrovascular risk factors were scrutinized due to their potential contribution to vascular dementia and vascular parkinsonism. Special attention was given to assessing activities of daily living (ADLs), which served as a crucial factor distinguishing dementia from mild cognitive impairment (MCI). APOE status played a pivotal role in gauging the likelihood of Alzheimer's Dementia (AD). The presence of APOE4 heightened the risk of AD, particularly in early onset cases, while APOE2 could potentially serve as a protective factor. Psychiatric history was examined to identify behavioral changes and assess whether conditions like depression or anxiety contributed to cognitive symptoms. The Geriatric Depression Scale (GDS) helped differentiate between pseudodementia/depression and other psychiatric illnesses affecting cognitive function. This information was crucial in pinpointing specific cognitive disorders (e.g., Parkinson's disease dementia, behavioral variant frontotemporal dementia (FTD), impulse control disorders in the context of dopamine agonists). A meticulous examination of clinical findings focused on gait, tremor, and bradykinesia. The presence of rest tremor, bradykinesia, or rigidity prompted consideration of parkinsonism, or other forms of parkinsonism such as Dementia with Lewy Bodies (DLB), progressive supranuclear palsy (PSP), or FTD. Comprehensive neuropsychological battery results were analyzed to discern patterns of cognitive impairment, differentiating between executive function deficits and memory impairments. Deviations in tasks such as Trails suggested executive dysfunction, potentially indicating subcortical dementia like DLB, PDD, vascular dementia, or vascular parkinsonism. Poor performance on WAIS-R or WAIS-III indicated memory impairment, typically associated with cortical dementias like AD. Imaging studies were instrumental in the evaluation. Patterns like diffuse or parietal atrophy suggested AD, while frontal-temporal atrophy indicated FTD. The presence of widespread white matter disease (WMD) burden aligned with vascular dementia or vascular parkinsonism. Specific assessments included the evaluation of the swallow tail sign, associated with PD, and midbrain atrophy, assessed through sagittal images using the midbrain-to-pons ratio (midbrain area/pontine area). Regarding the rating system, no cases received a perfect score of 100, as most presented with mixed pathologies, combining features such as amyloid beta AD changes and alpha-synuclein aggregates with parkinsonism or alpha-synuclein alongside evidence of tauopathy in PD-PSP variants. Ratings between 50% and 80% indicated varying degrees of likelihood for a specific pathology, with ratings above 80% signifying a stronger likelihood of the disease or pathology being present.

**Neurologist 9** The assessment began with a thorough review of the individual's medical history, with a focus on identifying major diagnoses that could impact cognition. This included conditions like traumatic brain injury (TBI), significant psychiatric disorders, stroke-related issues, and apoE status. Subsequently, the individual's medication history was analyzed, considering potential biases introduced by medications commonly used for Alzheimer's disease (AD) or Parkinson's disease (PD), which might have implied a higher likelihood of these conditions. Functional status assessment followed, encompassing activities of daily living (ADL) and instrumental activities of daily living (iADL), providing insights into the individual's everyday capabilities. A comprehensive physical examination was conducted, emphasizing the identification of notable abnormalities that could offer insights into cognitive status. Psychiatric and cognitive testing scales were administered, and the results were carefully analyzed for consistency and coherence. These results were also cross-referenced with the person's reported functional status. In cases of significant discrepancy, consideration was given to underlying mood or psychiatric disorders that may have influenced information accuracy. Chronology of symptoms, often absent from person-level histories, was evaluated with a particular focus on the Neuropsychiatric Inventory Questionnaire (NPI-Q), which inquired about symptoms experienced within the last 30 days. During the review of imaging studies, the gathered information was taken into account. Attention was paid to imaging findings that may have indicated AD or significant vascular disease. Unusual symptoms in the person-level history, such as new motor problems or agitation, prompted consideration of rare conditions like frontotemporal dementia (FTD), Huntington's disease, or Creutzfeldt-Jakob disease. Subsequently, a detailed review of the imaging data was conducted to identify specific features that could be indicative of these particular disorders. Lastly, the interpretation of cognitive testing scale results was influenced significantly by the individual's functional status. This guided the determination of whether the person exhibited signs of dementia, mild cognitive impairment (MCI), or fell within the spectrum of normal cognitive function. The aim was to construct a comprehensive assessment of the individual's cognitive state, accounting for these factors.

**Neurologist 10** The determination of cognitive status, including normal cognition, mild cognitive impairment (MCI), or dementia, relied primarily on neuropsychiatric test outcomes and the functional assessment scale. Notably, when individuals exhibited Parkinsonism, functional abilities were often influenced by motor impairments, making neuropsychiatric test results more influential than the Functional Activities Questionnaire (FAQ). Given the absence of distinct cutoff points for these categories, adjustments to the probability assessment were made based on individual judgment. Regarding the etiological diagnosis, a comprehensive evaluation incorporated all available clinical information and imaging data. For instance, cases presenting with Parkinsonism prompted a focused differential diagnosis that considered conditions like Dementia with Lewy Bodies (DLB), characterized by symptoms such as Parkinsonism, dementia, and hallucinations. Others included Parkinson's disease dementia (PDD), typically occurring after a prolonged history of Parkinson's disease, vascular injuries with attention to severe small vessel disease, especially within the basal ganglia, and normal pressure hydrocephalus (NPH), identified by enlarged brain ventricles. Conditions such as corticobasal degeneration (CBD) and progressive supranuclear palsy (PSP), though less common, required the presence of more typical symptoms like apraxia in CBD or abnormal vertical eye movement in PSP for diagnosis. For individuals diagnosed with MCI or dementia but without Parkinsonism, the differential diagnosis primarily encompassed Alzheimer's disease (AD), frontotemporal dementia (FTD), and vascular injuries. FTD, for example, might exhibit pronounced non-memory impairments, along with psychiatric and behavioral symptoms, and asymmetrical brain atrophy in frontal and/or temporal lobes. Additionally, vascular injuries played a substantial role in cognitive impairment and sometimes coexisted with AD pathology. In these instances, probability assessments were adjusted based on clinical judgment. For the remaining etiologies, establishing a diagnosis necessitated a detailed clinical history.

**Neurologist 11** The evaluation process initiated with an assessment of the provided case profiles, encompassing baseline information like age, education, language, and required assistance. Supplementary data, including genetic test results such as apoe4 status, medication records, and relevant details, were also considered. Subsequently, various cognitive and physical examinations, along with associated indices, were reviewed to detect neurocognitive dysfunction. From these comprehensive case profiles, preliminary hypotheses were formulated to guide the diagnostic process, ultimately leading to specific diagnoses or a set of potential options. A meticulous evaluation of imaging studies for each case followed, examining different sequences and views for signs of cerebral atrophy or structural changes, including white matter disease. These imaging findings were correlated with case profile hypotheses to generate a list of probable diagnoses. Probability ratings were assigned to these diagnoses, reflecting the likelihood of their presence. The rating process initially involved determining whether cases met criteria for normal cognition, mild cognitive impairment (MCI), or dementia. In ambiguous cases distinguishing between dementia and MCI, probability ratings were provided for both, especially when the differentiation between MCI and mild dementia was uncertain based on testing outcomes. Subsequently, probable contributing factors to the diagnoses were identified by selecting the types of dementia most likely present. Many cases presented with multiple potential contributing causes, often including vascular dementia alongside Alzheimer's disease. Quantifying the likelihood of each diagnosis involved assigning scores of 70 or higher to those with a high probability, regardless of an individual factor's relatively low contribution to their dementia. Higher scores indicated a greater likelihood of that diagnosis being the primary cause. Causes with similar probabilities scores did not reflect an equal degree of causality to the individual's condition but merely reflected an equal probability of occurrence. Scores ranging from 20 to 30 suggested the presence of dementia, though with a minor role in the clinical presentation. Scores below 10 indicated a very low probability, implying little to no significance.

**Neurologist 12** While reviewing clinical data in conjunction with MRI scans, a notable absence was observed regarding information on symptom onset and progression. This critical aspect of history-taking has the potential to offer valuable insights into the diagnosis, as the pace of progression varies among different forms of dementia. For diagnostic purposes, reliance was placed on MMSE scores, employing a cutoff of 24 to diagnose dementia. Functional capacity assessments assisted in distinguishing between MCI and dementia. Psychiatric questionnaires proved useful in orienting toward specific diagnoses, such as Parkinson's dementia, dementia with Lewy bodies (DLB), or infectious causes. The evaluation of depression's role in cognition was challenging, but the Geriatric Depression Scale provided some guidance. In cases of uncertainty, the MRI findings played a pivotal role. For instance, clear frontotemporal atrophy with behavioral disturbances and language involvement suggested frontotemporal dementia, while temporal lobe atrophy leaned more toward Alzheimer's dementia. In cases of DLB or Parkinson's dementia, clinical presentation bore more weight when MRI results were unremarkable. Moderate to severe white matter abnormalities pointed to vascular dementia. In most cases, a shortlist of potential diagnoses was compiled before reviewing the MRI. However, there were instances where MRI results were conclusive and prompted a change in the diagnosis. For example, one case indicated possible Creutzfeldt-Jakob disease due to hallucinations and corresponding MRI findings. In another, an MRI revealed significant encephalomalacia with ventricular enlargement following a head injury. A young case with a cavum septum pellucidum was attributed to chronic traumatic encephalopathy. Lastly, global atrophy in an individual with a history of alcohol abuse and seizures pointed to alcoholic dementia. Providing a percentage of certainty for each diagnosis proved beneficial, as many cases presented mixed pathology, especially in Parkinson's dementia, where vascular disease often contributed to the clinical picture.

### Neuroradiologist approach to the ratings

**Neuroradiologist 1** The evaluation of MRI scans initiated with a global perspective to exclude multiple infarcts and identify notable brain atrophy patterns. The presence and severity of white matter lesions, chronic infarcts and microhemorrhages were recorded. Subsequent assessment focused primarily on volume loss, particularly emphasizing hemispheric asymmetry. The initial evaluation determined whether dominant frontal and anterior temporal or parietal and medial temporal volume loss was evident. A more detailed sub-analysis of each region was conducted, focusing on grading severity and documenting regional and focal volume loss in real-time. The lobar volume loss evaluation was done systematically, starting with the frontal lobes, including attention to asymmetry when present. Sub-analyses of specific regions within the frontal lobes were conducted, such as the anterior insula, cingulate gyrus, precentral gyrus, and caudate nucleus. Evaluation of temporal lobe volume loss was also carried out, distinguishing mesial and non-mesial temporal lobe atrophy. Sub-analyses of hippocampal, amygdala and parahippocampal atrophy were included, with special attention to anterior, lateral, and posterior temporal lobe atrophy, including fusiform, middle, and inferior temporal gyrus volume. The assessment for atrophy was extended to parietal and occipital lobe, documenting brainstem and cerebellar atrophy. When appraising ventricular size, a comparison was made relative to sulcal size. Findings favoring an Alzheimer's disease (AD) pattern included presence of predominant parietal and medial temporal lobe atrophy, or less frontal lobe involvement than parietal and temporal lobes. Deviations from the AD pattern, such as predominant frontal, anterior temporal, or occipital involvement, enlarged ventricles, or multiple infarcts, supported non-AD dementia patterns, including those indicative of Lewy body dementia, vascular dementia, prion disease, frontotemporal lobar degeneration and its variants, normal pressure hydrocephalus, traumatic brain injury, psychiatric diagnoses, and/or other conditions. A rating scale from 0 to 100 was employed to assess the likelihood of various diagnostic considerations. A rating of 0 was selected when no evidence supported a particular diagnosis, while a rating of 100 indicated the imaging strongly suggested that entity. Ratings of 50 were assigned when imaging findings were equally likely to represent the entity in question.

**Neuroradiologist 2** The approach to rating the cases followed a systematic checklist, starting with an assessment of the entire brain, then moving through various lobes: frontal, temporal, parietal, occipital, and the brain stem. Within this framework, the aim was to determine the possible causes of dementia based on imaging findings. Initially, features indicative of normal pressure hydrocephalus were sought. These features typically stood out from other conditions and included disproportionate ventricular enlargement, an acute callosal angle at the posterior commissure level, sulcal crowding near the vertex, and Sylvian fissure enlargement. Next, the focus shifted to assessing the overall burden of white matter disease, characterized by T2 FLAIR hyperintensities. Examination was carried out in regions with encephalomalacia or gliosis, which might signify prior infarcts, helping establish a potential vascular component to dementia, either as the sole cause or a contributing factor alongside other processes. Further examination was directed toward atrophy patterns, aiming to identify specific neurodegenerative processes. Disproportionate atrophy in the medial, basal, and lateral temporal lobes and the medial parietal lobes suggested Alzheimer's disease. Relative preservation of medial temporal lobe structures hinted at dementia with Lewy bodies or Parkinson's disease dementia, although the absence of clinical history posed challenges for this diagnosis, as clinical features and typical MRI findings of medial temporal lobe preservation are valuable in a clinical setting. For frontotemporal lobar degeneration and its variants, the search was for frontal and/or temporal atrophy, predominately left posterior perisylvian or parietal atrophy, anterior temporal atrophy, predominant left posterior fronto-insular atrophy, midbrain atrophy relative to the pons ('hummingbird' sign), concavity of the dorsolateral midbrain, thinning of the tectal plate, or T2 hyperintense rim along the putamen with patchy or confluent T2 FLAIR hyperintensity in the rolandic subcortical white matter. In the quest for Prion disease indicators, examination included cortical/gyriform diffusion hyperintensity, often accompanied by thalamic and basal ganglia diffusion hyperintensity. Also explored were signs of encephalomalacia and gliosis typical of prior traumatic brain injury.

**Neuroradiologist 3** During case reviews, emphasis was placed on patient age and MRI findings as essential factors guiding the diagnostic process. Age served as a key determinant, informing the assessment of volume loss, particularly relevant in cases of Alzheimer's and frontotemporal lobar degeneration (FTLD). Each MRI sequence contributed uniquely to diagnostic considerations: T1-weighted images held importance in gauging volume loss, discerning distinctive patterns within the hippocampus, temporal lobes, and parietal lobes for Alzheimer's disease, and focusing on volume loss within the frontal and temporal lobes for FTLD. In the assessment for normal pressure hydrocephalus, attention was drawn to ventriculomegaly and its proportionality to volume loss. T1-weighted images were also instrumental in identifying cerebellar atrophy, indicative of conditions like alcoholism or phenytoin use for seizures. Diffusion-weighted images played a critical role in detecting signs of Creutzfeldt-Jakob Disease, characterized by hyperintensity in regions such as the insula, cingulate gyrus, frontal gyri, medial thalami, and possibly the basal ganglia. This sequence was also valuable for identifying infarcts. T2/FLAIR and other T2-weighted images were essential for assessing small vessel disease burden, aiding in the evaluation of vascular dementia. They were also instrumental in detecting potential evidence of infectious, inflammatory, metabolic, or drug-related hyperintensity. The susceptibility-weighted images were used to assess for microhemorrhages, which could be associated with Alzheimer's or Lewy body disease. Psychiatric diseases were typically exempt from numerical ratings as their diagnosis could not usually be ascertained through imaging. Ratings spanned from 70 to 90 in cases where a single diagnosis was highly confident. In scenarios where multiple potential diagnoses were considered, ratings ranged from 40 to 70 for each disease state, reflecting the estimated likelihood of each condition.

**Neuroradiologist 4** Each case was approached by first reviewing the demographic information; however, as the project progressed, the demographic data became less informative, and by the midpoint of the project, demographics were reviewed only as a later step. The images were assessed using the SLICER software. The T2 and FLAIR sequences were carefully evaluated to gauge the extent of small vessel disease and infarcts, serving as indicators of potential vascular causes of cognitive impairment. These sequences also proved valuable for the exclusion of infectious, inflammatory, or toxic causes. The DWI sequence was employed to identify acute infarcts and to investigate neurodegenerative conditions such as Creutzfeldt-Jakob disease or fatal familial insomnia. Susceptibility-weighted images were analyzed to identify microhemorrhages, assess their extent and location, and rule out other potential causes of cognitive decline. However, the most pivotal sequences were the volumetric sequences acquired in all three anatomical planes. They were instrumental in assessing global or lobar-specific volume loss. Specific regions of interest included the hippocampal volume assessed through coronal sequences to rule out Alzheimer's disease, the precuneus evaluated via sagittal sequences, and the parietal lobes examined in axial sequences. If frontal lobe volume loss was evident, then the temporal lobes were assessed for signs of frontotemporal dementia. Cerebellar volume loss or infratentorial volume loss led to considerations of alcohol abuse or phenytoin use, or cerebellar ataxias, while brainstem involvement indicated potential multisystem atrophy. Disproportionate ventricular dilatation raised suspicions of normal pressure hydrocephalus. The rating scale used was comprehensive, and in cases where complete information was lacking, the diagnosis was assigned to the best of the ability. A diagnosis was rated as 100 when highly confident, and as 50 when uncertainty existed. Additionally, some cases were assigned a probability score between 50 and 100 when confident in excluding other potential causes, based on the imaging data.

**Neuroradiologist 5** The approach to MR exams began with an evaluation of axial T2/FLAIR images, if available. If multiple regions of gliosis were observed alongside areas of encephalomalacia, resulting from prior infarctions in multiple vascular territories, consideration was given to the possibility of multi-infarct dementia. Moreover, when encephalomalacia and gliosis predominantly affected the temporal lobes, cerebral autosomal dominant arteriopathy with subcortical infarcts and leukoencephalopathy became a potential inclusion in the diagnostic considerations. Following the FLAIR sequence, assessment of diffusion-weighted images, if accessible, primarily served to rule out more acute conditions like Creutzfeldt-Jakob disease, herpes encephalitis, or other forms of encephalitis. Subsequently, T1-weighted images were reviewed, preferably in 3D format, to examine ventricle and sulci dimensions. The presence of ventriculomegaly and sulcal crowding at the vertex prompted consideration of normal pressure hydrocephalus as a potential diagnosis. Additionally, gyri were evaluated to identify areas exhibiting volume loss. T2-weighted images were especially helpful in this regard, as they enhanced the visibility of cerebrospinal fluid and accentuated regions of atrophy. Once the order of diagnostic differentials was established, a diagnostic rating was assigned. In this rating system, a score of 100 indicated absolute certainty, an exceedingly rare occurrence in radiology. Conversely, a score of less than 20 signified extreme unlikelihood, 25 denoted unlikeliness, 50 implied the possibility of the diagnosis, while a range of 50-75 indicated a probable diagnosis. Finally, a score exceeding 75 suggested a high likelihood of the diagnosis being accurate.

**Neuroradiologist 6** The review process began with an examination of the provided individual-level demographics for each case. Subsequently, all images provided for each case underwent analysis using the SLICER software. T2/FLAIR sequence was the basis for assessing small vessel changes, subacute to chronic infarcts, encephalomalacia from traumatic brain injury, and any areas displaying signal abnormalities indicative of potential alternative causes, such as neurodegenerative, infectious-inflammatory, or toxic-metabolic etiologies. T2/FLAIR sequence was also employed to investigate seizure-related changes. T2-weighted images played a key role in evaluating ventricular size, examining the posterior fossa for small infarcts, and observing major intracranial arterial flow voids. Diffusion-weighted images were used to identify acute infarcts and regions with reduced diffusivity, potentially linked to other neurodegenerative, infectious-inflammatory, toxic-metabolic conditions, or seizure-related changes. Susceptibility-weighted images were utilized to detect areas featuring parenchymal microhemorrhage or calcification. Lastly, high-resolution T1-weighted images were employed to analyze regional volume loss patterns suggestive of specific neurodegenerative processes. The evaluation process included the completion of the online ADRD radiologist task survey. During the assessment of sections regarding regional predominate atrophy, the high-resolution T1-weighted images were revisited to ensure response accuracy. In the final section, person-level demographics and imaging findings were synthesized to arrive at the best-guess probability for each diagnosis. The rating scale corresponded to the likelihood of the best-guess diagnosis. For instance, if there was high confidence that a case represented a particular diagnosis, it was assigned a score of 100, with a score of 0 given to all other diagnoses. In cases of diagnostic uncertainty, where the estimated probability was $50\%$, a score of 50 was assigned.

**Neuroradiologist 7** Brain volume loss was assessed based on age-appropriate norms, with T1 and T2/FLAIR sequences aiding in the evaluation of volume loss within each lobe. These sequences were particularly useful for assessing cerebrospinal fluid presence near the convexity. Brainstem volume loss was primarily evaluated through mid-sagittal and axial images, which allowed for the examination of the pontine belly and cerebral peduncle size, respectively. Coronal images provided insights into hippocampal volume, determined by the prominence of the temporal horns of the lateral ventricle. Sagittal images were employed to assess cerebellar volume loss. FLAIR sequences played a crucial role in detecting encephalomalacia, gliosis, infarcts, and white matter changes. Distinct patterns were observed in various dementia types, such as parieto-temporal volume loss favoring Alzheimer's dementia. Extensive white matter changes with or without microhemorrhages in individuals over 60 years pointed to vascular dementia. White matter changes in younger individuals raised consideration of alternative causes like infections or metabolic factors. Alcohol use often correlated with cerebellar volume loss. Traumatic brain injury was suspected in cases with FLAIR signal changes and peripheral volume loss in the anterior temporal and inferior frontal lobes, with or without susceptibility, along with corpus callosum and brainstem findings, suggestive of diffuse axonal injury. Frontal and temporal lobe volume loss indicated frontotemporal dementia. The 'hummingbird' sign on sagittal images led to consideration of progressive supranuclear palsy, particularly when combined with brainstem volume loss. Asymmetric ventricular prominence relative to cortical volume loss hinted at normal pressure hydrocephalus, with the corpus callosal angle measured on coronal images to confirm the diagnosis. While no specific findings were linked to psychiatric disorders, the presence of a cavum septum pellucidum was weakly correlated. Multiple findings in a case, such as global volume loss, extensive white matter changes and microhemorrhages, leaned toward vascular dementia over Alzheimer's disease due to the subjective nature of volume loss assessment. A higher rating was assigned to the diagnosis with more MRI findings supporting it, though no case received a perfect score of 100, with ratings exceeding 80 indicating a dominant diagnosis.

### Acknowledgments

NACC data are contributed by the NIA-funded ADRCs: P30 AG062429 (PI James Brewer, MD, PhD), P30 AG066468 (PI Oscar Lopez, MD), P30 AG062421 (PI Bradley Hyman, MD, PhD), P30 AG066509 (PI Thomas Grabowski, MD), P30 AG066514 (PI Mary Sano, PhD), P30 AG066530 (PI Helena Chui, MD), P30 AG066507 (PI Marilyn Albert, PhD), P30 AG066444 (PI John Morris, MD), P30 AG066518 (PI Jeffrey Kaye, MD), P30 AG066512 (PI Thomas Wisniewski, MD), P30 AG066462 (PI Scott Small, MD), P30 AG072979 (PI David Wolk, MD), P30 AG072972 (PI Charles DeCarli, MD), P30 AG072976 (PI Andrew Saykin, PsyD), P30 AG072975 (PI David Bennett, MD), P30 AG072978 (PI Neil Kowall, MD), P30 AG072977 (PI Robert Vassar, PhD), P30 AG066519 (PI Frank LaFerla, PhD), P30 AG062677 (PI Ronald Petersen, MD, PhD), P30 AG079280 (PI Eric Reiman, MD), P30 AG062422 (PI Gil Rabinovici, MD), P30 AG066511 (PI Allan Levey, MD, PhD), P30 AG072946 (PI Linda Van Eldik, PhD), P30 AG062715 (PI Sanjay Asthana, MD, FRCP), P30 AG072973 (PI Russell Swerdlow, MD), P30 AG066506 (PI Todd Golde, MD, PhD), P30 AG066508 (PI Stephen Strittmatter, MD, PhD), P30 AG066515 (PI Victor Henderson, MD, MS), P30 AG072947 (PI Suzanne Craft, PhD), P30 AG072931 (PI Henry Paulson, MD, PhD), P30 AG066546 (PI Sudha Seshadri, MD), P20 AG068024 (PI Erik Roberson, MD, PhD), P20 AG068053 (PI Justin Miller, PhD), P20 AG068077 (PI Gary Rosenberg, MD), P20 AG068082 (PI Angela Jefferson, PhD), P30 AG072958 (PI Heather Whitson, MD), P30 AG072959 (PI James Leverenz, MD).

ADNI is funded by the National Institute on Aging, the National Institute of Biomedical Imaging and Bioengineering, and through generous contributions from the following: AbbVie, Alzheimer's Association; Alzheimer's Drug Discovery Foundation; Araclon Biotech; BioClinica, Inc.; Biogen; Bristol-Myers Squibb Company; CereSpir, Inc.; Cogstate; Eisai Inc.; Elan Pharmaceuticals, Inc.; Eli Lilly and Company; EuroImmun; F. Hoffmann-La Roche Ltd and its affiliated company Genentech, Inc.; Fujirebio; GE Healthcare; IXICO Ltd.; Janssen Alzheimer Immunotherapy Research & Development, LLC.; Johnson & Johnson Pharmaceutical Research & Development LLC.; Lumosity; Lundbeck; Merck & Co., Inc.; Meso Scale Diagnostics, LLC.; NeuroRx Research; Neurotrack Technologies; Novartis Pharmaceuticals Corporation; Pfizer Inc.; Piramal Imaging; Servier; Takeda Pharmaceutical Company; and Transition Therapeutics. The Canadian Institutes of Health Research is providing funds to support ADNI clinical sites in Canada. Private sector contributions are facilitated by the Foundation for the National Institutes of Health (www.fnih.org). The grantee organization is the Northern California Institute for Research and Education, and the study is coordinated by the Alzheimer's Therapeutic Research Institute at the University of Southern California. ADNI data are disseminated by the Laboratory for Neuro Imaging at the University of Southern California.
